# Scalp EEG seizure warning in children under a pre-specified, locked evaluation: confound audit, structured chance levels and a negative result

**DOI:** 10.64898/2026.11.10.26367449

**Authors:** Mufeng Chen, Guojing Yu, Xiwen Lai, Pan Gong, Shuang Wang, Xiaoyan Liu, Qi Wu, Yiming Zhang, Bomiao Sun, Shenglin Xu, Jiayi Yao, Hui Zhang, Hongrui Zhang, Zeyu Wu, Jingtong Li, Quansheng Ren, Taoyun Ji

**Affiliations:** School of Electronics, Peking University, Beijing, China; Department of Pediatrics, Peking University First Hospital, Beijing, China; Aerospace Information Research Institute, Chinese Academy of Sciences, Beijing, China; Mathematical Institute, University of Oxford, Oxford, United Kingdom; School of Computer Science, Beijing University of Posts and Telecommunications, Beijing, China; Key Laboratory of Media Audio & Video (Communication University of China), Ministry of Education, Beijing 100024, China; School of Computer Science, Shanghai Jiao Tong University, Shanghai, China; College of Electronic and Optical Engineering & College of Flexible Electronics (Future Technology), Nanjing University of Posts and Telecommunications, Nanjing, China; Biology Group, The Second High School Attached to Beijing Normal University, Beijing, China; School of Information Engineering, Minzu University of China, Beijing, China; China Communications Standards Association, 52 Huayuan North Road, Haidian District, Beijing 100191, China; PKUFH-Ningxia Women & Children’s Hospital, Yinchuan, China

**Keywords:** seizure prediction, scalp EEG, statistical process control, chance level, pre-registration, paediatric epilepsy

## Abstract

**Objective:** Reported performance of scalp electroencephalographic (EEG) seizure prediction is often inflated by lenient evaluation and by the vigilance-state, clock-time and admission-trend structure of monitoring data. We asked how much short-horizon warning survives a pre-specified, locked evaluation.

**Approach:** Continuous video-EEG from 21 monitored children (1315 h) formed the development cohort. A statistical-process-control warning method was evaluated under a written protocol, then re-designed as a state-conditioned, directional scheme on seven global EEG features, frozen with a hash-recorded configuration and time-stamped pre-specification, assessed by nested leave-one-patient-out cross-validation, and applied once to two validation cohorts (a second batch from the same unit and the public CHB-MIT database from Children’s Hospital Boston; 101 evaluable seizures).

**Main results:** The original method detected 44% of evaluable seizures at 0.48 false alarms per hour, no better than a random predictor (51%, p = 0.86); its statistic moved three times more with vigilance state than with seizure approach. The frozen scheme detected 15 of 45 development seizures (33%) at 0.47 false alarms per hour, above analytic (21%, p = 0.034) and permutation chance, but a predictor using only clock time, vigilance state and days since admission reproduced most of this, and the signal was confined to the last 30 min. In validation it detected 19 seizures (19%) at 0.60 false alarms per hour against 26% chance (p = 0.96); no secondary endpoint was met, and with the child as the unit of analysis no cohort was above chance.

**Significance:** Vigilance state, clock time and admission trend inflate apparent warning performance unless the chance model preserves that structure, and the short-horizon signal did not generalise beyond its development cohort. Scalp-EEG warning should be evaluated under pre-specified rules against structured chance levels; the frozen scheme and framework are offered as a reference.

## 1. Introduction

Warning a patient or a carer minutes before a seizure has been a goal of electroencephalographic (EEG) research for decades [1–3]. Chronically implanted intracranial systems can forecast seizure likelihood better than chance in a subset of patients [4, 5]. Scalp EEG, which is available in every monitoring unit and is the only modality acceptable for most children, has a large retrospective literature reporting high sensitivity and low false-alarm rates [6, 7]. Independent audits, however, have found that most such studies leak information between training and test data or reward the recognition of patients rather than the anticipation of seizures [7–9]; that apparent preictal separability can reflect the time elapsed between recordings rather than the approach of a seizure [10]; that performance collapses across databases or when it is scored by alarms rather than by segments [11]; that prospective diary-based forecasting falls short of retrospective claims [12]; and that whether a method beats chance depends on which chance model is meant [13–15].

Three properties of continuous monitoring data make scalp-EEG warning easy to over-estimate, and they act even when cross-validation is performed correctly. First, the EEG changes with vigilance state. Features that separate sleep from wake also separate many pre-ictal from interictal segments, and false predictions cluster in sleep [16, 17]. Second, seizures follow circadian and multidien cycles [18, 19] and EEG features have diurnal profiles, so a statistic that is compared with a reference drawn from another time of day looks predictive. Multidien cycles are the basis of forecasting over days with implanted devices; the question addressed here is the complementary one of warning over minutes. Third, seizures cluster during presurgical monitoring, when medication is reduced [20], so a statistic that drifts upward over an admission gains apparent sensitivity when, as in our cohort, seizures fall late in the admission. Cross-validation removes none of these effects, because they are present in the held-out data as well; what removes them is a chance model that preserves the structure of the data, and conditioning of the statistic on the confounders.

Statistical process control (SPC) is a transparent framework for such a warning system: a monitored statistic is compared with the patient’s own recent baseline and an alarm is raised when a cumulative deviation exceeds a decision limit [21, 22]. It has been applied to seizure prediction on scalp EEG [23, 24] and on heart-rate variability [25], and separate sleep and wake references were proposed for intracranial prediction as early as 2006 [16]. What is missing is a pre-specified evaluation of such a scheme, locked before validation data are seen, compared with structure-preserving chance levels and audited for the three confounds.

Here, in continuous paediatric video-EEG, we evaluated a previously developed SPC warning method under a pre-specified protocol and found it indistinguishable from chance; we diagnosed why on the same data; we re-designed it as a state-conditional, directional SPC scheme with seven interpretable global features and froze it with a recorded configuration hash; we estimated its development-cohort performance by nested leave-one-patient-out cross-validation against four chance tests, two ablations, two published comparators and a predictor that uses only clock time, vigilance state and days since admission; and we applied the frozen scheme once, unmodified, to two validation cohorts that played no part in its development, a second batch of children from the same unit and the public CHB-MIT database. Two analyses added at revision, with the child rather than the seizure as the unit of analysis and with seizures stratified by their reported onset side, are reported as post hoc. We aim not at a clinically usable device but at establishing what survives rigorous evaluation, and at an evaluation framework that others can apply to their own methods on the same or similar data.

## 2. Materials and methods

### 2.1. Study design and pre-specification

This retrospective development and validation study follows the TRI-POD+AI structure [26] and written pre-specifications [27]. It has four stages (Fig. 1A). (1) The original method was evaluated on the development cohort (batch 1) under pre-specification v1 without deviation. The method had been developed on one pilot recording (study code IY7776, one of the 21 development children); v1 was written after inspecting the annotations and the interictal behaviour of the statistic, including its difference between sleep and wake, but before any pre-ictal value, alarm or performance figure had been seen. (2) The failure was diagnosed and the method re-designed on the same cohort with sight of it, so all development-cohort results are subject to selection. (3) The re-designed scheme was frozen on 29 September 2026: every learned quantity and every alarm constant went into a configuration file whose SHA-256 digest the runner verifies before each run, and the code constants went into a hash-recorded manifest (Supplementary Tables S1 and S6). Pre-specification v2 was written the same day, amended on 6 October and confirmed on 13 October 2026 (Supplementary Note S3); it fixes the endpoints, the decision rule and the power calculation. Both pre-specifications, the frozen configurations and the code manifest were deposited with a time-stamp before unblinding (Zenodo, doi:10.5281/zenodo.23439034; a public copy is described under Data availability). (4) The frozen scheme was run once on each validation cohort and on both cohorts pooled on 17 October 2026, after their quality control and after the deposit, with no prediction output inspected beforehand; the run log and the Zenodo time-stamp are the record. Nothing was tuned after the freeze. Two analyses added at revision, after the locked validation and after editorial review, are labelled post hoc throughout and are logged in pre-specification v2 §12.

**Figure 1:**
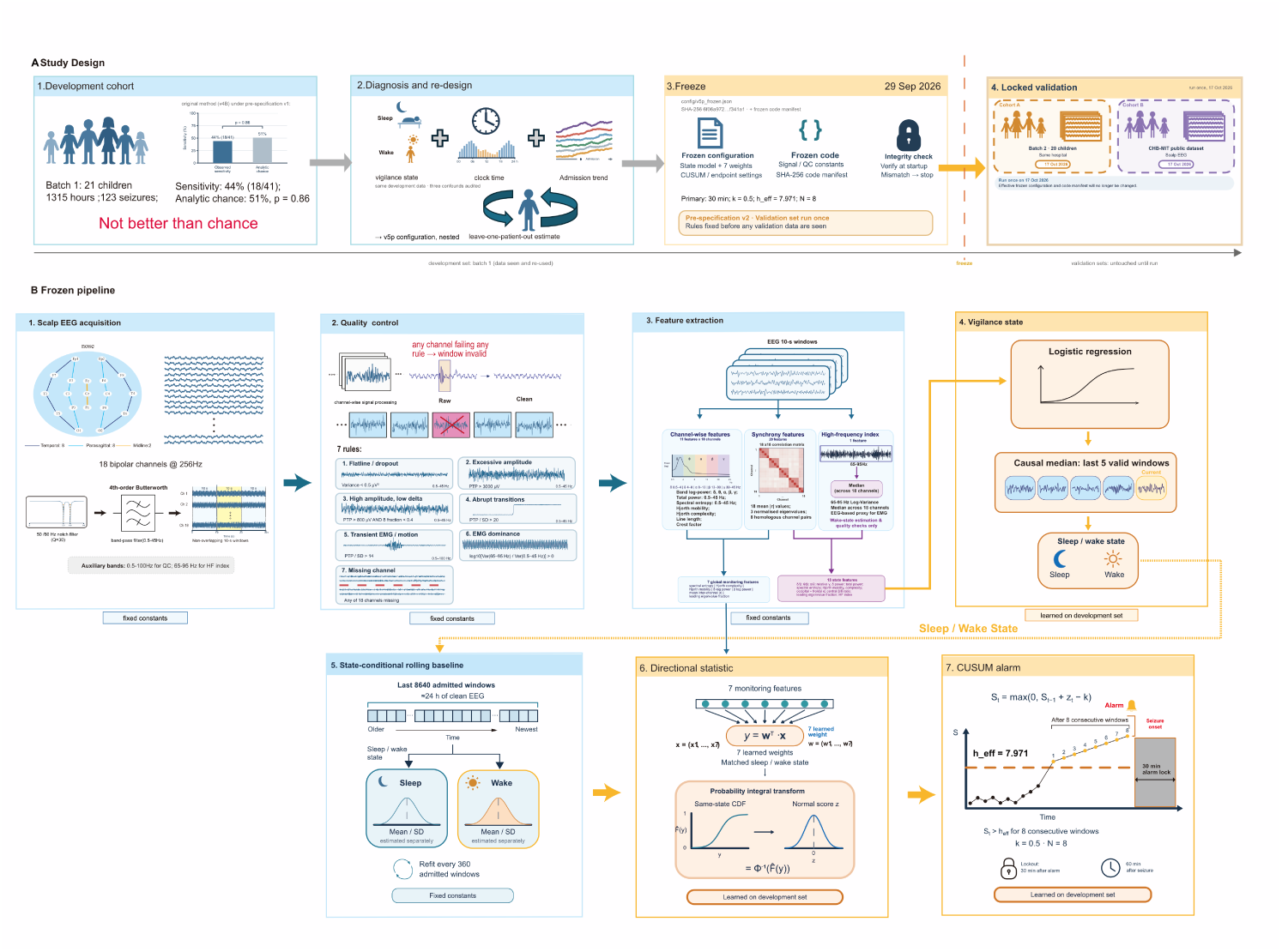
Study design and frozen pipeline. (A) Development cohort (batch 1: 21 children, 1315 h, 123 confirmed seizures) on which the original method (v4B) was evaluated under pre-specification v1 and found not better than chance (sensitivity 44%, 18 of 41, versus 51% analytic chance, p = 0.86); diagnosis and re-design on the same data, auditing three confounds (vigilance state, clock time, admission trend); freeze on 29 September 2026 (SHA-256-fingerprinted configuration, pre-specification v2); locked validation run once, on 17 October 2026, on cohort A (batch 2, 20 children) and cohort B (CHB-MIT) (dashed). (B) The seven-step frozen pipeline applied to each 10-s window; orange tags mark the three quantities learned on the development set (state coefficients, seven feature weights, CUSUM parameters); all other constants were fixed before the freeze, after inspection of the development cohort.

### 2.2. Participants, recordings and clinical characteristics

The development cohort comprised all 20 children of the unit’s first batch (June to August 2026), who were under continuous video-EEG monitoring for presurgical evaluation of focal epilepsy at the paediatric epilepsy monitoring unit of Peking University First Hospital, Beijing, together with the pilot child, who had been recorded in the same unit earlier in 2026, about four years after right hemispherotomy. The unit’s monitoring reports for all 21 children (Supplementary Table S8) give 16 boys and 5 girls with a median age of 9 years (interquartile range 5–13), an admission diagnosis of epilepsy in 20 (paroxysmal events in one), electroclinical focal seizures in all, with a left-sided electrographic onset reported in 13 children and a right-sided onset in 8, and antiseizure medication in 18 children, with dose reduction or withdrawal during monitoring documented for six. Table 1 lists, for each child, sex, age band, reported onset side, recording length, the share of windows passing quality control, calibration, seizure counts, estimated time asleep, seizure burden and the predominant side of interictal discharges. Recordings were made with a Nihon Kohden EEG-1100C system, 19 scalp electrodes in the 10–20 system with a common reference at 500 Hz, and electrocardiographic (ECG), electromyographic and other polygraphic channels; only the EEG channels enter the scheme. A physician of the unit (G.Y.) annotated every electrographic seizure onset (the gold standard), sleep and wake during routine review, and provocation procedures (hyperventilation, photic stimulation and sleep deprivation); only confirmed, spontaneous seizures were counted, and events marked with a question mark were excluded. Children are identified by study codes; to protect their privacy, individual ages are reported in five-year bands and the calendar dates of individual recordings are not given. The study was conducted in accordance with the Declaration of Helsinki and approved by the Ethics Committee of Peking University First Hospital (2021-066), which waived the requirement for informed consent for this retrospective analysis of routine recordings; patients and the public were not involved in its design, conduct or reporting.

**Table 1:**
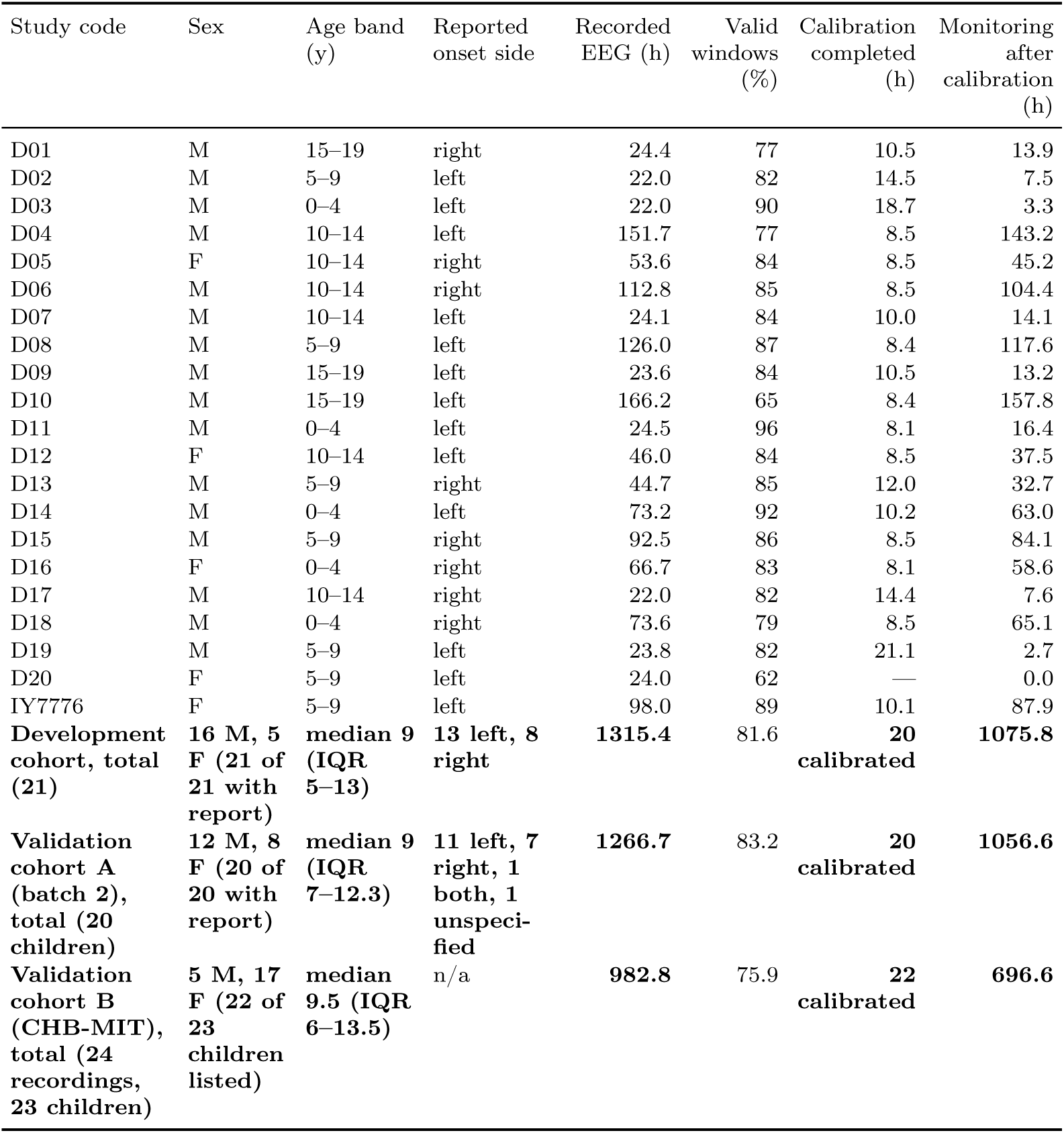
Development cohort: participants and recordings (part 1 of 2).

| Study code | Sex | Age band (y) | Reported onset side | Recorded EEG (h) | Valid windows (%) | Calibration completed (h) | Monitoring after calibration (h) |
|---|---|---|---|---|---|---|---|
| D01 | M | 15–19 | right | 24.4 | 77 | 10.5 | 13.9 |
| D02 | M | 5–9 | left | 22.0 | 82 | 14.5 | 7.5 |
| D03 | M | 0–4 | left | 22.0 | 90 | 18.7 | 3.3 |
| D04 | M | 10–14 | left | 151.7 | 77 | 8.5 | 143.2 |
| D05 | F | 10–14 | right | 53.6 | 84 | 8.5 | 45.2 |
| D06 | M | 10–14 | right | 112.8 | 85 | 8.5 | 104.4 |
| D07 | M | 10–14 | left | 24.1 | 84 | 10.0 | 14.1 |
| D08 | M | 5–9 | left | 126.0 | 87 | 8.4 | 117.6 |
| D09 | M | 15–19 | left | 23.6 | 84 | 10.5 | 13.2 |
| D10 | M | 15–19 | left | 166.2 | 65 | 8.4 | 157.8 |
| D11 | M | 0–4 | left | 24.5 | 96 | 8.1 | 16.4 |
| D12 | F | 10–14 | left | 46.0 | 84 | 8.5 | 37.5 |
| D13 | M | 5–9 | right | 44.7 | 85 | 12.0 | 32.7 |
| D14 | M | 0–4 | left | 73.2 | 92 | 10.2 | 63.0 |
| D15 | M | 5–9 | right | 92.5 | 86 | 8.5 | 84.1 |
| D16 | F | 0–4 | right | 66.7 | 83 | 8.1 | 58.6 |
| D17 | M | 10–14 | right | 22.0 | 82 | 14.4 | 7.6 |
| D18 | M | 0–4 | right | 73.6 | 79 | 8.5 | 65.1 |
| D19 | M | 5–9 | left | 23.8 | 82 | 21.1 | 2.7 |
| D20 | F | 5–9 | left | 24.0 | 62 | — | 0.0 |
| IY7776 | F | 5–9 | left | 98.0 | 89 | 10.1 | 87.9 |
| **Development cohort, total (21)** | **16 M, 5 F (21 of 21 with report)** | **median 9 (IQR 5–13)** | **13 left, 8 right** | **1315.4** | 81.6 | **20 calibrated** | **1075.8** |
| **Validation cohort A (batch 2), total (20 children)** | **12 M, 8 F (20 of 20 with report)** | **median 9 (IQR 7–12.3)** | **11 left, 7 right, 1 both, 1 unspeci-fied** | **1266.7** | 83.2 | **20 calibrated** | **1056.6** |
| **Validation cohort B (CHB-MIT), total (24 recordings, 23 children)** | **5 M, 17 F (22 of 23 children listed)** | **median 9.5 (IQR 6–13.5)** | n/a | **982.8** | 75.9 | **22 calibrated** | **696.6** |

**Table 1:** Development cohort: seizures, estimated sleep and interictal lateralisation (part 2 of 2).

| Study code | Confirmed seizures | Evaluable seizures, original rules | Evaluable seizures, frozen rules | Estimated time asleep (%) | Seizure burden | Predominant interictal lateralisation |
|---|---|---|---|---|---|---|
| D01 | 3 | 2 | 2 | 42 | sparse (< 3 per 24 h) | right 96% |
| D02 | 14 | 1 | 1 | 42 | clustered or very frequent (≥ 10 per 24 h) | left 81% |
| D03 | 12 | 0 | 0 | 41 | clustered or very frequent (≥ 10 per 24 h) | left 93% |
| D04 | 3 | 3 | 3 | 34 | sparse (< 3 per 24 h) | left 68% |
| D05 | 3 | 3 | 3 | 31 | sparse (< 3 per 24 h) | left 48% |
| D06 | 4 | 4 | 4 | 43 | sparse (< 3 per 24 h) | right 58% |
| D07 | 3 | 2 | 2 | 31 | sparse (< 3 per 24 h) | left 56% |
| D08 | 6 | 4 | 4 | 38 | sparse (< 3 per 24 h) | left 68% |
| D09 | 2 | 1 | 1 | 46 | sparse (< 3 per 24 h) | left 58% |
| D10 | 8 | 3 | 4 | 38 | sparse (< 3 per 24 h) | left 34% |
| D11 | 4 | 2 | 4 | 49 | frequent (3–10 per 24 h) | left 97% |
| D12 | 6 | 2 | 2 | 33 | frequent (3–10 per 24 h) | left 97% |
| D13 | 4 | 2 | 2 | 45 | sparse (< 3 per 24 h) | right 97% |
| D14 | 2 | 1 | 1 | 50 | sparse (< 3 per 24 h) | left 93% |
| D15 | 3 | 3 | 3 | 41 | sparse (< 3 per 24 h) | right 99% |
| D16 | 3 | 2 | 2 | 52 | sparse (< 3 per 24 h) | right 98% |
| D17 | 4 | 1 | 1 | 47 | frequent (3–10 per 24 h) | right 67% |
| D18 | 3 | 2 | 2 | 41 | sparse (< 3 per 24 h) | right 92% |
| D19 | 16 | 0 | 1 | 15 | clustered or very frequent (≥ 10 per 24 h) | left 97% |
| D20 | 15 | 0 | 0 | — | clustered or very frequent (≥ 10 per 24 h) | left 98% |
| IY7776 | 5 | 3 | 3 | 42 | sparse (< 3 per 24 h) | right 67% |
| **Development cohort, total (21)** | **123** | **41** | **45** | — | — | — |
| **Validation cohort A (batch 2), total (20 children)** | **126** | — | **51** | — | — | — |
| **Validation cohort B (CHB-MIT), total (24 recordings, 23 children)** | **198** | — | **50** | — | — | — |

Recorded EEG is the sum of file durations. Valid windows are those passing the frozen quality control (cohort value 81.6%). Calibration completes after 8 h of admissible recording with at least 4 h of valid windows; one child (D20) never completed calibration. Evaluable seizures under the original rules (90-min warning window; 41 seizures) and under the frozen rules (30-min window; 45 seizures) are counted after calibration. Estimated time asleep is from the EEG-only state estimator. Seizure burden is the clinical class assigned at review. Reported onset side is the side of electrographic seizure onset stated in the unit's monitoring report (CHB-MIT gives no side). Predominant interictal lateralisation is the side carrying the largest share of interictal discharges in the annotated review; recording gaps longer than 60 s are excluded from recorded EEG (two children). The last two rows summarise the validation cohorts (per-patient validation results in Supplementary Table S7); cohort A excludes one recording that was delivered in both batches and never completed calibration; for CHB-MIT, confirmed seizures are the annotated seizures and monitoring after calibration is summed over the 22 calibrated recordings. Sex and age at recording are transcribed from the unit's monitoring reports (Supplementary Table S8; available for all 21 development children and for all 20 cohort A children); to protect the children's privacy, age is given in five-year bands of completed years and each cohort is summarised by its median and interquartile range (IQR). For cohort B sex and age are taken from the database's public subject-information file (22 of the 23 children listed, aged 1.5–22 years; chb21 counted once, at the age of its first recording).

Validation cohort A (batch 2) comprised 21 recordings made in the same unit between July and September 2026, overlapping the last development recordings in time, and delivered after the freeze. One of them (D20: 24 h, 15 confirmed seizures, calibration never completed) proved to be a development child’s recording, recognised only after unblinding, so the cohort is 20 children (1267 h; 126 confirmed seizures, none provoked). The unit’s reports for all 20 give 12 boys and 8 girls with a median age of 9 years (interquartile range 7–12.3), a left-sided onset in 11, a right-sided onset in 7, seizures of both sides in one and an unspecified side in one (Supplementary Table S8). The cohort was processed by the same scripts as the development cohort, with only the data paths changed (Supplementary Methods S3): 83.2% of windows passed quality control, all 20 children completed calibration and 51 seizures in 19 children were evaluable; the duplicate recording contributed no monitored time and no evaluable seizure to any result. Cohort B is the public CHB-MIT Scalp EEG Database of paediatric recordings from Boston Children’s Hospital [28–30], recorded in a native bipolar montage at 256 Hz with annotated seizure onsets: 24 recordings from 23 children (chb21 is chb01 recorded 1.5 years later; both are kept as separate recordings), 686 files, 983 h and 198 annotated seizures. Its adapter was frozen on 14 October 2026 from the file headers, the annotation files and the frozen quality control alone, before any statistic had been computed (Supplementary Methods S3); 75.9% of windows passed quality control, 22 recordings completed calibration and 50 seizures in 17 recordings were evaluable. Our group had used CHB-MIT in 2026 for an unrelated deep-learning project; no constant of the frozen scheme derives from it, and it was not processed with this pipeline before the freeze.

### 2.3. Signal processing and the choice of global features

All analyses use the scalp EEG alone. Referential recordings are re-referenced to the 18-channel longitudinal bipolar montage by subtraction, which cancels the common reference; native bipolar recordings are used directly. The data are resampled to 256 Hz, notch-filtered at 50 and 60 Hz, band-pass filtered at 0.5–45 Hz (fourth-order Butterworth, zero-phase; this filter is the only non-causal step and was applied offline, whereas every later layer uses only past data) and cut into non-overlapping 10-s windows. A window is invalid if any channel fails any of seven quality rules: six signal-quality thresholds on variance, peak-to-peak amplitude, slow-wave share and high-frequency content, and a missing-channel rule (Supplementary Table S1). The thresholds were calibrated against the original method’s quality control on signal-quality measures only, so that the two schemes reject a similar share of data (81.6% of windows valid versus 80.7%; Supplementary Table S2).

The frozen scheme monitors seven global features, each a single number per window computed over all channels. Spectral entropy measures how evenly the power is spread over 0.5–45 Hz; Hjorth mobility and complexity [31] measure the dominant frequency and the bandwidth of the signal from its first and second derivatives; *δ* (0.5–4 Hz) and *β* (13–30 Hz) log power are averaged over channels; and two measures of scalp-wide synchrony are taken from the 18 *×* 18 zero-lag correlation matrix, the mean absolute inter-channel correlation and the fraction of variance carried by its first eigenvalue. Global rather than channel-wise features were chosen for three reasons. First, the scheme is meant to run on any child without patient-specific channel selection, so it must not depend on where the seizure focus lies relative to the electrodes: the children’s foci were on either side, none had intracranial localisation, and no map of each child’s epileptogenic zone exists against which channel-wise features could have been judged. Second, spectral, complexity and synchrony measures are the quantities most consistently reported to change before seizures on scalp EEG [1, 6], and the sleep–wake confound acts through the same quantities; the remedy adopted here is to condition them on vigilance state rather than to discard them. Third, among the candidate feature sets screened on the development cohort before the alarm layer was fixed (Supplementary Table S5), this set gave the highest segment-level separation between pre-ictal and interictal data, with a three-feature heart-rate statistic computed from the ECG channel serving as a negative control for EEG specificity. A consequence of this choice is that the scheme carries no spatial information: whether the proximity of the electrodes to the presumed epileptogenic zone affects detection cannot be tested with these features, and only a post hoc comparison by the onset side stated in the monitoring reports is possible (Section 3.7).

### 2.4. The original method

The original method (version 4B; Supplementary Methods S1) computed spectral entropy and Hjorth mobility per 10-s window and channel, expressed them as a squared Mahalanobis distance from the channel’s recent baseline, using a Ledoit–Wolf shrinkage estimate of the covariance [32], mapped each channel’s distance to a standard-normal score by a probability-integral transform against the baseline queue, fused the 19 channel scores with non-negative weights derived from the baseline covariance, and raised alarms from a one-sided cumulative sum (CUSUM) chart [21] when a risk transform of the chart exceeded a decision limit for a number of consecutive windows. Its warning window was 0.5–90 min and alarms and seizures were followed by a 1-h lock-out. Its three CUSUM parameters were selected per held-out patient on the other patients, from a grid of 700 combinations pruned by the Siegmund approximation to the average run length [33], under a pooled false-alarm cap of 0.5 per hour.

### 2.5. The frozen scheme

#### Vigilance state

Because sleep and wake differ more in these features than pre-ictal and interictal EEG do (Section 3.3), every window is first assigned a vigilance state and is compared only with windows of the same state. An EEG-only logistic estimator classifies each 10-s window as sleep or wake from 13 global features: the seven monitored features and six further band-power ratios and relative powers (*δ*/*β*, *θ*/*β* and *α*/*θ* ratios, relative *γ* and *δ* power and total log power; Supplementary Table S2). Each feature is first centred and scaled robustly, using the median and the median absolute deviation of that feature over the current baseline queue (the preceding 24 h of clean recording, defined below), so that the estimator sees each child’s features on the child’s own scale. The estimator’s output probability is smoothed by a causal median over the five most recent windows, and the window is labelled sleep when the smoothed probability exceeds 0.5. The coefficients were fitted on the development cohort against the clinicians’ sleep and wake marks (7727 labelled windows in 21 children). In leave-one-patient-out evaluation the estimator had a pooled area under the receiver operating characteristic curve (AUC) of 0.964 and an accuracy of 91.9%; the causal run-time version agreed with the marks in 86.9% of the labelled windows of the 20 calibrated children (an in-sample figure; Supplementary Table S2).

#### State-conditional rolling baseline

The baseline queue holds the most recent 8640 admitted windows, that is, 24 h of clean data: windows that are valid, lie outside provocation periods and outside the hour after a confirmed seizure or a recording gap. Pre-ictal windows are not excluded from the queue, because in real time they cannot be known. Before each block of 360 admitted windows (1 h), the queue is split by estimated state and the mean and standard deviation of each monitored feature are computed separately for sleep and for wake, using only data that precede the block; each new window is then standardised with the statistics of its own state. A window whose state has fewer than 300 windows in the queue is not monitored. The 24-h queue is a cap that was reached in 575 of 906 blocks and never in 8 of 20 development children (Supplementary Methods S2).

#### Directional statistic

The seven standardised features x (missing values set to 0, values clipped to *±*5) are combined into a single score y = w*·*x with seven frozen weights: +0.48 for spectral entropy, *−*0.02 for Hjorth complexity, *−*0.43 for Hjorth mobility, +0.10 for *δ* log power, +0.31 for *β* log power, *−*0.73 for the mean absolute inter-channel correlation and +0.42 for the first-eigenvalue fraction (Supplementary Table S1), so that y increases in the preictal direction. The weights are those of an L2-regularised logistic regression (C = 0.05) fitted on the 20 calibrated development children, with windows 15–0.5 min before each lead seizure as positives and windows more than 4 h from any seizure as negatives, each child weighted equally within its class (Supplementary Methods S2). The two synchrony features are highly collinear, so their individual weights should not be read physiologically; their net effect is that a fall in inter-channel synchrony raises y. The score is then mapped to a standard-normal scale, z = Φ*^−^*^1^(*F*^^^(y)), where *F*^^^ is the Blom-corrected empirical distribution [34] of y over the same-state windows of the current queue, so that interictal z is approximately standard normal within each state and one set of alarm parameters serves all patients.

#### Alarm

A one-sided CUSUM accumulates the excess of z over a reference value k, S_t = max(0, S_{t*−*1} + z_t *−* k), and raises an alarm when S has exceeded the decision limit h_eff for N consecutive valid windows. The reference value k sets the per-window excess that is allowed to accumulate, the decision limit h_eff sets how much accumulated excess is needed, and N guards against single-window spikes. After an alarm S is reset and further alarms are locked out for the length of the warning window; S is also reset at each confirmed seizure (60-min lock-out) and at recording gaps longer than 60 s, and it is held across invalid or unmonitored windows while the consecutive count restarts. The primary operating point (0.5–30 min warning window, false-alarm cap 0.5 per hour) uses k = 0.5, h_eff = 7.971 and N = 8 (80 s); secondary operating point 1 (cap 0.25 per hour) uses k = 0.75, h_eff = 7.971 and N = 8; secondary operating point 2 (0.5–20 min window, cap 0.5 per hour) uses k = 0.25, h_eff = 40.0 and N = 5. Each set was chosen on the pooled out-of-sample z series of the development children from 420 combinations pruned by the Siegmund average run length, as the combination with the highest pooled sensitivity under the cap, ties broken by the lower false-alarm rate. Only three quantities were learned from the development cohort: the state coefficients, the seven weights and the CUSUM parameters.

### 2.6. Calibration, evaluable seizures, alarm scoring and the warning window

Monitoring starts after calibration: 2880 admissible 10-s slots (8 h), of which at least 1440 must be valid, counted from the start of the recording; a seizure during calibration removes the preceding hour and suspends counting for the following hour.

A seizure is evaluable if it is confirmed (criterion E1), spontaneous (E2), at least one warning window after calibration (E3), at least 1.5 h after the preceding confirmed seizure by the annotated time stamps (E4; the 60-min lock-out plus the 30-min window) and preceded by at least 50% recorded time in the previous 90 min (E5). Time-stamp E4 was adopted in v2 after the index-based rule used during development was found to admit one seizure 5394 s after its predecessor; development results are given under both rules.

Each alarm is scored against the next confirmed seizure. An alarm raised 0.5–30 min before an evaluable seizure is true, and the seizure counts as detected; an alarm inside the window of a non-evaluable seizure, or less than 0.5 min before an onset, is neutral; all other alarms are false. Sensitivity is the proportion of evaluable seizures detected, with a Wilson 95% confidence interval [35]. The false-alarm rate is the number of false alarms per hour of alarm-eligible interictal time, which excludes warning windows, lock-outs, invalid windows and the hour after each seizure; it is therefore higher than the number of alarms per hour of recording, which is also reported. Lead time runs from the earliest alarm’s window to the onset.

The warning window of 0.5–30 min was fixed in pre-specification v2 and chosen with the unit’s clinical team. In an epilepsy monitoring unit an alarm prompts a look at the video rather than an intervention, so a warning more than about half an hour ahead has no use and would mostly reward slow drifts of the statistic; an alarm less than 30 s before the electrographic onset, on the other hand, may be triggered by the onset itself. The original method’s window of 0.5–90 min was the choice of pre-specification v1, and both are reported. How the signal evolves towards a seizure, from 8 h before the onset to the onset itself, is examined separately by horizon-resolved analyses (Section 3.5) rather than by widening the window, because a wider window adds interictal time to the target without adding signal.

### 2.7. Diagnosis of confounds, ablations and comparators

The original method’s normalised statistic was compared between clinician-marked sleep and wake, using windows within 5 min after a mark and more than 2 h from any seizure. For the frozen scheme, out-of-sample interictal z was profiled by clock time (4-h bins) and by estimated state; the position of each evaluable seizure within its child’s monitoring period and the autocorrelation of 30-min interictal block means of z were computed; and the segment AUC was recomputed with clock-matched references. Two ablations re-ran the whole nested procedure without state conditioning and with an unsigned Mahalanobis distance in place of the directional statistic. Two published approaches were re-implemented from their descriptions: multivariate SPC with T^2^ and Q charts [24], evaluated under our rules and under its published protocol, and a per-channel pre-ictal-pattern criterion [36], evaluated under its own protocol and, additionally, on seizure-free control epochs of the same recordings (Supplementary Note S4). The dependence of the signal on the horizon was explored by segment AUC at horizons from 10 min to 6 h, by disjoint rings before onset and by the percentile profile of z from 8 h before to 6 h after onset (Supplementary Note S7). Candidate feature sets were screened before the alarm layer was fixed, with the three-feature heart-rate statistic from the ECG channel as a negative control (Supplementary Table S5).

### 2.8. Statistical analysis

#### Chance references

Every performance figure is compared with four pre-specified chance references. (1) Analytic: a Poisson random predictor with the same false-alarm rate *λ* hits a warning window of length W with probability 1 *−* exp(*−λ*W), with W = 29.5 min; the number of detected seizures is tested against this probability with a one-sided binomial test [37, 38]. (2) Permutation: each patient’s alarms are re-placed uniformly over that patient’s alarm-eligible windows, keeping their number and their minimum spacing (1000 permutations) [13]. (3) Local jitter: each alarm is moved only within *±*1, *±*3 or *±*6 h of its original position (2000 permutations), which preserves the admission trend and, for *±*1 h, the diurnal profile. (4) Clock-matched: pre-ictal segments are scored only against sliding interictal blocks of the same patient that lie more than 8 h from any seizure and are centred within 2 h of the segment’s clock time, with at least five matched blocks per segment.

#### Unit of analysis

The pre-specified alarm-level tests count evaluable seizures, which are nested within children (one to four evaluable seizures per child in the development cohort, one to eight in the validation cohorts) whose false-alarm rates range from 0 to 0.92 per hour. The binomial test and the Wilson interval treat seizures as independent and are therefore anti-conservative. Three features of the design limit the problem and are reported alongside: the permutation and jitter references re-place each child’s own alarms within that child’s record, so the chance level is computed child by child; the segment-level AUC is computed per child and tested across children; and a design-effect adjustment for the clustering is given with the development result. At revision we added, post hoc, an explicit child-level analysis at the primary operating point (Supplementary Table S9): each child’s chance probability is computed from that child’s own false-alarm rate; a child-cluster bootstrap (20 000 resamples of children) gives a confidence interval for the pooled sensitivity and a p-value against the pre-specified chance level; the child-averaged sensitivity minus chance is given with the same bootstrap; an exact sign test and a one-sided Wilcoxon signed-rank test are applied to the per-child differences; and per-child one-sided binomial tests are corrected by the Benjamini–Hochberg procedure [39]. A second post hoc analysis stratifies the development and same-site validation seizures by the onset side stated in the monitoring reports (left, right, both or unspecified; CHB-MIT gives no side), comparing each side’s sensitivity with its child-wise chance level and the two sides with each other by Fisher’s exact test. Neither analysis was pre-specified; both are logged in pre-specification v2 §12 and change no pre-specified decision.

#### Endpoints

The primary endpoint of v2 is sensitivity at the primary operating point above the analytic chance level, with p < 0.05 on both the analytic and the *±*1 h jitter test; the uniform permutation is reported along-side because it proved the most lenient model on the development cohort. Key secondary endpoints, in a fixed order and without multiplicity adjustment, are the *±*3 h jitter test; the per-patient segment-level AUC (30-min pre-ictal segments versus non-overlapping interictal segments more than 4 h from any seizure; one-sided Wilcoxon test against 0.5); the two secondary operating points against their own chance levels; the clock-matched AUC; and, added before unblinding on 15 October 2026 (v2 §12), a confound-only comparator: a logistic predictor built from two circadian harmonics of clock time, the scheme’s own vigilance estimate and the number of days since the start of the recording, fitted on the development cohort with its own nested parameter selection (k = 0.5, h_eff = 40.0, N = 8), scored by the same CUSUM on the same windows as the frozen scheme and reported without a pass-or-fail criterion (Supplementary Note S6).

#### Calibration of the tests

Before unblinding, the false-positive rate of every test was estimated on effect-free simulations that keep each child’s clock-time profile, vigilance state, admission trend, missingness, seizure times and residual autocorrelation, or that circularly shift the real series (300 replicates each; Supplementary Note S5). The primary rule and the analytic test rejected in at most 3.3% of replicates, the *±*1 h jitter test in at most 8.3%, the uniform permutation in up to 16.7% and the per-patient segment AUC in up to 35.0%; segment-level AUC p-values are therefore reported as nominal and were not used for decisions.

#### Development-cohort estimates and power

Development-cohort estimates are nested leave-one-patient-out: each held-out child’s weights and CUSUM parameters, including the parameter search, come from the other 19 children only, whereas the vigilance estimator, the quality-control thresholds and the choice among candidate feature sets were fixed with sight of the whole cohort. Horizon, ring, percentile-profile and clock-matched analyses use interictal blocks more than 8 h from any seizure and are exploratory; other tests are named where they are reported, with Benjamini–Hochberg correction for per-child segment tests (Supplementary Table S3). The sensitivity–time-in-warning curve thresholds the 5-min causal moving average of z and holds the warning for 30 min; per-patient results and the alarm-level positive predictive value are descriptive. The power of the analytic test at the development effect and at half of it (Supplementary Note S1) was 0.55 and 0.20 for the 51 seizures counted in batch 2, 0.63 and 0.26 for the 50 counted in CHB-MIT and 0.86 and 0.39 for the 101 pooled, all counted before the locked run. The pooled analysis of both validation cohorts is primary, each cohort is also reported, and no cohort may be dropped after results are seen. Analyses used Python 3.14 with NumPy, SciPy and scikit-learn.

## 3. Results

### 3.1. Development cohort

The 21 children contributed 1315 h of EEG (22–166 h each) and 123 confirmed seizures (Table 1); 81.6% of the 473 565 windows passed the frozen quality control (62–96% per child). Calibration completed in 20 of 21 children after 8.1–21.1 h. Of the 123 seizures, 108 occurred in these 20 children and 74 after calibration (none provoked); 29 of the 74 began less than 1.5 h after the preceding confirmed seizure (E4 by time stamp; 28 by the index-based rule) and one fell within the first warning window after calibration (E3), leaving 44 (45) evaluable seizures in 19 children under the frozen rules; 41 seizures were evaluable under the original method’s rules.

### 3.2. The original method was not better than chance

Under pre-specification v1 the original method detected 18 of 41 evaluable seizures (43.9%, 95% CI 29.9–59.0%) within 0.5–90 min of onset at 0.478 false alarms per hour (269 false alarms in 562 h), with a median lead of 52.9 min (Table 2). A Poisson random predictor at the same rate and with the same window would detect 51.0% (p = 0.86; 27 detections of 41 would have been needed for p < 0.05), and shuffled alarms detected 42.4% (p = 0.48). The best in-sample parameter set detected 22 of 41 (53.7%), which is the chance level.

**Table 2:** Operating points against chance: operating points and sensitivity (part 1 of 2).

| Scheme | Warning window (min) | False-alarm cap (/h) | Detected / evaluable | Sensitivity, % (Wilson 95% CI) | False alarms per hour | Median lead (min) |
|---|---|---|---|---|---|---|
| **Development cohort (nested leave-one-patient-out; 45 evaluable seizures in 19 children unless stated)** |  |  |  |  |  |  |
| Primary operating point | 0.5–30 | 0.5 | 15/45 | 33.3 (21.4–47.9) | 0.473 | 14.7 |
| Primary, strict time-stamp E4 (44 seizures) | 0.5–30 | 0.5 | 16/44 | 36.4 (23.8–51.1) | 0.483 | 12.3 |
| Secondary 1 | 0.5–30 | 0.25 | 10/45 | 22.2 (12.5–36.3) | 0.244 | 15.8 |
| Secondary 2 | 0.5–20 | 0.5 | 11/45 | 24.4 (14.2–38.7) | 0.414 | 8.7 |
| Ablation: no state conditioning | 0.5–30 | 0.5 | 11/45 | 24.4 (14.2–38.7) | 0.459 | 16.7 |
| Ablation: unsigned statistic | 0.5–30 | 0.5 | 9/45 | 20.0 (10.9–33.8) | 0.449 | 13.5 |
| Comparator: multivariate SPC (T<sup>2</sup>/Q), our rules | 0.5–30 | 0.5 | 4/45 | 8.9 (3.5–20.7) | 0.579 | 15.8 |
| Original method (v4B), its own pre-specification | 0.5–90 | 0.5 | 18/41 | 43.9 (29.9–59.0) | 0.478 | 52.9 |
| **Locked validation (frozen parameters, run once; 1000 permutations, 2000 jitter resamples)** |  |  |  |  |  |  |
| Cohort A (batch 2), primary | 0.5–30 | 0.5 | 6/51 | 11.8 (5.5–23.4) | 0.598 | 11.7 |
| Cohort B (CHB-MIT), primary | 0.5–30 | 0.5 | 13/50 | 26.0 (15.9–39.6) | 0.610 | 13.7 |
| Pooled A + B, primary | 0.5–30 | 0.5 | 19/101 | 18.8 (12.4–27.5) | 0.602 | 13.7 |
| Cohort A (batch 2), secondary 1 | 0.5–30 | 0.25 | 2/51 | 3.9 (1.1–13.2) | 0.339 | 9.2 |
| Cohort A (batch 2), secondary 2 | 0.5–20 | 0.5 | 6/51 | 11.8 (5.5–23.4) | 0.470 | 8.5 |
| Cohort B (CHB-MIT), secondary 1 | 0.5–30 | 0.25 | 9/50 | 18.0 (9.8–30.8) | 0.354 | 13.8 |
| Cohort B (CHB-MIT), secondary 2 | 0.5–20 | 0.5 | 9/50 | 18.0 (9.8–30.8) | 0.557 | 8.7 |
| Pooled A + B, secondary 1 | 0.5–30 | 0.25 | 11/101 | 10.9 (6.2–18.5) | 0.344 | 13.8 |
| Pooled A + B, secondary 2 | 0.5–20 | 0.5 | 15/101 | 14.9 (9.2–23.1) | 0.498 | 8.7 |
| Cohort A (batch 2), confound-only comparator (clock, state, days) | 0.5–30 | fixed | 10/51 | 19.6 (11.0–32.5) | 0.608 | — |
| Cohort B (CHB-MIT), confound-only comparator (clock, state, days) | 0.5–30 | fixed | 3/50 | 6.0 (2.1–16.2) | 0.457 | — |
| Pooled A + B, confound-only comparator (clock, state, days) | 0.5–30 | fixed | 13/101 | 12.9 (7.7–20.8) | 0.557 | — |

**Table 2:**
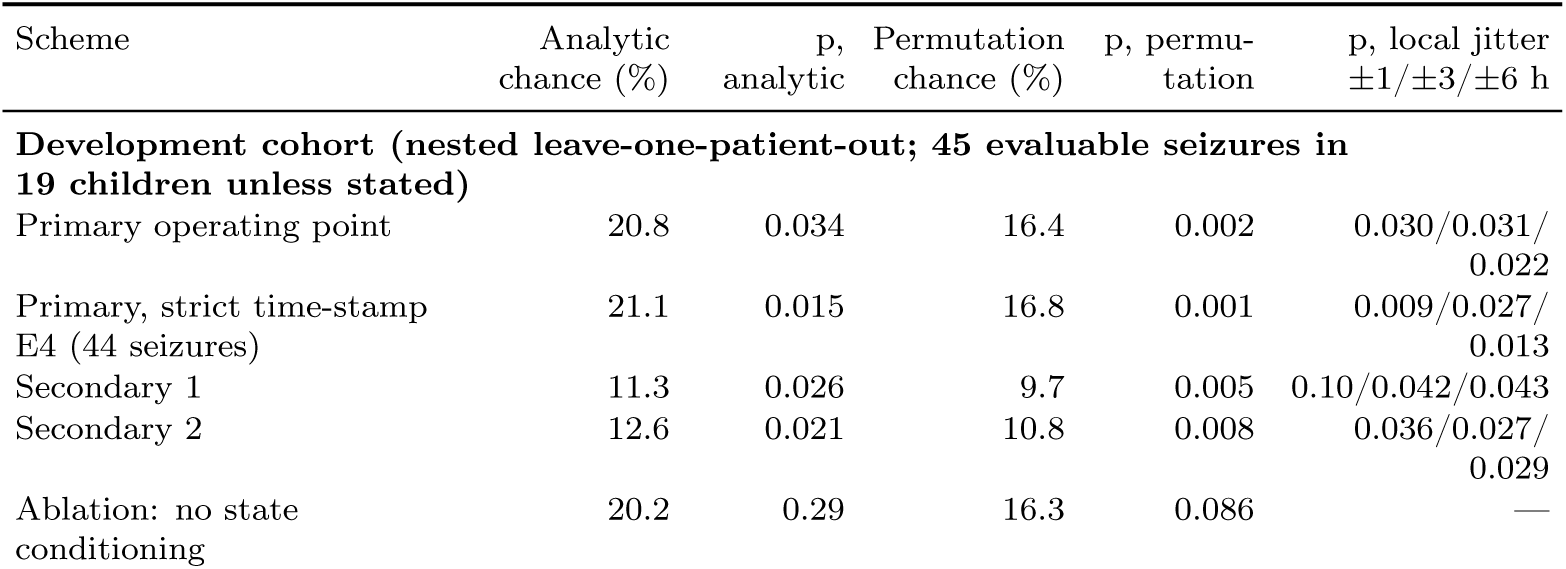
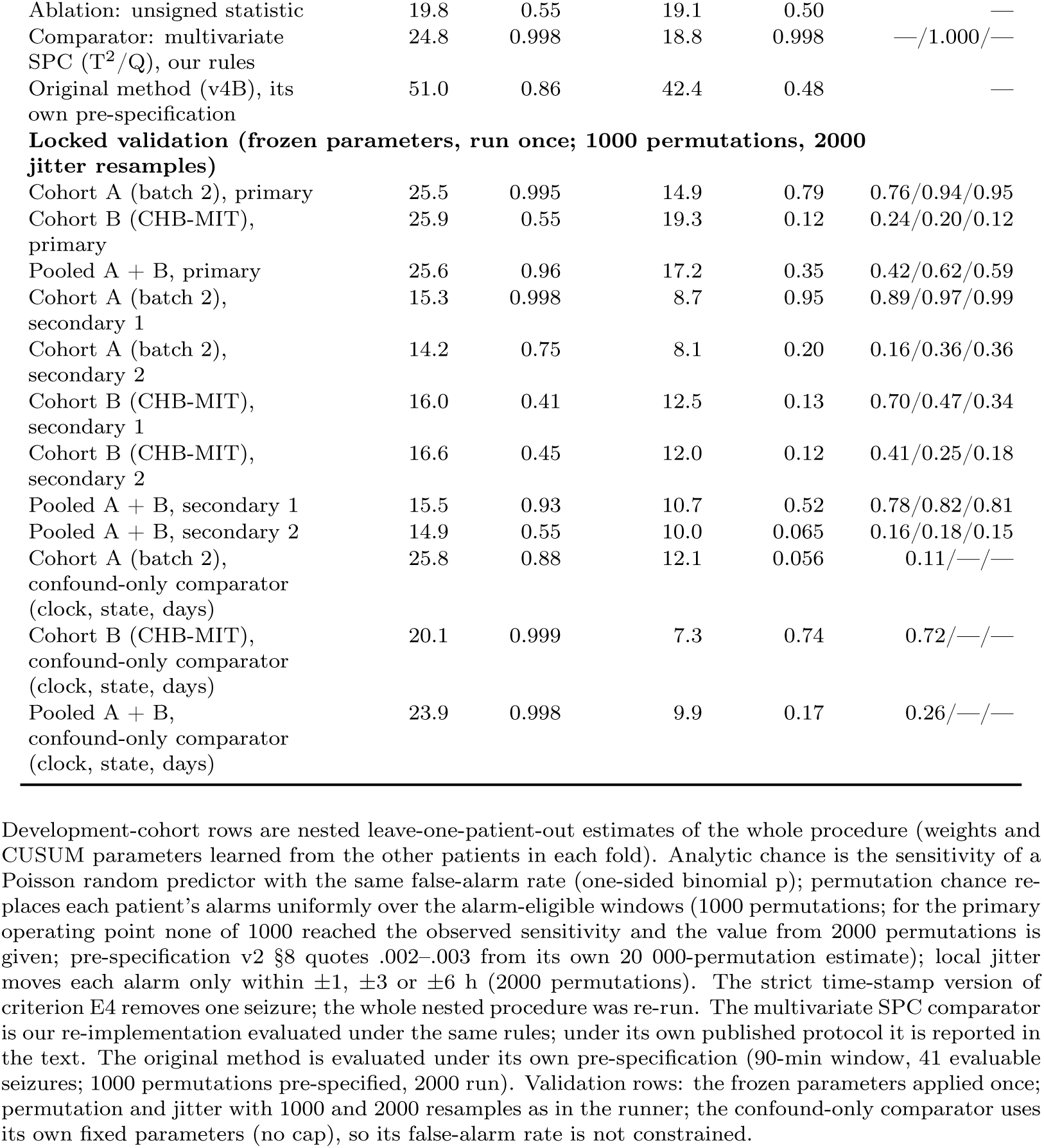
Operating points against chance: chance levels and p values (part 2 of 2).

### 3.3. Diagnosis: vigilance state, clock time and admission trend

In the original method’s normalised statistic, the 16 children with both kinds of mark had a mean of *−*0.06 in sleep and +0.56 in wake, a wake *−* sleep difference of 0.62 SD with the same sign in 13 of the 16 children (Fig. 2A; Supplementary Note S2), against a pre-ictal shift of +0.22 SD over the 41 evaluable seizures: a change of vigilance state moved the statistic by about three times the pre-ictal effect.

**Figure 2:**
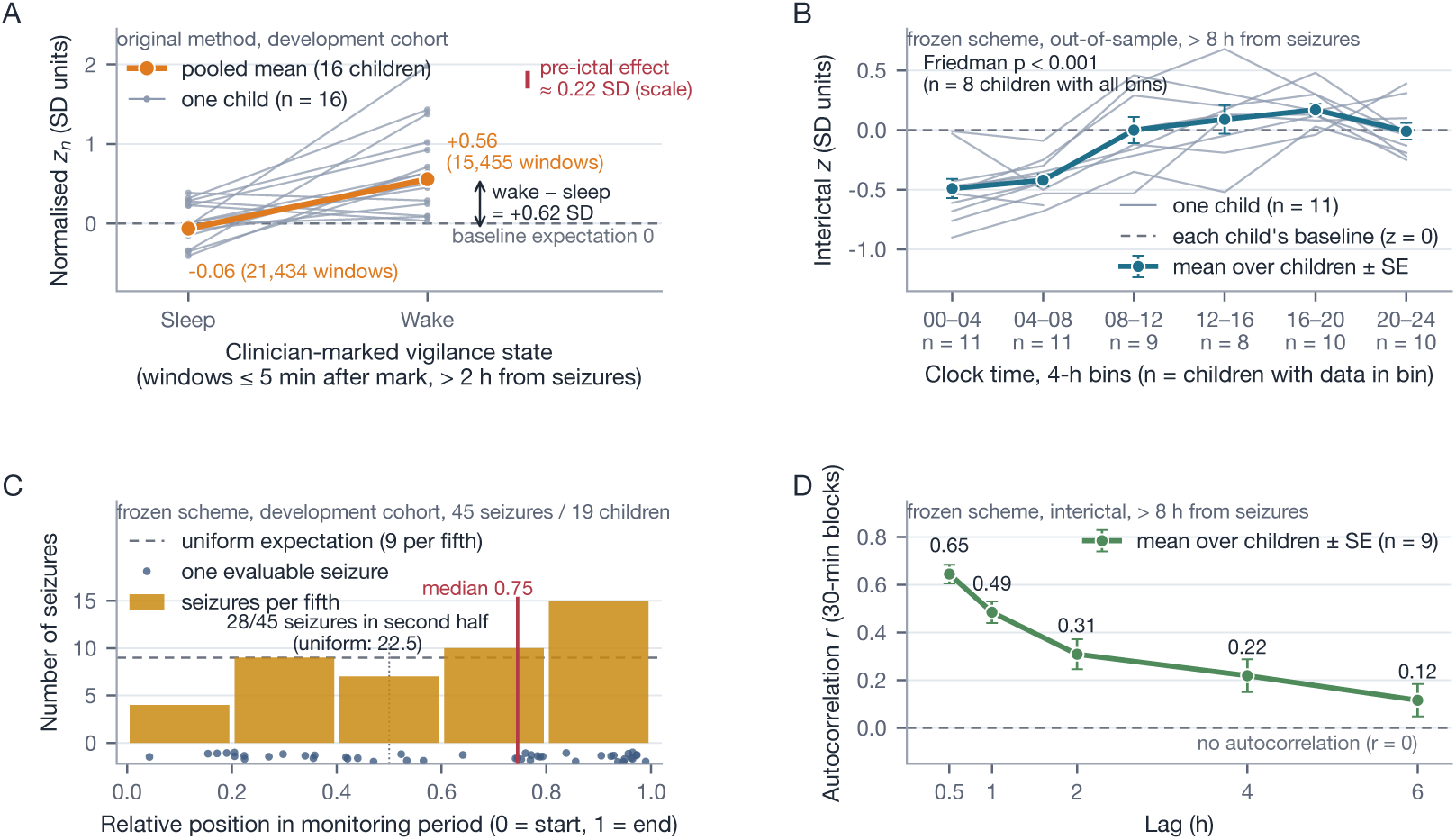
Three confounds of a scalp-EEG warning statistic (development cohort). (A) Original method: normalised statistic z_n in windows within 5 min after a clinician’s sleep or wake mark and more than 2 h from any seizure; grey lines, the 16 children with both kinds of mark; orange, their pooled means (sleep *−*0.06, 21 434 windows; wake +0.56, 15 455 windows); dashed line, baseline expectation (0); red bar, pre-ictal effect (*≈*0.22 SD) for scale. (B) Frozen scheme: out-of-sample interictal z (> 8 h from any seizure) by 4-h clock bin; grey, individual children; mean *±* SE over children; dashed, patient baseline (z = 0). (C) Relative position of the 45 evaluable seizures within each child’s monitoring period; dashed, uniform expectation. (D) Autocorrelation of interictal 30-min block means of z (mean *±* SE, 9 children); dashed, r = 0.

In the frozen scheme, state conditioning removed the state difference in out-of-sample interictal z (wake *−*0.22 versus sleep *−*0.20 in 11 children; paired Wilcoxon p = 0.58), but a clock-time profile remained (Fig. 2B): interictal z more than 8 h from any seizure averaged *−*0.49 at 00–04 h and *−*0.42 at 04–08 h against 0.00 to +0.17 during the day (Friedman test over 8 children, *χ*^2^ = 24.1, p < 0.001). Evaluable seizures fell late in the admissions (median relative position 0.75; 28 of 45 in the second half; Fig. 2C), as did alarms (58%), and the interictal 30-min block means of z were autocorrelated for hours (r = 0.65 at a lag of 0.5 h, falling to 0.12 at 6 h, in 9 children; Fig. 2D). A daytime seizure late in a recording is therefore compared with a reference that is richer in night-time and earlier-admission data, which is why a uniform permutation of alarms understates chance.

### 3.4. Development-set performance of the frozen scheme

At the primary operating point the frozen scheme detected 15 of 45 evaluable seizures (33.3%, 95% CI 21.4–47.9%) within 0.5–30 min of onset, with 324 false alarms in 684.7 h of alarm-eligible interictal time (0.473 per hour; per-patient median 0.39 per hour, range 0–0.92) and a median lead of 14.7 min (Fig. 3A; Table 2). The analytic chance level is 20.8% (p = 0.034; without the pilot child IY7776, 13 of 42 at 0.446 per hour, p = 0.056). No pre-specified uniform permutation (1000) reached 15 detections (chance level 16.4%; p = 0.002 with 2000 permutations), and local jitter of *±*1, *±*3 and *±*6 h gave chance levels of 23.7%, 22.0% and 20.7% (p = 0.030, 0.031 and 0.022). Under strict time-stamp E4 (44 seizures; the whole nested procedure re-run) the scheme detected 16 of 44 (36.4%) at 0.483 per hour (analytic p = 0.015; Table 2). The per-patient segment AUC averaged 0.672 (above 0.5 in 10 of the 13 children with sufficient data; one-sided Wilcoxon p = 0.024; child-level bootstrap 95% CI 0.53–0.81; Supplementary Table S3) and the pooled AUC was 0.641 (Fig. 3B); thresholding the smoothed statistic without the CUSUM gave 37.8% sensitivity at 20.4% time in warning and 46.7% at 30.7% (area under the sensitivity–time-in-warning curve 0.588; Fig. 3C).

**Figure 3:**
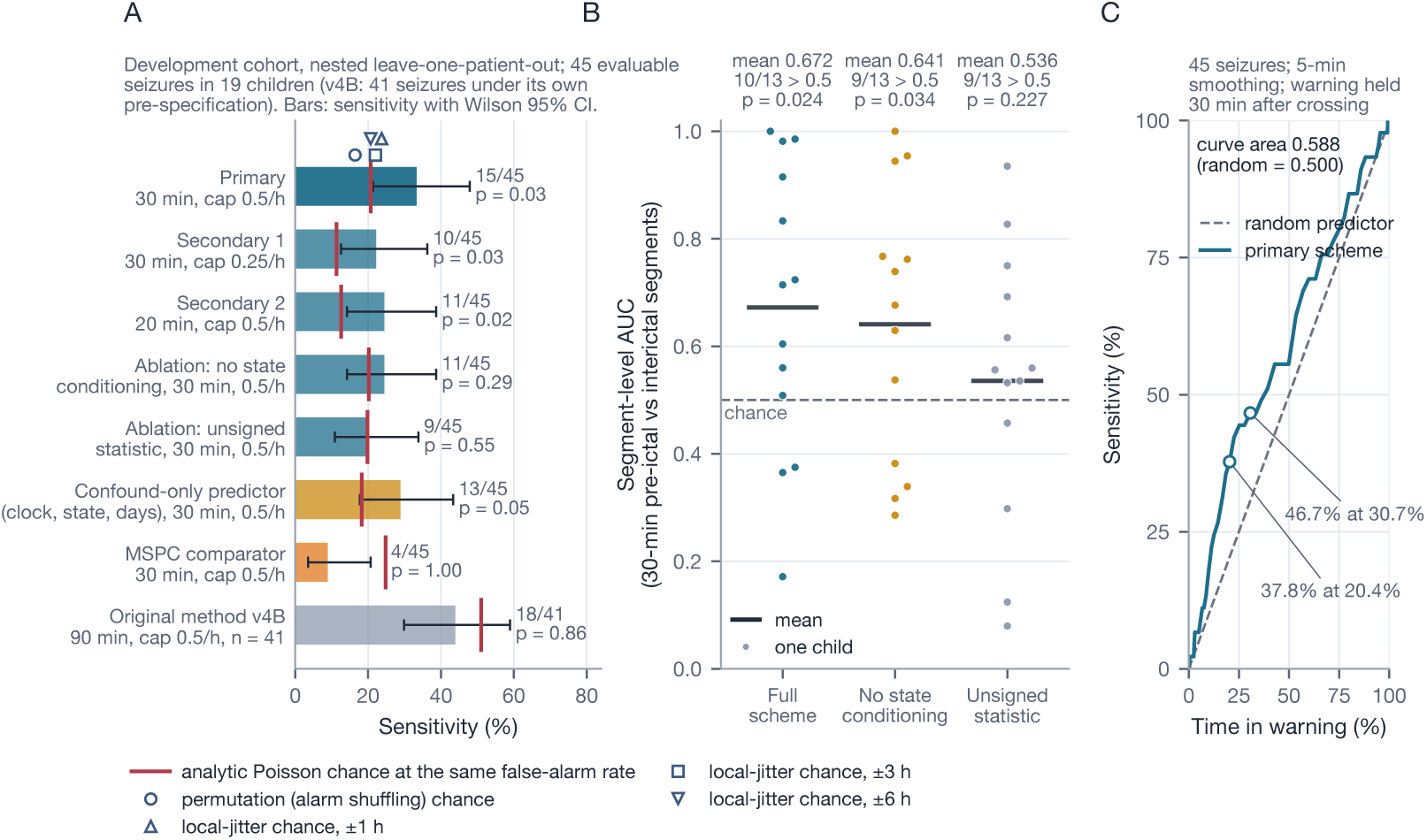
Development-cohort performance of the frozen scheme against chance (nested leave-one-patient-out; 45 evaluable seizures in 19 children). (A) Sensitivity with Wilson 95% CI for the primary operating point, the two secondary points, the two ablations, a confound-only predictor (clock time, estimated vigilance state and days since admission; no EEG waveform feature; sand), the multivariate SPC comparator and the original method (41 seizures, its own pre-specification). Red ticks mark the analytic Poisson chance level at each scheme’s own false-alarm rate, with the binomial p; for the primary point the permutation (alarm-shuffling) and local-jitter (*±*1, *±*3, *±*6 h) chance levels are also shown. (B) Per-patient segment-level AUC (30-min pre-ictal versus interictal segments) for the full scheme and both ablations; dashed, chance (0.5); bars, means; p, one-sided Wilcoxon. (C) Sensitivity versus time in warning for the smoothed primary statistic; dashed diagonal, random predictor.

Secondary operating point 1 (cap 0.25 per hour) detected 10 of 45 seizures (22.2%, 12.5–36.3%) at 0.244 per hour (analytic chance 11.3%, p = 0.026; permutation p = 0.005; *±*1 h jitter p = 0.10, failing that test), and secondary operating point 2 (20-min window, cap 0.5 per hour) detected 11 of 45 (24.4%, 14.2–38.7%) at 0.414 per hour (chance 12.6%, p = 0.021; permutation p = 0.008; *±*1 h jitter p = 0.036). Under strict E4 both fell to 8 of 44 (18.2%) and lost analytic significance (p = 0.17 and 0.22), so neither is robust to a single seizure. The 20 folds selected 13 distinct CUSUM parameter sets, three of them equal to the frozen set; the frozen parameters applied in-sample detect 19 of 45 at 0.470 per hour, an optimistic figure that was not used.

### 3.5. Time course of the pre-ictal signal

The signal was confined to the last 30 min before onset (Fig. 4; Supplementary Note S7). The per-patient segment AUC fell from 0.710 at a 10-min horizon to 0.47 at 4–6 h (Fig. 4A); disjoint rings before onset scored 0.698–0.735 at 0.5–10 min but only 0.50–0.55 from 30 min to 8 h (Fig. 4B); the percentile of z within each child’s own interictal blocks was at chance from 8 h to 30 min before onset and rose only in the last half hour (Fig. 4C); and clock-matched references reduced the 30-min AUC from 0.664 to 0.641 (p = 0.082). Where a pre-ictal change is present at all, it develops within the last half hour, which is what the 30-min window captures.

**Figure 4:**
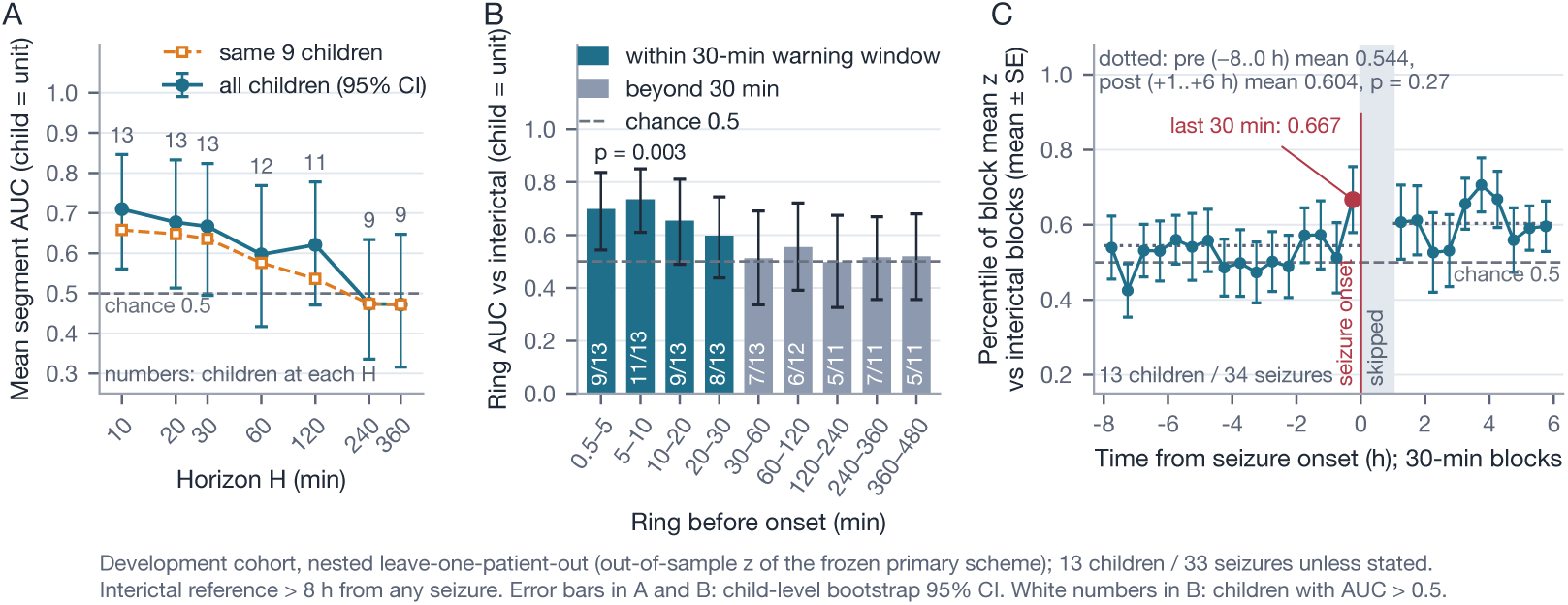
Time course of the pre-ictal signal (development cohort, nested leave-one-patient-out, out-of-sample z of the frozen primary scheme; chance = 0.5, dashed). (A) Per-patient mean AUC of the pre-ictal segment [onset *−* H, onset *−* 0.5 min] versus interictal blocks (> 8 h from any seizure) as a function of horizon H: circles, all available children (number shown), patient-bootstrap 95% CI; squares, the 9 children present at every horizon. (B) AUC of disjoint pre-ictal rings versus equal-length interictal blocks (13 children and 32–33 seizures for rings up to 60 min, 11–12 children and 30–32 seizures beyond; white numbers, children with AUC > 0.5; p, one-sided Wilcoxon). (C) Mean *±* SE percentile of 30-min block means of z within each child’s interictal blocks, *−*8 to +6 h around onset (13 children, 34 seizures); red, last 30 min (0.667); the first post-ictal hour is skipped; dotted, pre- and post-ictal means (0.544 versus 0.604, p = 0.27).

### 3.6. Ablations and comparators

Removing state conditioning reduced sensitivity to 11 of 45 (24.4%) at 0.459 per hour (analytic chance 20.2%, p = 0.29; permutation p = 0.086) and the mean segment AUC to 0.641; replacing the directional statistic by an unsigned Mahalanobis distance gave 9 of 45 (20.0%) at 0.449 per hour (chance 19.8%, p = 0.55; permutation p = 0.50) and a segment AUC of 0.536 (Fig. 3A, B; Table 2). Neither ablation is distinguishable from chance at the alarm level, and the full scheme’s gains over them (four and six seizures of 45) are within sampling error. The ECG-only control reached a mean segment AUC of 0.644 against 0.672 for the frozen EEG features (Supplementary Table S5). Multivariate SPC [24] under our rules detected 4 of 45 seizures (8.9%, 3.5–20.7%) at 0.579 per hour (analytic chance 24.8%, p = 0.998), and under its published protocol 1 of 50 seizures at 0.01 per hour, against the published 88.9% sensitivity at 0.39 per hour on CHB-MIT with patient-specific, segment-wise evaluation. The per-channel criterion of Segal et al. [36] (Supplementary Note S4) was met before 20 of 27 confirmed seizures (74.1%; published: 17 of 19 patients) but also before 104 of 150 seizure-free control epochs (69.3%; one-sided Fisher exact p = 0.40): without seizure-free controls it would have appeared to work.

The confound-only predictor (clock time, sleep–wake estimate and days since admission; no EEG waveform feature), trained leave-one-patient-out with the same labels and scored by the same CUSUM with its own nested parameter selection, detected 13 of 45 seizures (28.9%, 17.7–43.4%) at 0.410 per hour (analytic chance 18.2%, p = 0.055; permutation p = 0.024; *±*1 h jitter p = 0.13), with a mean segment AUC of 0.594 (10 of 13 children above 0.5; p = 0.073) and a sensitivity–time-in-warning curve at least as high as the frozen scheme’s (area 0.627 versus 0.588). Per child, the frozen scheme’s segment AUC exceeded the confound-only predictor’s by 0.078 on average (7 of 13 children; p = 0.19). Tuned in-sample under the same cap, the confound-only predictor reached 16 of 45 at 0.410 per hour and passed all four chance tests (Supplementary Note S6; Fig. 3A).

### 3.7. Per-child results, onset side and what an alarm means

Nine of the 19 children had no detection, six of them with less than 10 h of alarm-eligible interictal time; the per-patient false-alarm rate exceeded the 0.5 per hour cap in 8 of 19 (Fig. 5A); no child was individually distinguishable from chance (Supplementary Tables S3 and S4). With the child as the unit of analysis (post hoc; Supplementary Table S9), the child-cluster bootstrap of the pooled sensitivity gave 33.3% (95% CI 20.0–45.2%; p = 0.031 against the pre-specified chance level of 20.8%). Against each child’s own chance level, which is 18.4% when weighted by seizures, below the pooled level because the children with most evaluable seizures had the lowest false-alarm rates, the pooled difference was +14.9 percentage points (95% CI +3.9 to +25.4; p = 0.004); but the child-averaged sensitivity was 28.9% against a child-averaged chance of 17.0% (difference +11.9; 95% CI *−*0.4 to +24.7; p = 0.029), 9 children lay above and 7 below their own chance level (sign test p = 0.40; Wilcoxon p = 0.060) and no child was significant on its own. The development effect was therefore carried by about half of the children. By reported onset side, 7 of 26 seizures in the 11 children with a left-sided onset were detected (26.9%; chance 20.1%, p = 0.26) and 8 of 19 seizures in the 8 children with a right-sided onset (42.1%; chance 16.1%, p = 0.006); the difference between the sides is within sampling error (Fisher exact p = 0.35).

**Figure 5:**
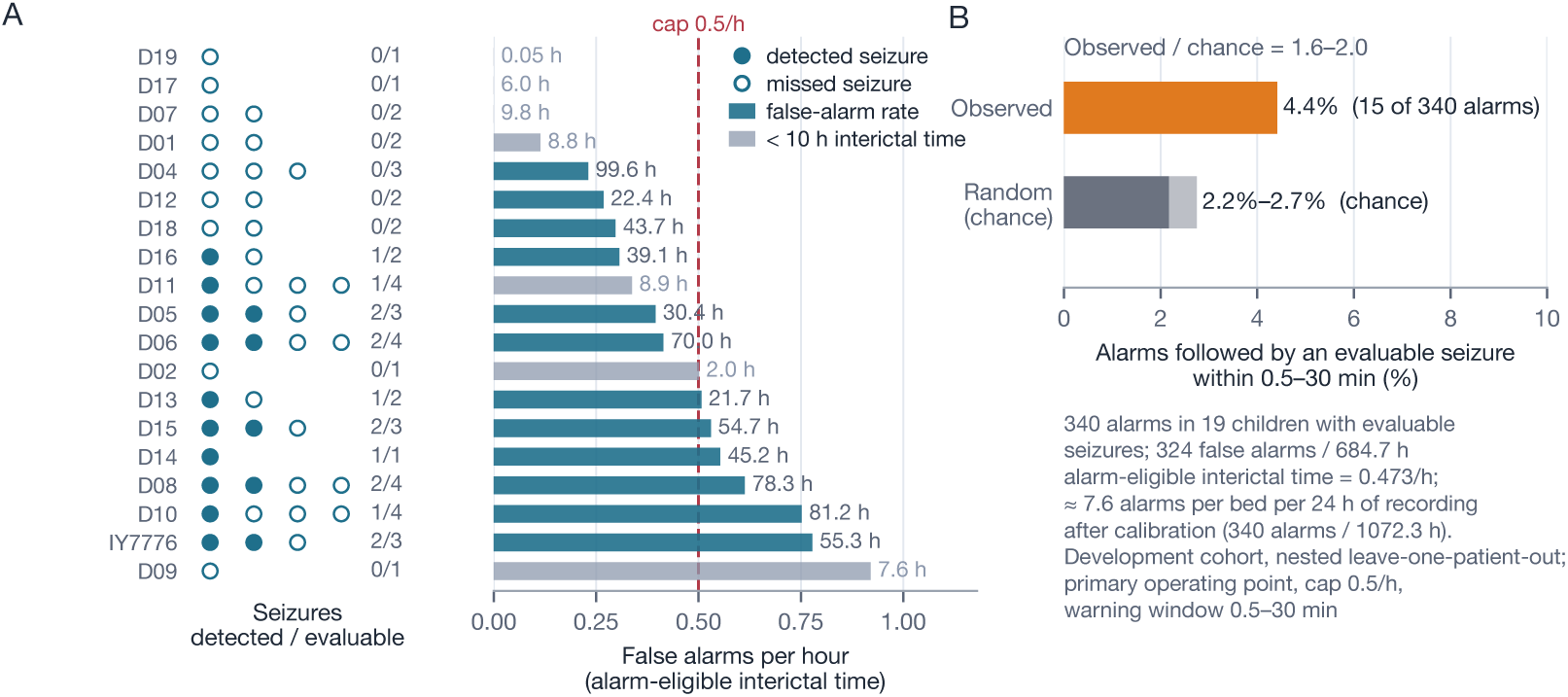
What an alarm means at the primary operating point (development cohort, nested leave-one-patient-out; false-alarm cap 0.5/h, warning window 0.5–30 min). (A) The 19 children with at least one evaluable seizure, sorted by false-alarm rate. Left: evaluable seizures shown as filled (detected) or open (missed) markers with the count. Right: false alarms per hour of alarm-eligible interictal time; dashed, the 0.5/h cap; interictal hours printed at the bar end, grey for children with less than 10 h. (B) Of 340 alarms (15 true, 324 false, 1 neutral), 15 (4.4%) were followed by an evaluable seizure within 0.5–30 min, against 2.2–2.7% expected for randomly placed alarms (surrogate and analytic estimates), i.e. 1.6–2.0 times chance. Pooled: 324 false alarms in 684.7 h = 0.473/h; about 7.6 alarms per bed per 24 h of recording after calibration.

Of the 340 alarms (15 true, 324 false, 1 neutral), 15 (4.4%) were followed by an evaluable seizure within 0.5–30 min, against 2.2–2.7% for as many randomly placed alarms (Fig. 5B). That is about 7.6 alarms per bed per 24 h, and with 1.65 confirmed seizures per monitoring day even a perfect predictor at this false-alarm budget could not exceed about 12% of alarms followed by a seizure.

### 3.8. Locked validation

The frozen scheme was run once on each validation cohort and on both pooled on 17 October 2026 (Supplementary Note S3); 101 seizures were evaluable (51 in cohort A, 50 in cohort B; Table 1).

Pooled, the scheme detected 19 of 101 evaluable seizures (18.8%, 95% CI 12.4–27.5%) within 0.5–30 min of onset at 0.602 false alarms per hour (575 in 955 h; median lead 13.7 min), against an analytic chance level of 25.6% (p = 0.96), a permutation chance level of 17.2% (p = 0.35) and *±*1, *±*3 and *±*6 h jitter chance levels of 17.8%, 19.3% and 19.1% (p = 0.42, 0.62 and 0.59; Fig. 6A; Table 2). The pre-specified primary rule was not met. Cohort A gave 6 of 51 (11.8%, 5.5–23.4%) at 0.598 per hour against 25.5% (p = 0.995) and cohort B 13 of 50 (26.0%, 15.9–39.6%) at 0.610 per hour against 25.9% (p = 0.55; Table 2); in both cohorts the false-alarm rate exceeded the 0.5 per hour cap.

**Figure 6:**
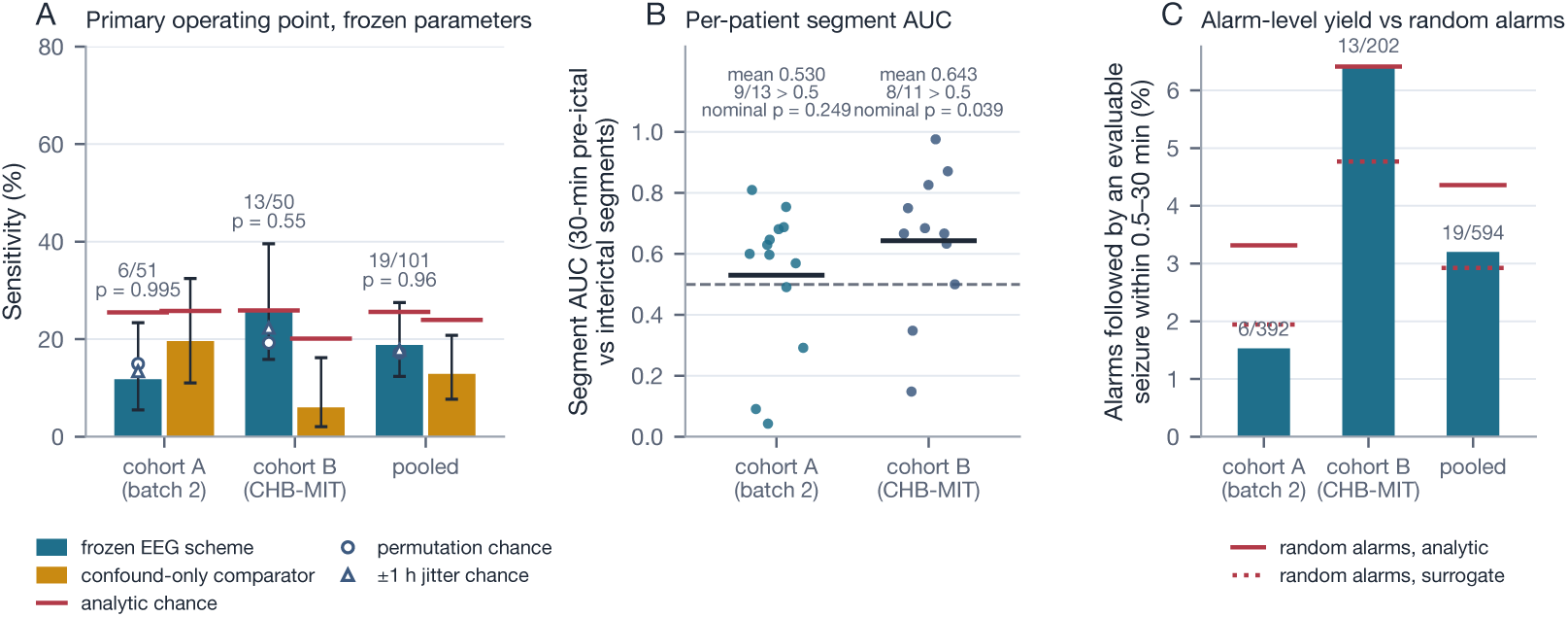
Locked validation (frozen parameters applied once, 17 October 2026). (A) Sensitivity at the primary operating point (0.5–30 min window) with Wilson 95% CI for cohort A (batch 2; 51 evaluable seizures in 19 children), cohort B (CHB-MIT; 50 seizures in 17 recordings) and pooled, for the frozen scheme (blue) and the confound-only comparator (sand); red ticks, analytic Poisson chance at each scheme’s own false-alarm rate, with the analytic p of the frozen scheme; markers, permutation and *±*1 h jitter chance for the frozen scheme. (B) Per-patient segment-level AUC (30-min pre-ictal versus interictal segments) by cohort; bars, means; dashed line, chance; p nominal (Supplementary Note S5). (C) Proportion of alarms followed by an evaluable seizure within 0.5–30 min, observed (bars, with counts) against randomly placed alarms (red: analytic, solid; surrogate, dotted).

No key secondary endpoint was met (pooled; Table 2): the per-patient segment AUC was 0.582 (17 of 24 above 0.5; nominal p = 0.053; Fig. 6B); secondary operating point 1 (cap 0.25 per hour) detected 11 of 101 (10.9%) at 0.344 per hour against 15.5% (p = 0.93); secondary operating point 2 (0.5–20 min window) detected 15 of 101 (14.9%) at 0.498 per hour against 14.9% (p = 0.55; permutation p = 0.065; *±*1 h jitter p = 0.16); the clock-matched segment AUC was 0.584 (11 of 18; p = 0.14); and the confound-only comparator detected 13 of 101 (12.9%) at 0.557 per hour against 23.9% (p = 0.998), six fewer than the frozen scheme. The only nominally significant value among the pre-specified endpoints was the segment AUC of cohort B (0.643; 8 of 11 above 0.5; p = 0.039; clock-matched 0.654, p = 0.15), a test that rejects effect-free series with this time structure a third of the time (Supplementary Note S5); values from the 20-min run that were not pre-specified are in Supplementary Note S7. In cohort A the segment AUC was 0.530 (p = 0.25) and the comparator detected 10 of 51 against the scheme’s 6.

With the child as the unit of analysis (post hoc; Supplementary Table S9), the child-cluster bootstrap interval of the pooled sensitivity was 10.2–27.6% (p = 0.93 against 25.6%); the child-averaged sensitivity was 14.7% against a child-wise chance of 24.6% (difference *−*9.9 percentage points; 95% CI *−*19.3 to *−*0.9); 11 children lay above and 19 below their own chance level, and none was individually above chance. Cohort A alone gave 4 children above and 11 below (bootstrap p = 1.00), cohort B 7 above and 8 below (p = 0.51). By onset side in cohort A, 3 of 30 left-sided seizures (10.0%) and 2 of 18 right-sided seizures (11.1%) were detected, both below their chance levels (20.6% and 17.9%); over the development and same-site validation cohorts together, 10 of 56 left-sided (17.9%) and 10 of 37 right-sided (27.0%) seizures were detected (Fisher exact p = 0.31).

Of the 594 alarms, 19 (3.2%) were followed by an evaluable seizure within 0.5–30 min, against 2.9–4.4% for randomly placed alarms (cohort A 1.5% against 1.9–3.3%; cohort B 6.4% against 4.8–6.4%; Fig. 6C), about 8.1 alarms per bed per 24 h; 14 of 19 children in cohort A and 10 of 17 recordings in cohort B had no detection, and the per-patient false-alarm rate exceeded 0.5 per hour in 10 of the 20 calibrated children of cohort A and in 15 of the 22 calibrated recordings of cohort B (Supplementary Table S7). Under a true sensitivity of 27.1%, half the development effect, 19 or fewer detections of 101 would have occurred with probability 0.035 (post hoc).

## 4. Discussion

### 4.1. Principal findings

A warning scheme that had looked promising on a pilot recording was in-distinguishable from chance under a pre-specified protocol with chance levels matched to its own false-alarm rate, and the reasons are not peculiar to it: its statistic moved three times as much with vigilance state as with seizure approach, its target seizures clustered late in the admissions, and even a state-conditioned statistic drifts with the clock and is autocorrelated for hours. The re-designed scheme beat chance on the development cohort on the primary endpoint and on the segment AUC but not on the clock-matched AUC, and a predictor that uses only clock time, vigilance state and days since admission reproduced most of its performance. In the locked validation the signal did not reproduce: 19 of 101 evaluable seizures at 0.60 false alarms per hour against a chance level of 25.6%, below chance in the same-site cohort and at chance in the public cohort, no secondary endpoint met, and the false-alarm cap not held. The study therefore ends with two pre-specified negative results and a quantified account of why development estimates of scalp-EEG warning are inflated.

### 4.2. What the audit shows, and what evaluations should report

The three confounds are known individually: vigilance-dependent false predictions and the use of separate sleep and wake references [16, 17], circadian and multidien seizure cycles [18, 19, 40], and the clustering of seizures in monitoring units [20]. This study quantifies their joint effect on a scalp warning statistic in a paediatric cohort, as has been done for an intracranial measure [41], and shows, as for diary-based forecasting [14], that the chance model decides the verdict: for the same alarms, uniform permutation gives a chance level of 16%, the analytic Poisson model 21% and *±*1 h jitter 24%, and clock matching left the 30-min segment AUC non-significant. The three confounds persist when the two problems documented by earlier audits are avoided, namely validation on windows drawn at random from temporally grouped labels, which lets a network learn fake seizures [8], and testing that is not patient-independent, as in all but 4% of published work [7]: testing here was patient-independent and alarms were scored on the continuous record, yet these confounds alone reproduced most of the development performance. We suggest that scalp-EEG warning studies report, as a minimum, the analytic chance level at the achieved false-alarm rate [37, 38], a permutation that preserves the alarm structure [13, 42], a local-jitter test that preserves the admission trend and a clock-matched comparison; that the false-positive rate of each test be checked on effect-free simulations with the time structure of the data, as the segment-level tests here rejected a third of the time; and that criteria defined on pre-ictal data alone, such as per-channel pattern thresholds [36], be applied to seizure-free epochs of the same recordings, where in our hands they fired almost as often.

### 4.3. What did not survive, and why

On the development cohort, state conditioning and the sign of the statistic were associated with the above-chance performance and the signal was confined to the last 10–30 min; none of this transferred. Three pre-unblinding analyses had anticipated the outcome. The confound-only predictor reproduced most of the development performance (13 versus 15 of 45; segment AUC 0.594 versus 0.672; paired p = 0.19). The development estimate was the best of eight nested runs, with a crude Šidák bound near 0.24 and p = 0.056 without the pilot child. And effect-free simulations with the cohort’s time structure showed that segment-level tests reject a third of the time, so the decision rested on the alarm-level rule with local jitter; the one nominally significant pre-specified validation value, the segment AUC of cohort B (0.643, p = 0.039), is of exactly that kind. The post hoc child-level analysis sharpens the picture: the development effect was carried by about half of the children and did not reach significance by the sign or the Wilcoxon test, whereas in validation most children fell below their own chance level. The frozen thresholds also failed to hold their false-alarm cap on new patients (0.60 per hour against 0.47 in development), as the autocorrelation of the statistic had led us to expect: empirical thresholds cannot promise a false-alarm rate elsewhere. The comparator failed as well, which shows that the clock, state and admission structure behind the development result is real but that its alignment with seizures is cohort-specific. We never claimed that the development pattern was EEG-specific: a three-feature heart-rate statistic reached similar segment-level separation in screening (mean AUC 0.64 against 0.67; Supplementary Table S5), in line with a single ECG lead carrying as much pre-ictal information as 21 scalp channels [43], and the pattern is compatible with a change of arousal or with a slow, medication-related drift of excitability [44]. Whether any short-horizon change exists in scalp EEG is left open; a plausible one, found with sight of the data, did not survive a locked test.

The absence of signal beyond 30 min contrasts with reports of pre-ictal states lasting hours in intracranial recordings [45] and with the circadian and multi-day structure exploited by implanted and sub-scalp forecasters [5, 46, 47]. Within an admission of a few days with medication withdrawal, a scalp statistic that tracks slow cycles would be hard to separate from the admission trend, which is the third confound.

### 4.4. Clinical meaning

The operating point was chosen with the monitoring unit’s clinical team, for whom an alarm prompts a look at the video rather than an intervention and one false alarm every 2 h was judged tolerable. At that rate the scheme produced roughly eight alarms per bed per day in development and validation; one alarm in twenty-three was followed by a seizure in development, against one in forty for random alarms, and one in thirty-one in validation, within the random range. With these seizure rates even a perfect predictor at the same false-alarm budget could not exceed about one alarm in eight followed by a seizure, because the positive predictive value of any in-unit warning is bounded by the seizure rate and not only by the algorithm [14]. Patients and carers expect prediction and detection devices to combine high sensitivity with few false alarms [48, 49]; the scheme meets neither expectation, and its value is as a reference: an interpretable, frozen scheme with documented chance-corrected performance against which future claims on the same data, including machine-learning ones, can be measured under the same rules, in the way that standardised frameworks serve seizure detection [50, 51].

### 4.5. Strengths and limitations

The strengths of the study are the pre-specification of both evaluations; a hash-recorded freeze and a time-stamped deposit before any validation data were seen; nested cross-validation of the weights and of the parameter selection; four chance models whose false-positive rates were calibrated by simulation; ablations, comparators and a confound-only predictor evaluated under identical rules; a locked validation in two cohorts, one of them public; and clinicians’ electrographic onsets and sleep–wake marks as the gold standard.

The limitations are several. Development-cohort figures are development-set estimates: every design choice was made with sight of these data, and with 45 evaluable seizures a difference of three seizures is within sampling error [52]; four feature sets at two warning windows went through the same nested procedure and the frozen scheme tied for the highest sensitivity, so the development estimate was the best of eight. The pre-specified tests treat seizures as independent although they are nested in children; a design effect of about 1.3 would move the boundary development p-value from 0.034 to about 0.05, and the post hoc child-level analysis gives the same verdict as the pre-specified tests in every cohort while showing how unevenly the development effect was distributed. The cohorts are small for a prediction study, 65 recordings and 3565 h in all, drawn from one tertiary paediatric unit apart from CHB-MIT, with focal epilepsies mostly under presurgical evaluation and medication withdrawal, and no sociodemographic subgroup was examined. The validation had a power of 0.86 for the development effect and 0.39 for half of it, so a half-sized true effect could have been missed, although the observed validation sensitivity lies below chance rather than between chance and the development estimate. The scheme uses no spatial information, so the relation between electrode position and the epileptogenic zone, which is the natural next question for a scalp method, is untested here beyond the post hoc comparison by reported onset side. The pre-specifications are internal documents, written on the day of the freeze and deposited with a time-stamp before unblinding rather than publicly registered, and the primary rule of v2 was amended before its confirmation (13 October 2026) to add the *±*1 h jitter test. The Siegmund approximation used to prune the CUSUM grid omits the overshoot correction; corrected, the nominal false-alarm rate of the frozen thresholds is about 0.02 per hour, more than twenty times below the rates realised, a gap explained by the autocorrelation of interictal z (lag-1 about 0.5) and by non-zero per-child interictal means (*−*0.62 to +0.16). Calibration needs 8 h and therefore excludes short recordings and the first night; quality control discards one window in five; the zero-phase filter is offline; and the alarm-eligible denominator makes the false-alarm rate higher than the number of alarms per hour of recording. The sleep–wake gold standard is the clinicians’ routine review marks, which cover 7727 windows (about 21 h, 1.6% of all windows), not polysomnographic staging. One recording delivered in both batches was recognised as a duplicate only after unblinding and entered no performance figure; validation data should be de-duplicated by file identity before the run. The comparators were re-implemented from their descriptions; not reproducing their published performance under our rules says nothing about the original studies beyond their lack of a chance comparison.

### 4.6. What a conclusive study would need

The negative result is specific to this scheme, these features and these cohorts, and it leaves the existence of a short-horizon scalp-EEG signal open. A study able to settle the question would need several hundred evaluable seizures from more than one centre, so that an effect half the size of our development estimate could be detected with adequate power; a publicly registered pre-specification with the chance models, the operating point and the decision rule fixed in advance; channel-resolved features together with a localisation of each child’s epileptogenic zone, so that the proximity question raised above can be tested rather than assumed; and the same confound audit, because vigilance state, clock time and admission trend will be present in any monitoring-unit data set. The framework and the frozen scheme described here are offered for that purpose.

## 5. Conclusions

In the development cohort, scalp-EEG seizure warning in children under continuous monitoring carried a short and weak above-chance signal of the kind that is easily mistaken for a strong one. In two validation cohorts tested once under locked rules it carried none, and with the child as the unit of analysis no cohort was above chance. Vigilance state, clock time and admission trend inflate apparent performance unless the evaluation is pre-specified and the chance model preserves the structure of the data. The frozen scheme and the evaluation framework around it are offered as a reference rather than as a device: the short-horizon signal did not generalise beyond the cohort on which it was found.

## Data Availability

The hospital recordings contain identifiable video and cannot be shared. Pre-specifications v1 and v2, the frozen configurations and the code manifest were deposited on Zenodo with a time-stamp before unblinding (version 1 of the record, doi:10.5281/zenodo.23439034, created on 17 October 2026). Four files of that archive refer to recordings by the hospital's internal record numbers, so the files of version 1 are under restricted access and are available to editors and reviewers on request. Version 2 of the record is public (doi:10.5281/zenodo.23439033 resolves to the latest version): it is a copy in which these numbers are replaced by a fixed string and nothing else is changed, with a script that verifies the copy against the original (Supplementary Note S3). De-identified per-window feature arrays, the per-seizure and per-patient outputs underlying every figure and table and the evaluation code will be added to the same record on acceptance. The CHB-MIT Scalp EEG Database is publicly available from PhysioNet.

## Ethical statement

The study was conducted in accordance with the principles embodied in the Declaration of Helsinki and in accordance with local statutory requirements. It was approved by the Ethics Committee of Peking University First Hospital (2021-066), which waived the requirement for informed consent for this retrospective analysis of routine recordings. No names, photographs, exact ages or dates of individual children are reported.

## Data availability statement

The hospital recordings contain identifiable video and cannot be shared. Pre-specifications v1 and v2, the frozen configurations and the code manifest were deposited on Zenodo with a time-stamp before unblinding (version 1 of the record, doi:10.5281/zenodo.23439034, created on 17 October 2026). Four files of that archive refer to recordings by the hospital’s internal record numbers, so the files of version 1 are under restricted access and are available to editors and reviewers on request. Version 2 of the record is public (doi:10.5281/zenodo.23439033 resolves to the latest version): it is a copy in which these numbers are replaced by a fixed string and nothing else is changed, with a script that verifies the copy against the original (Supplementary Note S3). De-identified per-window feature arrays, the per-seizure and per-patient outputs underlying every figure and table and the evaluation code will be added to the same record on acceptance. The CHB-MIT Scalp EEG Database is publicly available from PhysioNet [29, 30].

## Supplementary material

Supplementary Methods S1–S3, Tables S1–S9, Notes S1–S7 and Appendices A and B (the pre-specifications in English translation) are provided as a separate file.

## Author contributions

**Mufeng Chen:** Conceptualization, Methodology, Software, Validation, Formal analysis, Investigation, Data curation, Writing – original draft, Visualization, Project administration. **Guojing Yu:** Investigation, Data curation, Resources, Project administration, Writing – review & editing. **Xiwen Lai:** Visualization, Writing – review & editing. **Pan Gong:** Resources, Investigation, Writing – review & editing. **Shuang Wang:** Resources, Investigation, Writing – review & editing. **Xiaoyan Liu:** Resources, Investigation, Writing – review & editing. **Qi Wu:** Visualization, Writing – review & editing. **Yiming Zhang:** Investigation. **Bomiao Sun:** Investigation. **Shenglin Xu:** Investigation, Conceptualization. **Jiayi Yao:** Conceptualization. **Hui Zhang:** Conceptualization. **Hongrui Zhang:** Conceptualization, Methodology, Investigation. **Zeyu Wu:** Conceptualization. **Jingtong Li:** Investigation. **Quansheng Ren:** Conceptualization, Methodology, Supervision, Writing – review & editing. **Taoyun Ji:** Conceptualization, Resources, Supervision, Project administration, Funding acquisition, Writing – review & editing. All authors reviewed the manuscript and approved its submission.

## Acknowledgements

The High Performance Computing Platform of Peking University provided computational resources for data analysis and evaluation.

## Use of generative AI

During the preparation of this work the authors used Claude (Anthropic) in order to assist with grammar and language checking. After using this tool, the authors reviewed and edited the content as needed and take full responsibility for the content of the published article.

