## supplementary materials for "Scalp EEG seizure warning in children under a pre-specified, locked evaluation: confound audit, structured chance levels and a negative result"

Contents: Supplementary Methods S1 (original method and pre-specification v1), S2 (the frozen scheme in full) and S3 (validation cohort adapters); Supplementary Tables S1 (frozen parameters), S2 (quality control and vigilance estimator), S3–S4 (per-patient and per-seizure development results), S5 (feature-set screening and the ECG-only control), S6 (code manifest), S7 (per-patient validation results), S8 (clinical characteristics from the monitoring reports) and S9 (post hoc analyses added at revision: the child as the unit of analysis, and reported onset side); Supplementary Notes S1 (chance tests and power), S2 (vigilance tabulation of the original method), S3 (pre-specifications), S4 (comparators under their own protocols), S5 (simulation calibration of the chance tests), S6 (confound-only predictor) and S7 (horizon analyses in full); Appendices A and B (pre-specifications v1 and v2, English translations of the deposited originals).

#### Supplementary Methods S1. The original method (version 4B) and pre-specification v1

The original method operated on the 19 referential channels of the unfiltered 500 Hz recording in non-overlapping 10-s windows. Two features per channel — spectral entropy over 0.5–70 Hz and Hjorth mobility — were expressed as a squared Mahalanobis distance from the channel's recent baseline (Ledoit–Wolf shrinkage covariance). Each channel's distance was mapped to a standard-normal score by a probability-integral transform against the baseline queue, and the 19 scores were fused with non-negative weights derived from the baseline covariance. The baseline was a sliding queue of the most recent 2880 clean windows (8 h) refitted every 1440 admitted windows; after the first confirmed seizure of the monitoring period the queue was shortened irreversibly to 720 windows (2 h), refitted every 360 windows. A one-sided CUSUM of the fused score raised an alarm when its risk transform exceeded the decision limit h_eff for N consecutive windows; alarms and seizures were followed by a 1-h lock-out. The warning window was 0.5–90 min.

Pre-specification v1 fixed, before the cohort analysis: the gold standard (clinician-annotated electrographic onset of confirmed, spontaneous seizures), the admissible data, the calibration rule, the evaluable-seizure criteria E1–E5 (with E3 requiring at least 90 min after calibration and E4 at least 2.5 h after the preceding seizure, i.e. the 1-h lock-out plus the 90-min window), the alarm scoring, and two chance tests (analytic Poisson with an 89.5-min window; uniform permutation of each patient's alarms, pre-specified at 1000 permutations, of which 2000 were run). CUSUM parameters were selected per held-out patient on the other patients from a grid of 10 × 14 × 5 = 700 combinations ($k \in \{0.3, 0.4, 0.5, 0.6, 0.7, 0.8, 0.9, 1, 1.1, 1.2\}$; h_eff from 2 to 40, geometric; $N \in \{1, 2, 3, 5, 8\}$), after pruning combinations whose Siegmund average run length implied more than 0.5 false alarms per hour, under the constraint that the pooled false-alarm rate of the training patients did not exceed 0.5/h. Results: Table 2 of the main text and Supplementary Note S1. The full document is given in English translation as Supplementary Appendix A.

### Supplementary Methods S2. The frozen scheme in full

The frozen scheme (v5p; Fig. 1B of the main text; every constant in Supplementary Table S1) keeps the SPC framework and changes the signal layer, the baseline, the statistic and the warning window.

*Signal and quality control.* Referential recordings are re-referenced to the 18-channel longitudinal bipolar montage by subtraction, which cancels the common reference; native bipolar recordings are used directly. Data are resampled to 256 Hz, notch-filtered at 50 and 60 Hz, band-pass filtered at 0.5–45 Hz (fourth-order Butterworth, zero-phase, an offline implementation; all later layers use only past data) and cut into non-overlapping 10-s windows. A window is invalid if any channel fails any of seven rules (six amplitude, variance, slow-wave-share or high-frequency thresholds and a missing-channel rule; Supplementary Tables S1 and S2), whose thresholds were calibrated against the original method's quality control using signal-quality measures only (473 565 windows; 81.6% valid versus 80.7%; window-wise agreement 86.0%).

*Features.* Per window and channel, 11 spectral and time-domain quantities are computed, together with 29 synchrony quantities from the 18 × 18 zero-lag correlation matrix and a scalp high-frequency index (median log 65–95 Hz variance) that replaces the EMG channel. Seven global features are monitored: spectral entropy, Hjorth complexity, Hjorth mobility, δ and β log power averaged over channels, the mean absolute inter-channel correlation and the first eigenvalue fraction of the correlation matrix.

*Vigilance state.* An EEG-only logistic estimator classifies each window as sleep or wake from 13 global features standardised by the median and median absolute deviation of the current baseline queue, with a causal five-window median and a 0.5 threshold. Its coefficients were trained on the development cohort against clinicians' sleep and wake marks (7727 labelled windows in 21 children). In leave-one-patient-out evaluation the offline estimator had a pooled AUC of 0.964 and an accuracy of 91.9%; the causal run-time version agreed with the marks in 86.9% of the 7595 labelled windows of the 20 calibrated children (in-sample; Supplementary Table S2).

*State-conditional rolling baseline.* Before each block, the queue of the most recent 8640 admitted windows (24 h of clean data: valid, outside provocation periods and outside the hour after a confirmed seizure or a recording gap; pre-ictal windows are admitted) is split by estimated state, and the mean and SD of each monitored feature are computed separately for sleep and for wake; each window is standardised with the statistics of its own state. The baseline is refitted after every 360 newly admitted windows using only data preceding the block; the first 1440 admitted windows are bootstrapped with their own statistics inside the calibration period; a window whose state has fewer than 300 windows in the queue is not monitored. The 24-h queue is a cap: it was reached in 575 of 906 blocks and never in 8 of 20 children.

*Directional statistic and probability-integral transform.* The seven standardised features x (missing values set to 0, clipped to ±5) are combined into $y = w \cdot x$ with seven frozen weights (Supplementary Table S1) so that y increases in the pre-ictal direction: positive for spectral entropy, β power, δ power and the first eigenvalue fraction, negative for Hjorth mobility, Hjorth complexity and the mean absolute correlation. The weights are those of an L2 logistic regression (C = 0.05; C and the 15-min training span were chosen on the development cohort) fitted on the 20 calibrated development children, with windows 15–0.5 min before each lead seizure as positives (54 seizures with baseline coverage; 3981 windows) and windows more than 4 h from any seizure as negatives (at most 4000 per child; 53 263 windows), each child weighted equally within its class. Because the mean absolute correlation and the first eigenvalue fraction are highly collinear, individual weights should not be read physiologically; their net effect is that a fall in inter-channel synchrony raises y. Then $z = \Phi^{-1}(\hat{F}(y))$, with $\hat{F}$ the Blom-corrected empirical distribution (Blom 1958) of y over the same-state windows of the current queue (447–6785 windows in the development cohort), so that interictal z is approximately standard normal within each state (its out-of-sample interictal mean is about −0.2 and it retains a clock-time profile; Fig. 2B) and one set of alarm parameters can be applied to all patients.

*Alarm.* A one-sided CUSUM $S_t = \max(0, S_{t-1} + z_t - k)$ raises an alarm when S exceeds the decision limit $h_{eff}$ for N consecutive valid windows; S is then reset and alarms are locked out for the length of the warning window. S is also reset at each confirmed seizure (60-min lock-out) and at recording gaps (> 60 s), and held across invalid or unmonitored windows while the consecutive count restarts. The primary operating point (0.5–30 min window, false-alarm cap 0.5/h) uses k = 0.5, $h_{eff}$ = 7.971 and N = 8 (80 s); secondary point 1 (cap 0.25/h) k = 0.75, $h_{eff}$ = 7.971 and N = 8; secondary point 2 (0.5–20 min window, cap 0.5/h) k

= 0.25, h_eff = 40.0 and N = 5. Each set was chosen on the pooled out-of-sample z series of the development children from 420 combinations pruned by the Siegmund average run length, as the combination with the highest pooled sensitivity under the cap (ties broken by the lower false-alarm rate). Only three quantities were learned from the development cohort: the state coefficients, the seven weights and the CUSUM parameters.

**Resampling check.** Hospital recordings (500 Hz) are resampled to 256 Hz with a polyphase FIR (scipy resample_poly, 64/125, Kaiser β = 5, 2501 taps, zero-phase). Its passband ripple is ≤ 0.016 dB over 0.5–100 Hz, −3 dB at 123 Hz, −61 dB at 150 Hz, with a first stopband sidelobe of −53.5 dB at 152 Hz and ≥ 60 dB attenuation above 175 Hz; content at 128–150 Hz folds back onto 106–128 Hz, outside every feature band, and the sidelobe region 152–172 Hz folds onto 84–104 Hz with at least 53 dB attenuation. On 12 interictal segments of 10 min (4 children, 18 bipolar channels) the power spectral density at 256 Hz differed from that at 500 Hz by at most +0.011 dB in any band below 110 Hz; energy above 128 Hz was −29 dB relative to 0.5–45 Hz and its predicted aliased contribution below −65 dB in every feature band; per-window features computed at both rates agreed with r ≥ 0.999 (band powers, entropy, line length, high-frequency index) and r ≥ 0.988 (quality-control peak-to-peak ratio), and a control with an 8th-order 100-Hz low-pass applied before resampling changed no feature below 45 Hz and shifted the 65–95 Hz index by −0.028 log10, exactly the low-pass's own passband attenuation at 85–95 Hz (predicted −0.029). An independent re-derivation reproduced all values to within 0.01 dB. Resampling therefore neither broadens the spectrum nor aliases measurable energy into the features; spectral resolution is set by the 4-s Welch window (0.25 Hz) at both rates. Script: scripts/60_paper/exp_resample_check.py; outputs: data/processed/_cohort/dev_p/resample_check/.

### Supplementary Table S1. Frozen configuration of the scheme (version 5p)

Frozen at 2026-09-29 18:54 UTC; SHA-256 of the canonical JSON without the hash field: `6f06a97220796481aba6b75f941a4b20f54bd2a0ef1b7eb1e561896801f341a1`. The second file (20-min window, secondary operating point 2) has SHA-256 `c246ab568323edeb49218e805be7651539271be44b36384ee38edef81780b336`. The runner refuses to execute if the digest does not match. Values marked * are descriptive copies of constants fixed in code (verified identical).

| Item | Value |
|---|---|
| Montage | 18-channel longitudinal bipolar: Fp1-F3, F3-C3, C3-P3, P3-O1, Fp2-F4, F4-C4, C4-P4, P4-O2, Fp1-F7, F7-T3, T3-T5, T5-O1, Fp2-F8, F8-T4, T4-T6, T6-O2, Fz-Cz, Cz-Pz |
| Sampling rate | 256 Hz (resampled) |
| Window | 10 s, non-overlapping |
| Notch* | [50, 60] Hz, Q = 30 |
| Band-pass* | [0.5, 45] Hz, Butterworth order 4, zero-phase |
| QC: flat / dropped channel* | variance < 0.5 µV² |
| QC: extreme amplitude* | peak-to-peak > 3000 µV |
| QC: high amplitude not slow-wave* | peak-to-peak > 800 µV and δ fraction < 0.4 |
| QC: step / square wave* | peak-to-peak ÷ SD > 20 (0.5–45 Hz) |
| QC: burst EMG / movement* | peak-to-peak ÷ SD > 14 (0.5–100 Hz) |
| QC: EMG-dominated* | log10(65–95 Hz variance ÷ 0.5–45 Hz variance) > 0 |
| QC: missing channel* | any channel with missing samples in the window |
| QC rule | any channel failing any rule invalidates the window |
| State features (in coefficient order) | δ/β power ratio; θ/β power ratio; α/θ power ratio; relative γ power; relative δ power; total log power; spectral entropy; Hjorth mobility; Hjorth complexity; occipital minus frontal α; central β/δ ratio; first eigenvalue fraction; scalp high-frequency index (65–95 Hz) |
| State coefficients | -0.5955, -0.4554, -0.0702, -4.2491, +0.6305, -0.9601, +0.0497, -0.7618, -0.2936, -0.7044, -0.0221, -0.2332, -1.0503; intercept -0.5651 |
| State standardisation | median / (1.4826·MAD) of the baseline queue, clipped to ±8 |
| State smoothing / | causal median of last 5 valid windows; sleep if probability ≥ 0.5 |

| Item | Value |
|---|---|
| threshold | |
| Baseline queue | 8640 admitted windows (24 h clean); refit every 360; bootstrap 1440; minimum 300 windows per state |
| Admission to queue | valid & not provocation & not within 1 h after confirmed seizure & not within 1 h after gap |
| Monitored features (in weight order) | spectral entropy; Hjorth complexity; Hjorth mobility; δ log power; β log power; mean absolute inter-channel correlation; first eigenvalue fraction |
| Weights | +0.4758, -0.0234, -0.4291, +0.1010, +0.3124, -0.7275, +0.4184 (logistic regression, C=0.05, pre-ictal training span 15 min, 20 cases) |
| Clip* | ±5 |
| Probability-integral transform* | Blom ECDF of same-state baseline queue, >=100 windows, u clipped to [1e-6, 1-1e-6] — in practice z is bounded by the Blom extreme quantile of the same-state queue (about ±3.0 to ±3.8 for queues of 447–6785 windows); the 1e-6 clip never acts |
| Warning window | 3 windows (0.5 min) to 30 min before onset |
| Lock-out | 30 min after an alarm; 60 min after a confirmed seizure* |
| Lead for evaluability (E4) | 1.5 h |
| CUSUM, false-alarm cap 0.5/h | k = 0.5, h_eff = 7.971, N = 8; Siegmund ARL0 = 5774 windows (uncorrected approximation $(\exp(2kh) - 2kh - 1)/(2k^2)$; with the +1.166 overshoot correction 18 565 windows, ≈ 0.019/h); in-sample development result 19/45 at 0.470/h (optimistic; not used) |
| CUSUM, false-alarm cap 0.25/h | k = 0.75, h_eff = 7.971, N = 8; Siegmund ARL0 = 138501 windows (uncorrected approximation $(\exp(2kh) - 2kh - 1)/(2k^2)$; with the +1.166 overshoot correction 796 276 windows, < 0.001/h); in-sample development result 11/45 at 0.246/h (optimistic; not used) |
| CUSUM, 20-min window, cap 0.5/h (secondary 2) | k = 0.25, h_eff = 40.0, N = 5; lock 20 min after alarm |

### Supplementary Table S2. Quality-control calibration and vigilance-state estimator

| Item | Value | Source |
|---|---|---|
| Windows in development cohort | 473 565 | frozen quality control over 21 recordings |
| Valid under frozen QC / under original QC | 81.6% / 80.7% | calibration of the six thresholds (plus the missing-channel rule) on signal-quality measures only |
| Window-wise agreement of the two QC schemes | 86.0% | same |
| State estimator training labels | 7727 windows labelled from clinicians' sleep/wake marks (mark window + next two), 21 children | pre-specification v2 §1.3 |
| State logistic regression | C = 1.0, balanced class weights; labels = mark window plus the next two windows | dev_state_model.py |
| Offline estimator, leave-one-patient-out | pooled held-out AUC 0.964; accuracy 91.9% (label-weighted; per-patient values in state_model_cv_p.csv) | pre-specification v2 §1.3; dev_state_model.py |
| Causal run-time estimator vs clinicians' marks (in-sample) | 86.9% of the 7595 labelled windows of the 20 calibrated children with a run-time estimate | verification of 2 October 2026 |
| Frequency bands | δ 0.5–4, θ 4–8, α 8–13, β 13–30, γ 30–45 Hz; spectral entropy of the normalised 0.5–45 Hz spectrum | v5p_core.py |
| Lead seizure (weights training) | confirmed, spontaneous seizure at least 1.5 h after the preceding confirmed seizure; positives = windows 15–0.5 min before onset | pre-specification v2 §1.5; dev_v5.py |
| Queue cap reached | 575 of 906 baseline blocks; never reached in 8 of 20 calibrated children | same |
| Same-state reference windows for the PIT | 447–6785 per block | same |

**Supplementary Table S3. Per-patient results at the primary operating point (development cohort, nested leave-one-patient-out).** Segment AUC requires at least one pre-ictal and five interictal segments. Fold parameters are the CUSUM parameters selected on the other 19 patients.

| Study code | Evaluable seizures | Detected | False alarms | Alarm-eligible interictal time (h) | False alarms per hour | Lead time(s), min | Segment AUC | Pre-ictal / interictal segments | Fold parameters k / h_eff / N |
|---|---|---|---|---|---|---|---|---|---|
| D01 | 2 | 0 | 1 | 8.8 | 0.114 | — | — | 2 / 3 | 0.5 / 7.971 / 8 |
| D02 | 1 | 0 | 1 | 2.0 | 0.501 | — | — | 1 / 0 | 0.25 / 31.767 / 5 |
| D03 | 0 | 0 | 0 | 0.0 | — | — | — | 0 / 0 | 0.5 / 10.037 / 8 |
| D04 | 3 | 0 | 23 | 99.6 | 0.231 | — | 0.604 | 2 / 235 | 0.5 / 10.037 / 8 |
| D05 | 3 | 2 | 12 | 30.4 | 0.395 | 9.00, 20.17 | 0.915 | 3 / 51 | 0.5 / 12.638 / 5 |
| D06 | 4 | 2 | 29 | 70.0 | 0.414 | 24.50, 13.33 | 0.714 | 4 / 140 | 0.5 / 10.037 / 8 |
| D07 | 2 | 0 | 0 | 9.8 | 0.000 | — | — | 2 / 2 | 0.75 / 5.027 / 5 |
| D08 | 4 | 2 | 48 | 78.3 | 0.613 | 2.83, 16.83 | 0.724 | 4 / 172 | 0.5 / 12.638 / 1 |
| D09 | 1 | 0 | 7 | 7.6 | 0.920 | — | 1.000 | 1 / 13 | 0.75 / 5.027 / 3 |
| D10 | 4 | 1 | 61 | 81.2 | 0.752 | 5.33 | 0.375 | 4 / 198 | 0.75 / 5.027 / 2 |
| D11 | 4 | 1 | 3 | 8.9 | 0.337 | 27.33 | — | 4 / 1 | 0.5 / 12.638 / 3 |
| D12 | 2 | 0 | 6 | 22.4 | 0.268 | — | 0.171 | 2 / 35 | 0.5 / 12.638 / 5 |
| D13 | 2 | 1 | 11 | 21.7 | 0.507 | 14.67 | 0.833 | 2 / 42 | 0.5 / 10.037 / 5 |
| D14 | 1 | 1 | 25 | 45.2 | 0.553 | 23.33 | 0.981 | 1 / 108 | 0.5 / 7.971 / 8 |
| D15 | 3 | 2 | 29 | 54.7 | 0.530 | 11.67, 10.17 | 0.509 | 3 / 116 | 0.5 / 12.638 / 3 |
| D16 | 2 | 1 | 12 | 39.1 | 0.307 | 20.50 | 0.986 | 2 / 69 | 0.5 / 15.913 / 1 |
| D17 | 1 | 0 | 0 | 6.0 | 0.000 | — | — | 1 / 4 | 0.25 / 25.229 / 8 |
| D18 | 2 | 0 | 13 | 43.7 | 0.297 | — | 0.365 | 2 / 89 | 0.75 / 6.33 / 2 |
| D19 | 1 | 0 | 0 | 0.1 | 0.000 | — | — | 1 / 0 | 0.75 / 6.33 / 2 |
| IY7776 | 3 | 2 | 43 | 55.3 | 0.778 | 11.50, 23.83 | 0.560 | 3 / 125 | 0.5 / 7.971 / 8 |

Per-patient nominal segment-level tests (exact one-sided Mann–Whitney, Benjamini–Hochberg q across children): q < 0.05 for two children (D16, q = 0.021; D05, q = 0.032) when interictal segments are treated as independent; the lag-1 correlation of adjacent interictal segments is mostly 0.4–0.8 (range −0.13 to 0.77 in the 13 children with segment AUCs), and tests preserving this autocorrelation (circular shifts of seizure times; thinned references) leave no child with q < 0.05.

**Supplementary Table S4. The 45 evaluable seizures of the development cohort at the primary operating point.** Lead is the time from the earliest alarm inside the 0.5–30 min window to onset. The percentile is the share of the patient's interictal 30-min segments scoring below the pre-ictal segment (50% = indistinguishable).

| Study code | Seizure | Onset (h from start) | Clock time | Detected | Lead (min) | Last alarm before onset (min) | Pre-ictal segment score (z) | Percentile within own interictal segments (%) | Valid windows in warning window (%) |
|---|---|---|---|---|---|---|---|---|---|
| D01 | 1 | 15.4 | day 2, 00:41 | no | — | — | -0.47 | — | 79 |
| D01 | 2 | 23.9 | day 2, 09:10 | no | — | 275.0 | 0.17 | — | 72 |
| D02 | 1 | 17.7 | day 2, 03:50 | no | — | 172.8 | -0.69 | — | 79 |
| D04 | 1 | 142.6 | day 7, 10:21 | no | — | 627.5 | -0.25 | 40 | 76 |
| D04 | 2 | 145.5 | day 7, 13:12 | no | — | 105.8 | 0.20 | 81 | 37 |
| D04 | 3 | 150.2 | day 7, 17:56 | no | — | 58.3 | — | — | 20 |
| D05 | 1 | 29.7 | day 2, 14:42 | no | — | 44.8 | 0.56 | 94 | 89 |
| D05 | 2 | 34.0 | day 2, 19:00 | yes | 9.0 | 9.0 | 0.51 | 92 | 88 |
| D05 | 3 | 50.4 | day 3, 11:24 | yes | 20.2 | 20.2 | 0.32 | 88 | 88 |
| D06 | 1 | 64.1 | day 4, 05:58 | no | — | 310.8 | -0.31 | 36 | 85 |
| D06 | 2 | 89.3 | day 5, 07:12 | yes | 24.5 | 24.5 | 0.81 | 96 | 95 |
| D06 | 3 | 106.7 | day 6, 00:34 | yes | 13.3 | 13.3 | 0.81 | 96 | 96 |
| D06 | 4 | 111.9 | day 6, 05:48 | no | — | 53.5 | -0.00 | 57 | 86 |
| D07 | 1 | 14.8 | day 1, 23:03 | no | — | — | -0.40 | — | 89 |
| D07 | 2 | 21.1 | day 2, 05:18 | no | — | — | -0.66 | — | 97 |
| D08 | 1 | 30.8 | day 2, 16:43 | yes | 2.8 | 2.8 | 0.55 | 84 | 89 |
| D08 | 2 | 95.6 | day 5, 09:32 | no | — | 704.3 | 0.23 | 69 | 78 |
| D08 | 3 | 101.7 | day 5, 15:36 | no | — | 81.7 | 0.03 | 55 | 84 |
| D08 | 4 | 121.8 | day 6, 11:47 | yes | 16.8 | 16.8 | 0.51 | 83 | 92 |
| D09 | 1 | 12.7 | day 1, 21:44 | no | — | 32.0 | 1.02 | 100 | 96 |
| D10 | 1 | 32.8 | day 2, 19:12 | no | — | 52.5 | 0.34 | 73 | 36 |
| D10 | 2 | 132.8 | day 6, 23:13 | no | — | 131.2 | -1.66 | 1 | 93 |
| D10 | 3 | 158.2 | day 8, 00:40 | yes | 5.3 | 5.3 | 0.37 | 76 | 66 |

| Study code | Seizure | Onset (h from start) | Clock time | Detected | Lead (min) | Last alarm before onset (min) | Pre-ictal segment score (z) | Percentile within own interictal segments (%) | Valid windows in warning window (%) |
|---|---|---|---|---|---|---|---|---|---|
| D10 | 4 | 160.3 | day 8, 02:42 | no | — | 127.8 | -1.60 | 1 | 100 |
| D11 | 1 | 11.8 | day 1, 21:20 | no | — | 70.3 | 0.50 | — | 99 |
| D11 | 2 | 20.4 | day 2, 06:00 | no | — | 240.5 | -1.46 | — | 93 |
| D11 | 3 | 22.1 | day 2, 07:42 | no | — | 342.3 | -0.78 | — | 93 |
| D11 | 4 | 23.6 | day 2, 09:12 | yes | 27.3 | 27.3 | 0.33 | — | 99 |
| D12 | 1 | 18.6 | day 2, 04:50 | no | — | — | -0.63 | 17 | 98 |
| D12 | 2 | 39.9 | day 3, 02:08 | no | — | 244.3 | -0.56 | 17 | 92 |
| D13 | 1 | 26.4 | day 2, 14:24 | yes | 14.7 | 14.7 | 0.68 | 90 | 99 |
| D13 | 2 | 29.5 | day 2, 17:29 | no | — | 77.0 | -0.04 | 76 | 83 |
| D14 | 1 | 28.9 | day 2, 14:31 | yes | 23.3 | 23.3 | 1.56 | 98 | 98 |
| D15 | 1 | 40.0 | day 3, 00:47 | yes | 11.7 | 11.7 | 0.50 | 78 | 98 |
| D15 | 2 | 88.1 | day 5, 00:57 | yes | 10.2 | 10.2 | -0.14 | 42 | 93 |
| D15 | 3 | 94.0 | day 5, 06:50 | no | — | 363.5 | -0.35 | 33 | 96 |
| D16 | 1 | 45.6 | day 3, 11:40 | yes | 20.5 | 20.5 | 0.77 | 100 | 80 |
| D16 | 2 | 53.2 | day 3, 19:13 | no | — | 142.2 | 0.22 | 97 | 100 |
| D17 | 1 | 20.3 | day 2, 05:12 | no | — | — | -0.82 | — | 91 |
| D18 | 1 | 11.3 | day 1, 20:25 | no | — | — | -1.12 | 7 | 47 |
| D18 | 2 | 23.1 | day 2, 08:13 | no | — | — | -0.09 | 66 | 98 |
| D19 | 1 | 21.7 | day 2, 06:41 | no | — | — | -0.30 | — | 97 |
| IY7776 | 1 | 28.1 | day 2, 13:08 | yes | 11.5 | 11.5 | 0.35 | 69 | 94 |
| IY7776 | 2 | 47.1 | day 3, 08:05 | no | — | 175.2 | -0.38 | 16 | 74 |
| IY7776 | 3 | 75.6 | day 4, 12:35 | yes | 23.8 | 23.8 | 0.66 | 83 | 86 |

### Supplementary Methods S3. Validation cohort adapters

**Cohort B (CHB-MIT).** Cohort B is the public CHB-MIT Scalp EEG Database of paediatric recordings from Boston Children's Hospital, native bipolar at 256 Hz with annotated seizure onsets: 24 recordings from 23 children (chb21 is the same child as chb01 recorded 1.5 years later; both are kept as separate recordings), 686 files, 983 h and 198 annotated seizures. The adapter was frozen on 14 October 2026 from the EDF headers, the annotation files and the frozen quality control alone, before any statistic or alarm was computed: it maps the 10–10 electrode names onto the montage, derives the montage by subtraction for the three common-reference files of chb12, orders files by the header clock (recording gaps > 60 s, as in cohort A) and takes every annotated seizure as confirmed and spontaneous. Of the windows, 75.9% passed quality control; 22 recordings completed calibration and 50 seizures in 17 recordings met the evaluability criteria. Our group had used CHB-MIT earlier in 2026 for an unrelated deep-learning project (cross-patient seizure classification); no constant of the frozen scheme derives from it and it was not processed with the present pipeline before the freeze: it is unused by this scheme rather than unseen by the authors. Electrode names follow the 10–10 convention (T7/T8/P7/P8 for T3/T4/T5/T6; upper-case FP1, FZ, CZ, PZ) and are mapped onto the frozen montage; duplicated channel labels take the first occurrence; three files of chb12 are common-reference recordings (CS2 or an unspecified reference, with O1 labelled '01') from which the montage is derived by subtraction, the reference cancelling; recordings are ordered by the EDF header clock (the header date is a surrogate, the time of day is real and agreed with the annotation files in every file that gives a time); gaps > 60 s between files are treated like recording gaps in cohort A; every annotated seizure is taken as confirmed and spontaneous (no provocation information exists); E5 keeps the v1 definition and E4 is applied by time stamp; calibration follows the frozen rule. Scripts: src/epifound/v5p/chbmit.py and scripts/40_v5p_frozen/chbmit_{inventory,extract_features,build_cohort}.py; the adapter files are part of the code manifest.

**Cohort A (batch 2).** Recordings were processed by the same scripts as the development cohort (inventory and annotation parsing, feature caches, the frozen feature and quality-control stage, structuring of clinician annotations into seizure events, provocation periods and recording gaps), with only the data paths changed. The evaluability table uses the frozen scheme's own calibration rule (v5p meta) for E3, as the original method is not run on validation data; E1, E2 and E5 are computed exactly as for the development cohort. Cohort A as delivered on 17 October 2026: 21 recordings, 653 EDF files, 1290.7 h at 500 Hz with all 19 standard electrodes; 82.8% of 464 656 windows passed the frozen quality control; 20 of 21 recordings completed calibration (D20 had 1104 clean windows in the calibration period, below the 1440 required); 149 annotated events, 141 confirmed, none provoked; 51 seizures in 19 children met the evaluability criteria; 1056.6 h of monitoring after calibration. After unblinding, D20 was found to be the same recording as the uncalibrated child of the development cohort (identical file names, sizes and start times); it contributed no monitored time and no evaluable seizure to any development or validation figure and is excluded from the cohort description in the main text (20 children, 1266.7 h, 83.2% valid windows, 126 confirmed seizures). Folder names supplied with the recordings were replaced by study codes before processing.

### Supplementary Note S1. Chance tests and power

**Analytic.** A Poisson predictor with rate λ (the observed false-alarm rate per hour of alarm-eligible time) places at least one alarm in a window of length W with probability $1 - \exp(-\lambda W)$; W = 29.5 min for the 0.5–30 min window, 19.5 min for the 20-min window and 89.5 min for the original method. The number of detected seizures is compared with Binomial(n, $1 - \exp(-\lambda W)$) by a one-sided test. **Permutation.** Each patient's alarms are re-placed uniformly over that patient's alarm-eligible windows, keeping their number and minimum spacing; 1000 permutations (2000 for the original method). **Local jitter.** Each alarm is moved to a uniformly random time within ±J of its original time (J = 1, 3 or 6 h), truncated to the monitoring period; 2000 permutations. **Clock-matched AUC.** Pre-ictal segments are scored only against sliding (10-min step) interictal blocks of the same patient, more than 8 h from any seizure, whose centre clock time is within 2 h of the centre of the pre-ictal segment (at least five such blocks). **Segment AUC.** Pre-ictal segment = one warning window ending 0.5 min before onset; interictal segments = non-overlapping blocks of the same length more than 4 h (main analysis) or 8 h (horizon analyses) from any confirmed seizure, with at least 50% valid windows; segment score = mean z; AUC per patient; one-sided Wilcoxon signed-rank test of the per-patient AUCs against 0.5.

**Power** (one-sided α = 0.05, analytic test with chance 20.8%): with the development effect (33.3%) and with half of it (27.1%). Pre-specification v2 was drafted when about 30 children were expected in batch 2; batch 2 delivered 21 recordings (20 children after the duplicate of a development recording was excluded) with 51 evaluable seizures in 19 children.

| Evaluable seizures in validation | Detections needed for $p < 0.05$ | Power at 33.3% | Power at 27.1% (half effect) |
|---|---|---|---|
| 45 (batch 2 alone, 21 recordings as announced on 14 October 2026) | ≥ 15 | 0.56 | 0.21 |
| 50 (CHB-MIT, counted from annotations and frozen quality control on 14 October 2026) | ≥ 16 | 0.63 | 0.26 |
| 51 (batch 2, counted from annotations and frozen quality control on 17 October 2026; 19 children) | ≥ 17 | 0.55 | 0.20 |
| 65 | ≥ 20 | 0.71 | 0.29 |
| 86 | ≥ 25 | 0.83 | 0.38 |
| 95 (batch 2 expected 45 + CHB-MIT 50) | ≥ 27 | 0.87 | 0.42 |
| 101 (counted: batch 2 51 + CHB-MIT 50; the pooled primary analysis) | ≥ 29 | 0.86 | 0.39 |
| 130 | ≥ 36 | 0.93 | 0.47 |
| 160 | ≥ 43 | 0.97 | 0.55 |

Permutation p-values are reported as the proportion of permutations reaching the observed sensitivity; with the $(b + 1)/(m + 1)$ convention [Phipson & Smyth 2010] the primary 2000-permutation value 0.002 becomes 0.0025.

The pooled analysis of both validation cohorts is the pre-specified primary validation analysis; cohorts are also reported separately and no cohort may be dropped after results are seen.

### Supplementary Table S5. Feature-set screening on the development cohort (segment-level AUC, independent of the CUSUM) and the ECG-only control

Screening was done before the alarm layer was fixed; weights are learned on the other patients. The ECG-only row uses three heart-rate features from the ECG channel with the same baseline and state conditioning; it is a negative control for EEG specificity, not a candidate scheme.

| Feature set | 30-min window: mean AUC / pooled AUC / p | 20-min window: mean AUC / pooled AUC / p | Sensitivity at 20% / 30% time in warning (30 min) |
|---|---|---|---|
| All 19 global features, learned weights | 0.684 / 0.648 / 0.011 | 0.653 / 0.649 / 0.039 | 40% / 47% |
| 16 EEG features, learned | 0.644 / 0.618 / 0.047 | 0.624 / 0.620 / 0.077 | 33% / 46% |
| **Core 7 features, learned (frozen)** | **0.672 / 0.641 / 0.024** | **0.673 / 0.646 / 0.017** | 37% / 46% |
| Core 3, equal weights | 0.661 / 0.613 / 0.016 | 0.682 / 0.619 / 0.012 | 32% / 40% |
| First eigenvalue fraction only | 0.691 / 0.649 / 0.002 | 0.705 / 0.659 / 0.001 | 29% / 34% |
| Spectral entropy only | 0.606 / 0.560 / 0.11 | 0.623 / 0.568 / 0.042 | 24% / 45% |
| Hjorth complexity only | 0.626 / 0.586 / 0.055 | 0.643 / 0.582 / 0.025 | 33% / 47% |
| δ log power only | 0.492 / 0.476 / 0.55 | 0.499 / 0.465 / 0.52 | 27% / 36% |
| Original two features, learned | 0.573 / 0.530 / 0.15 | 0.598 / 0.535 / 0.10 | 16% / 38% |
| ECG only, three features, learned (control) | 0.644 / 0.635 / 0.029 | 0.611 / 0.631 / 0.059 | 27% / 31% |

In the same screening, heart rate from the ECG channel was elevated throughout the 4 h before seizures (about +0.35 to +0.7 SD relative to the state-matched baseline; project records of 29 September 2026), i.e. a slow drift rather than a short pre-ictal change, compatible with an autonomic or medication-related process.

Source: project screening records of 29 September 2026. The single-feature eigenvalue-fraction scheme scored slightly higher than the frozen seven-feature statistic in screening but lower in the full nested evaluation with the alarm layer (11/45 versus 15/45 at the 30-min window; the screening table of the project records).

### Supplementary Note S2. Vigilance-state tabulation of the original method

The internal report of 29 September 2026 tabulated the original method's normalised statistic by clinician-marked state as sleep −0.10 (19 777 windows) and wake +0.62 (13 366 windows) in 15 children, a difference of 0.72 SD. The script that produced that table was not retained. A read-only recomputation from the stored per-window outputs with the stated rule (windows within 5 min after a mark, more than 2 h from any seizure) gives, for the 16 children with both kinds of mark, sleep −0.06 (21 434 windows) and wake +0.56 (15 455 windows), a difference of 0.62 SD; every variant of the rule that we tried gives 0.62–0.64 SD. The reproducible values are reported in the main text and Fig. 2A. The conclusion (the state difference is about three times the pre-ictal effect of ≈0.22 SD) is unchanged.

### Supplementary Table S6. Code manifest

SHA-256 digests (file bytes) of the scripts and configuration files of the frozen scheme as deposited on 2026-10-17 16:19 UTC, immediately before the locked runs. The covered set is the frozen runner's actual import closure plus the feature-extraction, freeze, adapter and configuration files, with the Python/NumPy/SciPy/scikit-learn versions. The manifest was first generated on 13 October 2026, when v2 took effect and the repository was restructured into a package layout, and regenerated as the clock-matched test, the CHB-MIT adapter and the confound-only comparator were added (Appendix B §12 records each version and its digest); this deposited version supersedes them. The digest of `config/confound_frozen.json` is that of the original file; the public copy of the deposit differs in its redacted list of training recordings (Note S3).

| File | SHA-256 (first 16 hex) |
|---|---|
| config/confound_frozen.json | 6e57a638f7a97124… |
| config/v5p_frozen.json | f2ea8d17d497eae6… |
| config/v5p_frozen_H20.json | d826b4351dd2942f… |
| pyproject.toml | 6d36d8693e40b328… |
| scripts/10_cohort/cohort_characterize.py | b557ff549221691d… |
| scripts/10_cohort/cohort_inventory.py | aafc2854295d7baa… |
| scripts/10_cohort/run_batch2_preprocessing.sh | 80f18a15d9e2a390… |
| scripts/20_original_method/cohort_extract_features.py | 9fe7fb7afb53f623… |
| scripts/30_v5_development/cohort_extract_features_x.py | 9fe025365dd06d29… |
| scripts/40_v5p_frozen/batch2_build_cohort.py | 5b08f2b7dfd32d18… |
| scripts/40_v5p_frozen/chbmit_build_cohort.py | 6e24b489a9ba4365… |
| scripts/40_v5p_frozen/chbmit_extract_features.py | f36a48feb6048bae… |
| scripts/40_v5p_frozen/chbmit_inventory.py | ecf7ba9d665e2b02… |
| scripts/40_v5p_frozen/cohort_extract_features_p.py | c96b314daf540c6e… |
| scripts/40_v5p_frozen/confound_freeze.py | 4256ab7447fbc416… |
| scripts/40_v5p_frozen/make_code_manifest.py | 82b44803900ab89f… |
| scripts/40_v5p_frozen/make_validation_root.py | 7fad087651497f59… |
| scripts/40_v5p_frozen/run_locked_validation.sh | d61b634702c0bf02… |
| scripts/40_v5p_frozen/v5p_freeze.py | 250bd7b403c32114… |
| scripts/40_v5p_frozen/v5p_meta.py | 74240cabc7423dca… |
| scripts/40_v5p_frozen/v5p_run.py | 096caaeddd01a97b… |
| scripts/60_paper/verify_chbmit.py | 3941f88903b9cde4… |
| src/epifound/**init**.py | 8118ece8a093d454… |
| src/epifound/common.py | d95ff201a47c3ea6… |
| src/epifound/paths.py | cf434a3fd2be53a4… |
| src/epifound/segments.py | de595584ed391b29… |
| src/epifound/spc2/**init**.py | c46edcb604794d35… |
| src/epifound/spc2/baseline.py | 3b05205a756ad094… |
| src/epifound/spc2/calibrate.py | 08d2127eeb21d5ad… |
| src/epifound/spc2/config.py | 5d026e547b753721… |
| src/epifound/spc2/cusum.py | aed7beb9d7ed74ea… |
| src/epifound/spc2/evaluate.py | 757813f92467abc3… |
| src/epifound/spc2/features.py | c4aa3d7a33bbd261… |
| src/epifound/spc2/quality.py | 32b6c1f1e3514fbc… |
| src/epifound/spc2/retro.py | 49abee329a5d3003… |
| src/epifound/v5p/**init**.py | 5c6e6c2883987dc2… |
| src/epifound/v5p/auc.py | 8a9333d5ba20e872… |
| src/epifound/v5p/causal.py | 84a555b83b6ea1ff… |
| src/epifound/v5p/chbmit.py | e4ba5695fabdfa52… |
| src/epifound/v5p/clockmatch.py | 02901951b4928e77… |
| src/epifound/v5p/confound.py | 7d22da07c4cfd99c… |
| src/epifound/v5p/core.py | 8ca79c4270d6531e… |
| src/epifound/v5p/dev.py | 02888eaae3f9f122… |
| src/epifound/v5p/devload.py | c127ee1fc789f713… |
| src/epifound/v5p/jitter.py | 9eaf1b6ca181e6e5… |
| src/epifound/v5p/meta.py | 0d62bfd37efa200c… |
| src/epifound/v5p/screen.py | 0907dfda696f38cd… |
| src/epifound/v5p/state_model.py | c3bc8af321c80b90… |

### Supplementary Note S4. Comparators under their own protocols

**Multivariate SPC ($T^2$ and Q charts) [Rukhsar et al. 2019].** Under the published protocol (40 min of interictal modelling per patient, a per-patient choice of the cumulative contribution ratio of the principal components, a 60-min pre-ictal period and no chance comparison) our re-implementation detected 1 of 50 seizures in the 14 children with suitable data at 0.01 false alarms per hour (9 alarms in 977 h); with the contribution ratio fixed at 98% it detected none and was in the 'pre-ictal' state for essentially all of the time (median 100%). The published result (88.9% sensitivity at 0.39/h) was obtained on CHB-MIT (10 patients, 90 seizures) with patient-specific, segment-wise evaluation. Under our rules (calibration-period modelling, leave-one-patient-out choice of the contribution ratio, control-limit multiplier and firing-power threshold, continuous simulation) it detected 4 of 45 evaluable seizures (8.9%) at 0.579/h (387 false alarms in 668 h; analytic chance 24.8%, p = 0.998; permutation 18.8%, p = 0.998; ±3 h jitter 22.8%, p = 1.00). Source: comparator report CMP_mspc_H30/report.txt in the analysis outputs.

**Per-channel pre-ictal-pattern criterion [Segal et al. 2025].** The criterion declares a pre-ictal interval on a channel when the median Mahalanobis distance of the channel's 10-dimensional feature vector (spectral entropy, Hjorth mobility and complexity, five relative band powers, line length and crest factor; the published Higuchi fractal dimension and detrended fluctuation analysis were replaced by line length and crest factor) from a 2-h baseline taken 8–6 h before the event exceeds the 99th percentile of the baseline distances for at least 15 min within the 6 h before the event. Real events were the 27 confirmed seizures of 17 children with 8 h of recording and no other confirmed seizure in the preceding 8 h; control events were 150 random times in 15 children with no seizure of any kind from 8 h before to 2 h after.

| Threshold | Real seizures | Seizure-free control epochs | Fisher exact p |
|---|---|---|---|
| ≥ 1 channel | 20/27 = 74.1% | 104/150 = 69.3% | 0.40 |
| ≥ 3 channels | 17/27 = 63.0% | 85/150 = 56.7% | 0.35 |
| ≥ 5 channels | 16/27 = 59.3% | 72/150 = 48.0% | 0.19 |
| ≥ 10 channels | 13/27 = 48.1% | 42/150 = 28.0% | 0.034 |

Channels meeting the criterion: median 8 (IQR 0–15) before seizures versus 4 (0–10) before control epochs (Mann–Whitney p = 0.057). Start of the 'pre-ictal interval': 204 ± 96 min (median 201) before seizures and 233 ± 103 min (median 258) before control epochs; published 83 ± 60 min. At least one detection in 14 of 17 children with seizures (published 17 of 19) and in all 15 children with control epochs. Source: analysis log segal_control_p.log.

### Supplementary Note S5. Simulation calibration of the chance tests (before unblinding)

Each test was applied to effect-free series (300 replicates per null model; permutation 200 and jitter 300 resamples per replicate) generated from the development cohort's out-of-sample z with the alarm parameters of the nested leave-one-patient-out evaluation. Null A: for each child, z = constant + two circadian harmonics + admission-day trend + state (+ state × first harmonic) + AR(1) residuals with bootstrapped innovations, fitted outside ±30/60 min of seizures; missing and unmonitored windows and seizure times as in the real data. Null C: the same structured mean with moving-block bootstrap of the residuals (blocks of 1 h), which keeps longer memory and the tails. Null B: the real z circularly shifted over its valid windows, which keeps every serial property and only breaks the alignment with seizure times and clock. Entries are the proportion of replicates with p < 0.05 (the false-positive rate at α = 0.05); the real-data p-values are the calibration script's own recomputation with 200 permutations and 300 jitter resamples per test and therefore differ slightly from the pre-specified runs reported in Table 2 (1000 permutations, 2000 jitter resamples). Script: scripts/30_v5_development/a1_null_calibration.py.

| Test | Real data p | Null A | Null C | Null B |
|---|---|---|---|---|
| Analytic (Poisson random predictor) | 0.034 | 3.3% | 3.0% | 0.3% |
| Uniform permutation (alarm shuffling) | 0.010 | 15.3% | 16.7% | 3.7% |
| Local jitter ±1 h | 0.030 | 5.0% | 8.3% | 4.7% |
| Local jitter ±3 h | 0.020 | 1.7% | 2.3% | 1.3% |
| Local jitter ±6 h | 0.020 | 1.7% | 2.3% | 2.3% |
| Per-patient segment AUC (Wilcoxon > 0.5) | 0.024 | 31.7% | 35.0% | 3.3% |
| Clock-matched segment AUC (±2 h) | 0.082 | 16.7% | 19.7% | 6.7% |
| **Primary rule: analytic ∧ ±1 h jitter** | passed | 2.0% | 3.0% | 0.3% |

Mean sensitivity under the nulls: A 0.158 at 0.297/h; C 0.238 at 0.545/h; B 0.159 at 0.477/h (real data 15/45 at 0.473/h). Reading: the alarm-level primary rule and the analytic test hold their level under all three nulls; the uniform permutation and, above all, the segment-level AUC tests reject effect-free series far too often when the cohort's clock, state and admission-day structure is present, because pre-ictal segments sample that structure at seizure-typical times. Segment AUC p-values in the main text are therefore nominal and descriptive; the validation decision uses the primary rule only, as pre-specified.

### Supplementary Note S6. Confound-only predictor (clock time, vigilance state, days since admission)

A logistic predictor was trained leave-one-patient-out on the same labels as the frozen weights (windows 15–0.5 min before lead seizures versus windows more than 4 h from any seizure, children weighted equally within class; L2, C = 1.0) from eight non-EEG inputs: two circadian harmonics of clock time, days since the start of recording, the scheme's own EEG-estimated sleep–wake state, and state × first harmonic. Its score was rank-transformed per child (Blom, over all monitored windows: non-causal, favouring the predictor) and scored exactly like the frozen statistic. Script: scripts/30_v5_development/a2_confound_predictor.py.

| Scheme and parameters | Sensitivity (95% CI) | False alarms/h | Analytic chance / p | Permutation chance / p | ±1 h jitter chance / p |
|---|---|---|---|---|---|
| Frozen EEG scheme, fold parameters (main result) | 15/45 = 33.3% (21.4–47.9%) | 0.473 | 20.8% / 0.034 | 16.3% / 0.000 | 23.7% / 0.033 |
| Confound-only, nested leave-one-patient-out parameters (same protocol) | 13/45 = 28.9% (17.7–43.4%) | 0.410 | 18.2% / 0.055 | 17.0% / 0.024 | 23.3% / 0.130 |
| Confound-only, frozen EEG parameters (above the cap) | 19/45 = 42.2% (29.0–56.7%) | 0.844 | 34.0% / 0.155 | 30.8% / 0.066 | 40.8% / 0.474 |
| Confound-only, in-sample best parameters k = 0.75, h = 25.229, N = 3 (favouring the predictor) | 16/45 = 35.6% (23.2–50.2%) | 0.410 | 18.3% / 0.004 | 16.9% / 0.000 | 25.2% / 0.006 |

| Segment-level metric (30 min) | Frozen EEG scheme | Confound-only |
|---|---|---|
| Per-patient segment AUC, mean (children > 0.5 / with data) | 0.672 (10/13) | 0.594 (10/13) |
| Wilcoxon p (AUC > 0.5; nominal, see Note S5) | 0.0239 | 0.0732 |
| Pooled AUC | 0.641 | 0.591 |
| Area under the sensitivity–time-in-warning curve | 0.588 | 0.627 |
| Sensitivity at 10% / 20% / 30% time in warning | 18% / 37% / 46% | 20% / 36% / 52% |

Chance levels and p-values in this note are recomputed by the comparator script with its own random seeds and resample counts and therefore differ slightly from the pre-specified values in Table 2.

Paired per-child difference in segment AUC (EEG − confound-only): mean +0.078, EEG higher in 7 of 13 children, one-sided Wilcoxon p = 0.190. Reading: most of the frozen scheme's development-cohort performance is reproduced by a predictor that knows only the time of day, the estimated vigilance state and the day of admission; the EEG-specific increment is small and not statistically established.

### Supplementary Note S7. Horizon analyses: full results

The per-patient AUC of the full pre-ictal segment fell with the horizon: 0.710 at 10 min (13 children; one-sided Wilcoxon p = 0.013), 0.677 at 20 min (p = 0.029), 0.667 at 30 min (p = 0.048), 0.597 at 60 min (12 children; p = 0.14), 0.621 at 120 min (11; p = 0.13), 0.474 at 240 min and 0.472 at 360 min (9 children each; Fig. 4A); in the 9 children present at every horizon the 240-min AUC was 0.16 lower than the 30-min AUC (bootstrap 95% CI −0.32 to −0.03; two-sided Wilcoxon p = 0.074). Disjoint rings located the signal in the last 30 min: AUC 0.698 at 0.5–5 min (9 of 13 above 0.5; p = 0.021), 0.735 at 5–10 min (11 of 13; p = 0.003), 0.654 at 10–20 min (9 of 13; p = 0.040) and 0.597 at 20–30 min (8 of 13; p = 0.12), but 0.50–0.55 in every ring from 30 min to 8 h before onset (Fig. 4B). The percentile of 30-min block means of z within each child's interictal blocks was at chance from 8 h to 30 min before onset (−8 to −2 h: 0.498; slope −0.001 per hour, p = 0.97), rose to 0.667 in the last 30 min (against 0.533 over −8 to −0.5 h, p = 0.13; against the preceding block, 0.512, p = 0.019), and the mean over the 8 h before onset (0.544) was no higher than over 1 –6 h after onset (0.604; p = 0.27; Fig. 4C). A surrogate circularly shifting each child's seizure times (2000 shifts, preserving autocorrelation) confirmed the last-30-min excess (p = 0.002; positive in 13 of 16 children) and the absence of a far pre-ictal excess (p = 0.82), but with shifts restricted to multiples of 24 h, preserving clock time, the last-30-min excess was no longer significant (p = 0.25; 9 children). There is thus no pre-ictal state lasting hours: only the last 10–30 min stand out, and the mean post-ictal elevation is at least as large as the pre-ictal one.

Scored against clock-matched interictal blocks only (10 children with at least five matched blocks), the 30-min AUC of the same seizures fell from 0.664 unmatched to 0.641 matched (above 0.5 in 5 of 10; p = 0.082) and the 10-min AUC from 0.648 to 0.596 (p = 0.22), whereas subtracting the leave-one-patient-out cohort clock profile before scoring left the 30-min AUC at 0.679 (9 of 13; p = 0.013). Part of the apparent signal is therefore clock-time structure; the clock-matched AUC, the most conservative test, is a key secondary validation endpoint.

**Locked validation at the 20-min window.** The run with the 20-min configuration (secondary operating point 2) also yields the other endpoints at that window. Only its sensitivity at the primary cap was pre-specified; the rest are listed for completeness and are nominal. Per-patient segment AUC (20-min pre-ictal segments): pooled 0.599 (p = 0.021); cohort A 0.577 (p = 0.094); cohort B 0.629 (p = 0.051). Clock-matched segment AUC (20 min, ±2 h): pooled 0.563 (11 of 18 above 0.5; p = 0.16); cohort A 0.522 (6 of 10 above 0.5; p = 0.31); cohort B 0.614 (5 of 8 above 0.5; p = 0.16). Sensitivity at the 0.25/h cap with the 20-min window: pooled 10 of 101 at 0.277/h (analytic p = 0.37; permutation p = 0.083); cohort A 2 of 51 at 0.272/h (analytic p = 0.94; permutation p = 0.74); cohort B 8 of 50 at 0.288/h (analytic p = 0.075; permutation p = 0.016). Under effect-free series with the cohort's time structure, the uniform permutation test rejects in up to 16.7% and the segment-AUC test in up to 35.0% of replicates (Note S5); none of these values entered the decision rule.

### Supplementary Table S7. Per-patient results of the locked validation (primary operating point)

Frozen parameters applied once to each validation cohort; false-alarm rate per hour of alarm-eligible interictal time; segment AUC recomputed from the stored out-of-sample z with the development rule (at least one pre-ictal and five interictal segments). Nominal statistics only (Note S5).

| Cohort | Case | Evaluable seizures | Detected | False alarms | Alarm-eligible interictal time (h) | False alarms per hour | Segment AUC (pre-ictal / interictal segments) |
|---|---|---|---|---|---|---|---|
| cohort A (batch 2) | A01 | 4 | 0 | 21 | 28.7 | 0.731 | 0.630 (3 / 54) |
| cohort A (batch 2) | A02 | 1 | 0 | 1 | 5.4 | 0.186 | 0.600 (1 / 5) |
| cohort A (batch 2) | A03 | 3 | 1 | 41 | 53.9 | 0.760 | 0.754 (3 / 126) |
| cohort A (batch 2) | A04 | 0 | 0 | 1 | 0.1 | 6.667 | — |
| cohort A (batch 2) | A05 | 4 | 1 | 17 | 37.5 | 0.454 | 0.646 (4 / 70) |
| cohort A (batch 2) | A06 | 2 | 0 | 0 | 6.9 | 0.000 | — |
| cohort A (batch 2) | A07 | 2 | 0 | 0 | 9.5 | 0.000 | — |
| cohort A (batch 2) | A08 | 5 | 0 | 6 | 24.5 | 0.245 | 0.091 (5 / 22) |
| cohort A (batch 2) | A09 | 7 | 2 | 79 | 105.7 | 0.747 | 0.491 (7 / 255) |
| cohort A (batch 2) | A10 | 1 | 0 | 57 | 97.2 | 0.586 | 0.043 (1 / 257) |
| cohort A (batch 2) | A11 | 2 | 0 | 1 | 8.3 | 0.120 | — |
| cohort A (batch 2) | A12 | 2 | 0 | 4 | 1.6 | 2.581 | — |
| cohort A (batch 2) | A13 | 1 | 0 | 0 | 3.2 | 0.000 | — |
| cohort A (batch 2) | A14 | 4 | 0 | 1 | 9.1 | 0.110 | 0.292 (4 / 6) |
| cohort A (batch 2) | A15 | 3 | 1 | 43 | 74.8 | 0.575 | 0.597 (3 / 177) |
| cohort A (batch 2) | A16 | 3 | 0 | 13 | 45.0 | 0.289 | 0.810 (3 / 91) |
| cohort A (batch 2) | A17 | 2 | 0 | 26 | 41.5 | 0.626 | 0.569 (2 / 94) |
| cohort A (batch 2) | A18 | 1 | 0 | 5 | 10.0 | 0.501 | 0.688 (1 / 16) |
| cohort A (batch 2) | A19 | 2 | 0 | 0 | 3.2 | 0.000 | — |
| cohort A (batch 2) | A20 | 2 | 1 | 70 | 79.1 | 0.885 | 0.681 (2 / 221) |

| Cohort | Case | Evaluable seizures | Detected | False alarms | Alarm-eligible interictal time (h) | False alarms per hour | Segment AUC (pre-ictal / interictal segments) |
|---|---|---|---|---|---|---|---|
| cohort B (CHB-MIT) | chb01 | 4 | 2 | 8 | 11.3 | 0.705 | 0.500 (4 / 13) |
| cohort B (CHB-MIT) | chb02 | 3 | 0 | 6 | 15.3 | 0.392 | 0.633 (1 / 30) |
| cohort B (CHB-MIT) | chb03 | 0 | 0 | 20 | 6.9 | 2.894 | — |
| cohort B (CHB-MIT) | chb04 | 3 | 0 | 38 | 66.3 | 0.573 | 0.667 (1 / 135) |
| cohort B (CHB-MIT) | chb05 | 3 | 1 | 13 | 15.8 | 0.823 | 0.667 (3 / 27) |
| cohort B (CHB-MIT) | chb06 | 5 | 0 | 14 | 25.3 | 0.553 | 0.348 (5 / 46) |
| cohort B (CHB-MIT) | chb07 | 3 | 1 | 15 | 29.6 | 0.507 | 0.871 (3 / 62) |
| cohort B (CHB-MIT) | chb08 | 0 | 0 | 1 | 0.8 | 1.316 | — |
| cohort B (CHB-MIT) | chb09 | 4 | 1 | 2 | 37.8 | 0.053 | 0.684 (4 / 57) |
| cohort B (CHB-MIT) | chb10 | 5 | 3 | 12 | 23.0 | 0.522 | 0.976 (4 / 31) |
| cohort B (CHB-MIT) | chb11 | 0 | 0 | 4 | 14.1 | 0.285 | — |
| cohort B (CHB-MIT) | chb13 | 2 | 0 | 0 | 6.0 | 0.000 | 0.750 (2 / 6) |
| cohort B (CHB-MIT) | chb14 | 1 | 0 | 2 | 0.5 | 3.774 | — |
| cohort B (CHB-MIT) | chb15 | 7 | 4 | 8 | 5.2 | 1.538 | — |
| cohort B (CHB-MIT) | chb16 | 3 | 0 | 0 | 2.5 | 0.000 | — |
| cohort B (CHB-MIT) | chb17 | 0 | 0 | 0 | 3.7 | 0.000 | — |
| cohort B (CHB-MIT) | chb18 | 1 | 0 | 12 | 9.9 | 1.211 | 0.148 (1 / 27) |
| cohort B (CHB-MIT) | chb19 | 2 | 0 | 12 | 8.1 | 1.476 | — |
| cohort B (CHB-MIT) | chb20 | 1 | 0 | 3 | 5.1 | 0.591 | — |
| cohort B (CHB-MIT) | chb21 | 1 | 1 | 8 | 9.8 | 0.821 | 0.826 (1 / 23) |
| cohort B (CHB-MIT) | chb22 | 2 | 0 | 11 | 6.4 | 1.719 | — |
| cohort B (CHB-MIT) | chb23 | 0 | 0 | 0 | 6.6 | 0.000 | — |

**Supplementary Table S8. Clinical characteristics transcribed from the unit's video-EEG monitoring reports.** One report per recording; sex, age, clinical diagnosis at admission and medication are taken from the report header, seizure types from its EEG-diagnosis section (onset side as stated by the reporting physician), and the length of the monitoring period from the report's start and end times. To protect the children's privacy, age at recording is given in five-year bands of completed years, intervals since earlier procedures in whole years, and the monitoring period as a length in hours without calendar dates; where the delivered EDF files span more than 12 h less than the reported period, their span is given in brackets. The EEG diagnosis was 'abnormal EEG' in every report. 'Yes' under dose reduction means that the report documents a reduction or withdrawal of antiseizure medication during monitoring, naming the drug where the report does; 'not stated' means the report is silent, not that no change was made. Aetiology and imaging are not part of these reports. D20 is listed once (development cohort); a report was delivered for every child. The report of the pilot child (IY7776) was received as a photograph of the printed report and transcribed. Abbreviations: ASM, antiseizure medication; BRV, brivaracetam; CBZ, carbamazepine; CLB, clobazam; LCM, lacosamide; LEV, levetiracetam; LTG, lamotrigine; OXC, oxcarbazepine; PER, perampanel; TPM, topiramate; VNS, vagus nerve stimulation; VPA, valproate; ZNS, zonisamide.

| Cohort | Study code | Sex | Age band (y) | Clinical diagnosis at admission | Antiseizure medication at admission | Dose reduction or withdrawal during monitoring, as documented | Seizure types recorded (onset side) | Non-epileptic events recorded | Monitoring period in the report (h) |
|---|---|---|---|---|---|---|---|---|---|
| development | D01 | M | 15–19 | paroxysmal symptoms under evaluation | none | no medication | focal (right onset) | yes | 24 |
| development | D02 | M | 5–9 | epilepsy | LCM | not stated | focal (left onset) | no | 23 |
| development | D03 | M | 0–4 | epilepsy | OXC, VPA, CLB, PER | not stated | focal (left onset) | yes | 24 |
| development | D04 | M | 10–14 | epilepsy | VPA, PER | yes: PER and VPA tapered and stopped | focal (left onset) | no | 152 |
| development | D05 | F | 10–14 | epilepsy | OXC, LEV | not stated | focal (right onset) | yes | 54 |
| development | D06 | M | 10–14 | epilepsy | LEV, LTG, LCM | yes: LCM tapered and stopped | focal (right onset) | yes | 115 |
| development | D07 | M | 10–14 | epilepsy; corpus callosotomy 5 years and VNS implantation 8 years earlier | CLB, OXC, LCM | not stated | focal (left onset) | yes | 24 |
| development | D08 | M | 5–9 | epilepsy | OXC | yes: reduction mentioned in the report text (drug not named) | focal (left onset) | no | 142 (EDF files delivered span 126 h) |
| development | D09 | M | 15–19 | epilepsy | none | no medication | focal (left onset) | no | 24 |
| development | D10 | M | 15–19 | epilepsy | OXC, PER | yes: reduction mentioned in the report text (drug not named) | focal (left onset); focal to bilateral tonic-clonic (ictal EEG description) | no | 166 |
| development | D11 | M | 0–4 | epilepsy | LEV, OXC, CLB, TPM | not stated | focal (left onset) | yes | 25 |
| development | D12 | F | 10–14 | epilepsy | LEV, LCM | not stated | focal (left onset) | no | 46 |
| development | D13 | M | 5–9 | epilepsy | VPA, PER | not stated | focal (right onset) | no | 45 |
| development | D14 | M | 0–4 | epilepsy | VPA, ZNS, LCM, BRV | not stated | focal (left onset) | no | 73 |
| development | D15 | M | 5–9 | epilepsy | LEV, OXC | not stated | focal (right onset) | yes | 97 |

| Cohort | Study code | Sex | Age band (y) | Clinical diagnosis at admission | Antiseizure medication at admission | Dose reduction or withdrawal during monitoring, as documented | Seizure types recorded (onset side) | Non-epileptic events recorded | Monitoring period in the report (h) |
|---|---|---|---|---|---|---|---|---|---|
| development | D16 | F | 0–4 | epilepsy | VPA | yes: VPA dose reduced | focal (right onset) | no | 66 |
| development | D17 | M | 10–14 | epilepsy | OXC, PER | not stated | focal (right onset) | yes | 24 |
| development | D18 | M | 0–4 | epilepsy | OXC, LEV | yes: LEV dose reduced | focal (right onset) | no | 74 |
| development | D19 | M | 5–9 | epilepsy | LEV | not stated | focal (left onset) | no | 24 |
| development | D20 | F | 5–9 | epilepsy | none | no medication | focal (left onset) | yes | 25 |
| development | IY7776 | F | 5–9 | epilepsy; right hemispherotomy about 4 years earlier | LCM, PER | not stated | focal (left onset) | yes | 168 (EDF files delivered span 98 h) |
| cohort A (batch 2) | A01 | F | 10–14 | epilepsy | OXC, BRV | yes: BRV dose reduced | focal (right onset) | yes | 66 |
| cohort A (batch 2) | A02 | F | 0–4 | epilepsy | CLB, LEV, VPA, LCM | not stated | focal (right onset) | no | 23 |
| cohort A (batch 2) | A03 | M | 10–14 | epilepsy | VPA, LCM, CLB | yes: CLB dose reduced | focal (left onset) | yes | 96 |
| cohort A (batch 2) | A04 | M | 5–9 | epilepsy | LEV, OXC | not stated | focal (left onset) | yes | 19 |
| cohort A (batch 2) | A05 | M | 5–9 | epilepsy | LCM, PER | yes: reduction mentioned in the report text (drug not named) | focal (right onset) | yes | 69 |
| cohort A (batch 2) | A06 | M | 5–9 | epilepsy | LCM, LEV | not stated | focal (left onset) | yes | 24 |
| cohort A (batch 2) | A07 | M | 10–14 | epilepsy | OXC, LCM | not stated | focal (left onset) | no | 24 |
| cohort A (batch 2) | A08 | F | 5–9 | epilepsy | LCM | not stated | focal (left onset) | yes | 47 |
| cohort A (batch 2) | A09 | M | 0–4 | epilepsy | LCM, TPM, VPA | yes: VPA and LCM tapered and stopped | electrographic seizure (no clinical sign); focal (left onset) | yes | 174 |
| cohort A (batch 2) | A10 | M | 10–14 | epilepsy | VPA, PER | not stated | focal (unspecified onset) | no | 163 |
| cohort A (batch 2) | A11 | F | 10–14 | epilepsy; bilateral SEEG with radiofrequency thermocoagulation about 3 years earlier | OXC, LEV, PER | not stated | focal (left onset) | no | 23 |
| cohort A (batch 2) | A12 | F | 5–9 | epilepsy | OXC, PER | not stated | focal (left onset) | no | 24 |
| cohort A (batch 2) | A13 | F | 10–14 | epilepsy | PER, LCM, CLB | not stated | electrographic seizure (no clinical sign); focal (right onset) | no | 24 |
| cohort A (batch 2) | A14 | M | 5–9 | epilepsy | LCM, CBZ | not stated | focal (left onset) | no | 30 |
| cohort A (batch 2) | A15 | M | 10–14 | epilepsy | LTG, OXC, VPA | yes: reduction mentioned in the report text (drug not named) | focal (right onset) | no | 121 |
| cohort A (batch 2) | A16 | F | 10–14 | epilepsy | OXC, TPM, CLB | yes: CLB dose reduced | focal (right onset) | no | 70 |

| Cohort | Study code | Sex | Age band (y) | Clinical diagnosis at admission | Antiseizure medication at admission | Dose reduction or withdrawal during monitoring, as documented | Seizure types recorded (onset side) | Non-epileptic events recorded | Monitoring period in the report (h) |
|---|---|---|---|---|---|---|---|---|---|
| cohort A (batch 2) | A17 | M | 15–19 | epilepsy | LEV | yes: LEV dose reduced | focal (left onset); focal to bilateral tonic-clonic (ictal EEG description) | no | 90 |
| cohort A (batch 2) | A18 | F | 5–9 | epilepsy | VPA, LTG, LEV, CLB | not stated | focal (left onset); status epilepticus | no | 24 |
| cohort A (batch 2) | A19 | M | 5–9 | epilepsy | PER | not stated | focal (right onset) | no | 24 |
| cohort A (batch 2) | A20 | M | 5–9 | epilepsy | LEV, LTG, LCM, VPA | yes: reduction mentioned in the report text (drug not named) | focal (left onset); focal (right onset) | yes | 144 |

**Supplementary Table S9. Post hoc analyses added at revision (2026-11-05): the child as the unit of analysis, and reported seizure-onset side.** Primary operating point (0.5–30 min window; development cohort nested leave-one-patient-out; validation cohorts, frozen parameters). For each child with at least one evaluable seizure, the chance probability is the analytic Poisson level at that child's own false-alarm rate, $p_i = 1 − \exp(−FAR_i × 29.5 \text{ min})$; 'above chance' means observed detections exceed $n_i p_i$. Sign test: exact binomial on the children above versus below chance (one-sided). Wilcoxon: one-sided signed-rank test on per-child sensitivity minus chance. Bootstrap: 20 000 resamples of children, percentile 95% CI and one-sided p (share of resamples with difference ≤ 0). Per-child binomial: one-sided exact test per child, Benjamini–Hochberg q. The first result column resamples children and compares the pooled sensitivity with the pre-specified analytic chance level (pooled false-alarm rate); the second uses each child's own rate. These analyses were not pre-specified.

| Cohort | Children (seizures) | Detected | Pooled sensitivity, % (child-cluster bootstrap 95% CI; p versus the pre-specified analytic chance) | Pooled sensitivity − child-wise chance, % (bootstrap 95% CI; p) | Child-averaged sensitivity − chance, % (95% CI; p) | Children above / below chance | Sign test p | Wilcoxon p | Children with per-child p < 0.05 (q < 0.05) |
|---|---|---|---|---|---|---|---|---|---|
| development cohort (nested LOPO, primary operating point) | 19 (45) | 15 | 33.3 (20.0–45.2; chance 20.8, p = 0.031) | 33.3 − 18.4 = +14.9 (+3.9 to +25.4; p = 0.004) | 28.9 − 17.0 = +11.9 (-0.4 to +24.7; p = 0.029) | 9 / 7 | 0.402 | 0.060 | 0 (0) |
| validation cohort A (batch 2) | 19 (51) | 6 | 11.8 (3.8–19.6; chance 25.5, p = 1.000) | 11.8 − 20.3 = -8.5 (-17.1 to -1.9; p = 0.996) | 9.0 − 19.0 = -10.1 (-19.3 to -2.5; p = 0.998) | 4 / 11 | 0.982 | 0.988 | 0 (0) |
| validation cohort B (CHB-MIT) | 17 (50) | 13 | 26.0 (10.8–39.3; chance 25.9, p = 0.512) | 26.0 − 28.6 = -2.6 (-17.3 to +10.1; p = 0.661) | 21.1 − 30.9 = -9.8 (-27.3 to +7.4; p = 0.864) | 7 / 8 | 0.696 | 0.860 | 0 (0) |
| validation pooled A + B | 36 (101) | 19 | 18.8 (10.2–27.6; chance 25.6, p = 0.933) | 18.8 − 24.4 = -5.6 (-13.6 to +2.0; p = 0.926) | 14.7 − 24.6 = -9.9 (-19.3 to -0.9; p = 0.984) | 11 / 19 | 0.951 | 0.985 | 0 (0) |

| Cohort | Reported onset side | Children (with any detection) | Detected / evaluable | Sensitivity, % (Wilson 95% CI) | Chance, % (child-wise analytic) | p, binomial | Left vs right, Fisher p |
|---|---|---|---|---|---|---|---|
| development cohort | left | 11 (5) | 7 / 26 | 26.9 (13.7–46.1) | 20.1 | 0.258 | 0.35 |
| development cohort | right | 8 (5) | 8 / 19 | 42.1 (23.1–63.7) | 16.1 | 0.006 | |
| validation cohort A | left | 10 (2) | 3 / 30 | 10.0 (3.5–25.6) | 20.6 | 0.962 | 1.00 |
| validation cohort A | right | 7 (2) | 2 / 18 | 11.1 (3.1–32.8) | 17.9 | 0.860 | |
| validation cohort A | both | 1 (1) | 1 / 2 | 50.0 (9.5–90.5) | 35.3 | 0.581 | |
| validation cohort A | unspecified | 1 (0) | 0 / 1 | 0.0 (0.0–79.3) | 25.0 | 1.000 | |
| development + validation A | left | 21 (7) | 10 / 56 | 17.9 (10.0–29.8) | 20.4 | 0.729 | 0.31 |
| development + validation A | right | 15 (7) | 10 / 37 | 27.0 (15.4–43.0) | 17.0 | 0.085 | |
| development + validation A | both | 1 (1) | 1 / 2 | 50.0 (9.5–90.5) | 35.3 | 0.581 | |
| development + validation A | unspecified | 1 (0) | 0 / 1 | 0.0 (0.0–79.3) | 25.0 | 1.000 | |

### Supplementary Note S3. Pre-specifications

Pre-specification v1 (original method; executed, concluded ‘not better than chance’) and pre-specification v2 (frozen scheme; validation rules) are provided in English translation as Supplementary Appendices A and B (the deposited originals are in Chinese and are authoritative) and were deposited with a time-stamp before unblinding, together with the frozen configurations and the code manifest (Zenodo, doi:10.5281/zenodo.23439034; record created on 17 October 2026 at 16:19 UTC; SHA-256 of the deposited archive a06b55a96142d6fea1185ee562ecd2f3f226f20e76fb29588a4e62761936da49). The deposit exists in two versions. Version 1 is the original archive. Four of its 17 files refer to development recordings by the hospital’s internal EEG record numbers, so its files are under restricted access; editors and reviewers can obtain them on request. Version 2 is public (doi:10.5281/zenodo.23439033 resolves to the latest version). It is a copy in which each of the 37 occurrences of a record number is replaced by the string ‘[redacted record number]’ and nothing else is changed: 13 files are identical byte for byte, and a script in the archive verifies the other four against the original. The two frozen configurations of the warning scheme are among the unchanged files, so their embedded SHA-256 values can be recomputed from the public copy. The configuration of the confound-only comparator lists its training recordings, so its embedded SHA-256 and its file digest in Supplementary Table S6 can be reproduced only from version 1. Both pre-specifications were written on 29 September 2026, the day of the freeze, as internal documents. Before confirmation, v2 was amended on 6 October 2026 on five points, all recorded in its §12 with the reason and the statement that no validation data had been seen: (1) the clock-matched reference uses sliding interictal blocks whose centre lies within 2 h of the pre-ictal segment (the rule actually used for the development estimate), not 1.5 h; (2) the primary decision rule requires $p < 0.05$ on the analytic test and on the ±1 h local-jitter test, the uniform permutation being reported alongside, because the development cohort showed the uniform permutation to be the most lenient chance model; (3) the power table was recomputed for the 17 children then expected in batch 2 (about 36 evaluable seizures; extended on 14 October 2026 when 21 recordings were announced) and states that no cohort may be dropped after results are seen; (4) the strict-E4 permutation p of secondary operating point 1 was corrected to 0.042; (5) the training window of the weights was stated as 15–0.5 min before onset with equal weighting of children within each class. All five amendments were confirmed on 13 October 2026 (v2 §12), before any validation data had been processed. Entries of the v2 change log after confirmation (§12; each dated and marked as made without sight of any validation-set result): 13 October 2026, runner completed with the clock-matched test and the decision output (M2); 14 October, CHB-MIT adapter frozen from headers, annotation files and the frozen quality control alone (M4; Supplementary Methods S3); 14–15 October, pre-unblinding analyses A1 and A2 on the development cohort only (Notes S5 and S6); 15 October, the confound-only comparator added as key secondary endpoint 5 without a pass or fail criterion (Note S6); 17 October, inventory and quality control of cohort A without any prediction output, then the Zenodo deposit (record created 16:19 UTC, under an embargo that was lifted on 6 November 2026), then the six locked runs (16:22–16:43 UTC; log `logs/locked_validation_20261017_172207.log`), recorded after unblinding together with the finding that one delivered recording (D20) duplicated a development recording. No rule was deviated from. The deposited v2 ends with the entries of 15 October 2026; Appendix B translates the deposited version, and the later entries are in the live project document.

### Supplementary Appendix A. Pre-specification v1 (English translation of the deposited original)

*Translator’s note.* The original was written in Chinese on 29 September 2026 and deposited unchanged before unblinding (Zenodo, doi:10.5281/zenodo.23439034, file `prespecifications/prespecification_v1_2026-09-29.md`; SHA-256 of the deposit archive a06b55a96142d6fea1185ee562ecd2f3f226f20e76fb29588a4e62761936da49; the file is identical in versions 1 and 2 of the record, see Supplementary Note S3). This translation was prepared by the authors after the locked run for the reader’s convenience; the deposited original is authoritative. Section numbering, tables, numbers and file paths (which refer to the project repository) are reproduced unchanged. Chinese annotation labels are given in quotation marks with their meaning.

### EpiFound retrospective cohort experiment: pre-specification of the evaluation rules (v1)

**Frozen:** 29 September 2026, *before* any cohort prediction result (alarms, sensitivity, false-alarm rate) was seen. **Scope:** batch 1 (20 children) plus the pilot case IY7776, and later batches. **Principle:** any change of rule after this document must be recorded as a “deviation” in §12, stating what was changed, when, why, and whether results had been seen before the change.

Data already seen when this document was written: annotation text, recording durations, seizure times, artefact proportions, and the null behaviour of the interictal statistic z_t (mean, variance, exceedance rate, sleep–wake difference). **Not seen:** any pre-ictal z_t, any CUSUM alarm, any performance figure.

#### 1. Method version

The main analysis uses the current method (spc2 / v4B) without any change: spectral entropy + Hjorth mobility → per-channel Mahalanobis distance → probability integral transform with a Blom-corrected empirical CDF → fusion with non-negative weights derived from a Ledoit–Wolf covariance → one-sided CUSUM → alarm confirmed by N consecutive windows; NORMAL / POST_SEIZURE dual-mode sliding queue.

The engine `spc2.retro` was regression-tested on IY7776: its output is point-for-point identical to the original script (23 769 windows, maximum difference 0; all 46 re-estimations identical).

#### 2. Gold standard

**Seizure** = an event that the reviewing physician explicitly annotated as “发作” (seizure) or “seizure onset”. Several annotations of the same seizure (within 120 s and without an end marker between them) are merged into one; the onset is the earliest annotation (the most conservative choice for prediction).

| Category | Definition | Handling in the main analysis |
|---|---|---|
| Confirmed seizure | Annotation without a question mark | Counted as a seizure |
| Questionable seizure | Annotation marked “?” | **Not counted as a seizure**; treated as interictal |
| Provoked seizure | Confirmed, but within a hyperventilation or photic-stimulation period | Triggers the post-seizure lock-out; excluded from the sensitivity evaluation |
| Uncertain event | Parent-reported event, isolated “◆” bookmark, “electrographic evolution”, not annotated as a seizure by the physician | **Not counted as a seizure**; treated as interictal |

Rationale: of 47 parent-reported events only 3 coincide with a physician’s seizure annotation; for the others the physician re-watched the video and wrote an observation without judging it a seizure. Taking the physician’s judgement as the reference is standard practice in long-term video-EEG. Treating these events as interictal is conservative for us: any alarm before them counts as a false alarm.

#### 3. Periods excluded from the queue and from monitoring

| Period | Extent | Enters the baseline queue | Monitored / in the denominator |
|---|---|---|---|
| Post-seizure lock-out | 1 h after each confirmed seizure: `[s, s+360)` windows | No | No |
| Provocation tests | Hyperventilation: from 60 s before the first marker to 300 s after the last; photic stimulation: from 30 s before to 120 s after | No | No |
| Recording gaps | Missing or corrupt segments (3 in this batch, 2 h each) | — | — |
| Post-gap lock-out | 1 h after the end of a gap (whether a seizure occurred inside the gap is unknowable) | No | No |
| Post-alarm lock-out | 1 h after each alarm | As usual | No |
| Artefact windows | Failed the PHASE 0 quality control | No | No (S_t unchanged; the consecutive count is reset) |

Windows *before* a seizure enter the queue as usual (causal; identical to the IY7776 implementation).

#### 4. Calibration (initial baseline)

Admissible windows are accumulated from the start of the recording; calibration is complete when [2880 window slots have accumulated (a span of 8 h) **and** at least 1440 of them are clean]. If a confirmed seizure s occurs during calibration, the windows already counted within `[s−1 h, s)` are removed retroactively and no window within `[s, s+1 h)` is counted. The periods listed in §3 are not counted. A case that cannot complete calibration within its recording does not enter the prediction evaluation (the number of such cases is reported).

Rationale (`scripts/30_v5_development/exp_baseline_size.py`, interictal data only, 13 cases, 280 test points): baseline quality is determined by the **time span**, not by the number of windows. 720 windows drawn uniformly from 10.7 h (exceedance rate 9.4%) are equivalent to the full 2880 (9.8%), whereas 720 consecutive windows (spanning 2.7 h) give 15.1%.

#### 5. Rolling baseline

As implemented in the code: NORMAL queue of 2880 windows, re-estimated after every 1440 newly queued windows; from the first confirmed seizure after calibration the mode switches to POST_SEIZURE (irreversibly): queue of 720, re-estimated every 360; no re-estimation while the queue holds fewer than 500 windows. The z_t of the current window is computed with the parameters in force before the re-estimation.

#### 6. Evaluable seizures

A seizure enters the sensitivity denominator only if all of the following hold:

- **E1** confirmed (no question mark)
- **E2** spontaneous (not within a provocation period)
- **E3** occurs at least 90 min after calibration is complete
- **E4** at least 2.5 h after the preceding confirmed seizure (1 h lock-out + 90-min warning window)
- **E5** at least 50% of the windows in the warning window were recorded

Seizures are **not** excluded on the basis of pre-ictal data quality (artefact proportion): excluding “hard” seizures would inflate sensitivity.

Result for this batch: 128 seizure events → **41 evaluable**, in 18 of 21 cases.

#### 7. Alarms and scoring

The CUSUM increment uses **causal normalisation**: `inc_t = (z_t − μ_z − k) / σ_z`, with μ_z and σ_z the baseline parameters in force at that window. `S_t = max(0, S_{t−1} + inc_t)`; an alarm is raised when the number of consecutive valid windows with `S_t > h_eff` reaches N. `h_eff = −h·ln(1−θ)`.

Each case is simulated **once, continuously** (from the end of calibration to the end of the recording), with the following state transitions:

| Event | Handling |
|---|---|
| A confirmed seizure time is reached | S_t reset to 0, consecutive count reset, 1-h lock-out |
| Any alarm | S_t reset to 0, consecutive count reset, 1-h lock-out |
| Recording gap (adjacent windows more than 60 s apart) | S_t reset to 0, consecutive count reset |
| Artefact windows, provocation-test windows | S_t unchanged, consecutive count reset |

Warning window: `[0.5 min, 90 min]` before the seizure (APT_MIN = 3, APT_MAX = 540 windows).

| Alarm class | Definition | Counting |
|---|---|---|
| True alarm | Falls within the warning window of an **evaluable** seizure | The seizure counts as detected; the lead time is that of the **earliest** alarm in the window |
| Neutral alarm | Falls within the 90 min before a confirmed but **non-evaluable** seizure, or less than 0.5 min before an evaluable | Not counted |

| Alarm class | Definition | Counting |
|---|---|---|
| | seizure | |
| False alarm | All others (including alarms before questionable seizures and uncertain events) | False-alarm count |

#### 8. Metrics

- **Sensitivity** = detected evaluable seizures / evaluable seizures; Wilson 95% confidence interval
- **False-alarm rate** = false alarms / valid interictal monitoring time. Denominator = number of windows that passed quality control, were not locked out or excluded, and were not within 90 min before any confirmed seizure, ÷ 360. The pooled value and the per-case distribution (median, interquartile range) are both reported
- **Lead time**: median (interquartile range)
- **Time in warning** = proportion of time spent in the alert state after alarms (for comparison with a random predictor)

#### 9. Parameter selection: leave-one-patient-out cross-validation

Search space (fixed in advance):

| Parameter | Values |
|---|---|
| k | 0.30, 0.40, 0.50, 0.60, 0.70, 0.80, 0.90, 1.00, 1.10, 1.20 |
| h_eff | 14 geometrically spaced points from 2.0 to 40.0 |
| N | 1, 2, 3, 5, 8 |

For reporting, h = h_eff / 0.9163 at θ = 0.60.

**Stage 1 (ARL pruning):** with the median of the training cases' initial σ_z, compute k_norm = k/σ_z and keep the combinations whose Siegmund $ARL_0$ is at least 720 windows. **Stage 2 (empirical selection):** for each held-out patient, choose the parameters on the other patients: constraint, pooled training false-alarm rate ≤ 0.5/h; objective, maximal pooled training sensitivity; ties broken in turn by lower false-alarm rate, longer median lead time, larger k, larger h_eff. The chosen parameters are applied unchanged to the held-out patient. The final metrics pool the results of all held-out patients.

Calibrated cases without an evaluable seizure contribute to the false-alarm statistics but not to sensitivity.

#### 10. Statistical tests

1. **Better than chance:** (a) analytic: the expected sensitivity of a Poisson random predictor at the same time in warning, with a binomial test (Snyder 2008); (b) alarm-time surrogates: each case keeps its number of alarms, which are re-placed at random within the monitorable periods, 1000 times (Andrzejak 2009). Both are reported.
2. **False-alarm rate stratified by vigilance state** (sleep / wake). The interictal data had already shown a higher z_n in wakefulness (+0.62 versus −0.10); false alarms are expected to concentrate in wakefulness.
3. **Subgroups** (descriptive, no confirmatory inference): seizure burden, seizure duration, state at onset, scalp visibility, lateralisation / lobe of interictal discharges.

#### 11. Pre-specified sensitivity analyses and ablations

| Code | Content |
|---|---|
| S1 | Remove 1 h before and after every uncertain event (parent-reported, bookmark, questionable seizure, electrographic evolution) from the baseline and from the denominator |
| S2 | Lead-seizure criterion changed from 2.5 h to 4 h |
| S3 | Exclude the pilot case IY7776 (used during method development) |
| S4 | Normalisation as in the pilot experiment (the final re-estimated μ_z and σ_z used throughout) |
| A1 | Remove the probability integral transform: per-channel Mahalanobis distances standardised directly and then fused |
| A2 | Remove the covariance weights: simple mean of the per-channel z |
| A3 | Alarm-level fusion: per-channel CUSUM alarm counts (as in Zandi 2009) |
| A4 | Span-preserving sparse queue: in POST_SEIZURE mode draw 720 windows uniformly from the most recent 8 h |

A4 follows from the finding in §4; it is a candidate improvement of the current method, **not** the main analysis.

#### 12. Deviation log

| Date | Content | Reason | Results seen before the change? |
|---|---|---|---|
| — | — | — | — |

#### 13. Differences from the pilot experiment (IY7776)

| Item | Pilot experiment | This pre-specification | Note |
|---|---|---|---|
| Initial baseline | Set by hand to [S1 + 1 h, S1 + 9 h] | Accumulation rule of §4 | Under the uniform rule the calibration interval of IY7776 is [0, 10.12] h and the evaluable seizures remain S2, S4 and S8 |
| Increment normalisation | Final re-estimated $\mu_z$ and $\sigma_z$ used throughout | Parameters in force at the time | The pilot procedure is slightly non-causal; kept as S4 for comparison |
| Evaluation | One simulation per seizure; false-alarm rate averaged over seizures | One continuous simulation per case | The pilot procedure counts false alarms in the same period more than once |
| False-alarm denominator | Includes the 90 min before seizures | Excludes them | The false-alarm rate should be measured on interictal time |
| Alarms less than 0.5 min before a seizure | Counted as false | Neutral | Consistent with the previous row |
| Parameter grid | Config grid plus manual extension | Fixed grid of §9 | The pilot optimum lay outside the config grid |
| Documentation stating "±1 h around seizures not queued" | — | 1 h after a seizure | A documentation error; the code always used 1 h after the seizure; the documentation has been corrected |

### Supplementary Appendix B. Pre-specification v2 (English translation of the deposited original)

*Translator's note.* The original was written in Chinese, drafted on 29 September 2026, amended on 6 October and confirmed on 13 October 2026, and deposited unchanged before unblinding (Zenodo, doi:10.5281/zenodo.23439034, file `prespecifications/prespecification_v2_effective_2026-10-13.md`; SHA-256 of the deposit archive a06b55a96142d6fea1185ee562ecd2f3f226f20e76fb29588a4e62761936da49; in the public copy, version 2 of the record, one hospital record number is redacted in this file, see Supplementary Note S3). This translation was prepared by the authors after the locked run for the reader's convenience; the deposited original is authoritative. Section numbering, tables, numbers, dates, hashes and file paths (which refer to the project repository) are reproduced unchanged, except that a hospital record number by which the original names one recording is omitted. In the original, "用户" (the user) denotes the investigator who directed the analyses and confirmed each rule (M.C.); it is rendered as "the first author". Entries of the change log dated after 13 October 2026 belong to the live project document and are not part of the deposited file; they are summarised in Supplementary Note S3.

#### EpiFound v5p: method freeze and pre-specification of the validation rules (v2)

**Status: In force (13 October 2026; the first author confirmed, item by item, the five amendments of 6 October 2026).** Until the external time-stamped deposit (OSF/Zenodo) is made, no prediction evaluation may be run on any validation data; the runner was completed on 13 October 2026 with the fourth test of §7 and the decision rule of §8 (see §12). **Drafted:** 29 September 2026 **Frozen file:** `config/v5p_frozen.json` (sha256 `6f06a972…f341a1`, generated 29 September 2026 18:54 UTC); secondary endpoint 2 uses `config/v5p_frozen_H20.json` (sha256 `c246ab56…80b336`). The sha256 is the hash of the canonical JSON with the `sha256` field removed, verified by `v5p_run.py`.

**Relation to v1:** v1 (`docs/protocols/回顾性实验_评估规则预设_v1.md`) concerned the original method v4B; it has been executed and concluded "not better than chance". v2 concerns the improved method v5p. The gold standard, the excluded periods and the calibration rule follow v1; everything else is governed by this document.

#### 0. Development set and validation sets

| Data | Role | Note |
|---|---|---|
| Batch 1, 21 cases | **Development set** | Inspected repeatedly. Every design choice, weight and parameter of v5p comes from it. |

| Data | Role | Note |
|---|---|---|
| (including IY7776) | | Any number obtained on it can only be called a "development-set estimate" |
| Batch 2 (17 cases, expected shortly) | **Validation set A** | On arrival, placed in a separate directory; once this document is in force it is run **once only**, as in §9 |
| CHB-MIT (public; to be downloaded) | **Validation set B** | As above. Paediatric scalp EEG, native bipolar montage |
| Siena (public; to be downloaded) | Confound analyses only | Single recordings of only a few hours; the 8-h calibration cannot be completed, so no prediction evaluation |

Seen when this document was written: all data and results of the development set. **Not seen:** any data of validation sets A or B.

### 1. Method (frozen)

#### *1.1 Signal layer*

| Item | Value |
|---|---|
| Montage | 18-channel double-banana bipolar montage: Fp1-F3, F3-C3, C3-P3, P3-O1, Fp2-F4, F4-C4, C4-P4, P4-O2, Fp1-F7, F7-T3, T3-T5, T5-O1, Fp2-F8, F8-T4, T4-T6, T6-O2, Fz-Cz, Cz-Pz (T3/T4/T5/T6 = T7/T8/P7/P8). Referential data are obtained by subtraction |
| Sampling rate | Resampled to 256 Hz |
| Filtering | 50 Hz and 60 Hz notch (Q = 30); 0.5–45 Hz 4th-order Butterworth band-pass; all zero-phase |
| Windows | 10 s, non-overlapping, aligned to the start of the file |

#### *1.2 Quality control (any channel failing any rule → the whole window is invalid)*

| Rule | Threshold |
|---|---|
| Flat / dropped channel | Variance < 0.5 $\mu V^2$ |
| Extreme amplitude | Peak-to-peak > 3000 µV |
| High amplitude not slow-wave dominated | Peak-to-peak > 800 µV and δ (0.5–4 Hz) power fraction < 0.4 |
| Step / square wave | Peak-to-peak ÷ SD > 20 (0.5–45 Hz signal) |
| Burst EMG / movement | Peak-to-peak ÷ SD > 14 (0.5–100 Hz signal) |
| EMG dominated | $\log_{10}$(65–95 Hz variance ÷ 0.5–45 Hz variance) > 0 |
| Missing channel | Any channel missing |

The thresholds were calibrated against the original PHASE 0 quality control (development set, 473 565 windows: 81.6% valid versus 80.7% originally; window-wise agreement 86.0%). The calibration used signal-quality measures only and no seizure information.

#### *1.3 Vigilance-state estimation*

13 global features (EEG only): δ/β, θ/β, α/θ, relative γ, relative δ, total power, spectral entropy, Hjorth mobility, Hjorth complexity, occipital-minus-frontal α, central β/δ, first eigenvalue fraction, scalp high-frequency power (65–95 Hz). Standardised by the baseline queue's median / (1.4826·MAD) and clipped to ±8 → logistic regression (coefficients in the frozen file, trained on all 21 development cases) → median of the last 5 valid windows (causal) → ≥ 0.5 classified as sleep. Leave-one-patient-out validation on the development set: AUC 0.964, accuracy 91.9% (against the physicians' "sleep / wake" annotations, 7727 annotated windows).

#### *1.4 State-conditioned rolling baseline*

| Item | Value |
|---|---|
| Admissible windows | Valid, and not within a provocation period, the hour after a confirmed seizure or the hour after a recording gap. Windows *before* a seizure are admitted as usual |
| Queue | The most recent 8640 admissible windows (24 h of clean data) |
| Re-estimation | After every 360 new admissible windows; the current window uses the parameters from *before* the re-estimation |
| Bootstrap | The first 1440 admissible windows determine their state from their own statistics |
| Conditional statistics | Mean and SD of every monitored feature computed separately for the sleep and wake windows of the queue |
| State not covered | Fewer than 300 windows of the current state in the queue → the window is not monitored |

##### *1.5 Monitoring statistic*

7 global features (averaged over the 18 channels) and their frozen weights:

| Feature | Weight |
|---|---|
| Spectral entropy | +0.4758 |
| Hjorth complexity | −0.0234 |
| Hjorth mobility | −0.4291 |
| δ log power | +0.1010 |
| β log power | +0.3124 |
| Mean inter-channel \|r\| | −0.7275 |
| First eigenvalue fraction | +0.4184 |

x = state-conditioned standardised features (missing values set to 0, clipped to ±5); y = w·x. Origin of the weights: logistic regression (C = 0.05; positives are the windows 15–0.5 min before each lead seizure (87 windows), negatives the interictal windows more than 4 h from any seizure (at most 4000 per patient); every patient weighted equally within its class), fitted on the 20 calibrated development cases.

**Interpretation of the weights:** the mean |r| and the first eigenvalue fraction are highly correlated; their opposite signs are a consequence of collinearity, and their joint net effect is “falling synchrony → rising risk”. Individual weights should not be given a physiological interpretation.

##### *1.6 Probability integral transform*

$z = \Phi^{-1}(\hat{F}(y))$, where $\hat{F}$ is the empirical distribution (Blom correction) of y over the **same-state windows of the current block’s baseline queue** (at least 100 same-state windows in the queue; u clipped to $[10^{-6}, 1 - 10^{-6}]$). The y of the queue windows is recomputed after re-standardisation with the current block’s mean and SD.

##### *1.7 Alarms*

S_t = max(0, S_{t−1} + z_t − k); an alarm is raised when the number of consecutive valid windows with S_t > h_eff reaches N.

| Use | Warning window | False-alarm cap | k | h_eff | N | Frozen file |
|---|---|---|---|---|---|---|
| **Primary endpoint** | 0.5–30 min | 0.5/h | 0.50 | 7.971 | 8 | `v5p_frozen.json` |
| Secondary endpoint 1 | 0.5–30 min | 0.25/h | 0.75 | 7.971 | 8 | as above |
| Secondary endpoint 2 | 0.5–20 min | 0.5/h | 0.25 | 40.0 | 5 | `v5p_frozen_H20.json` |

Parameter selection: on the **out-of-sample** z series of the development cases (leave-one-patient-out), the combination with the highest pooled sensitivity under the false-alarm constraint.

State transitions:

| Event | Handling |
|---|---|
| A confirmed seizure time is reached | S reset to 0, consecutive count reset, 1-h lock-out |
| Any alarm | S reset to 0, consecutive count reset, lock-out of one warning-window length |
| Recording gap (adjacent windows more than 60 s apart) | S reset to 0, consecutive count reset |
| Invalid windows, provocation-test windows, windows whose state is not covered | S unchanged, consecutive count reset |

#### 2–4. Gold standard, excluded periods, calibration

As in v1 §2, §3 and §4, with two changes only:

- “clean windows” are defined by §1.2 of this document;
- the post-alarm lock-out equals the length of the warning window (1 h in v1).

The public data sets have no categories such as “questionable seizure”, “parent-reported event” or “provocation test”; their own seizure annotations are taken as given.

#### 5. Evaluable seizures

All of the following must hold:

- **E1** confirmed
- **E2** spontaneous (not within a provocation period)
- **E3** occurs at least one warning window after calibration is complete
- **E4** at least 1.5 h after the preceding confirmed seizure (1-h post-seizure lock-out + 0.5 h). **Computed from the annotated time stamps, interval ≥ 5400 s** (the development-set evaluation at the time judged this by subtracting 10-s window indices, which admitted one seizure with an actual interval of 5394 s through rounding; the development-set results under both conventions are listed in §8)
- **E5** at least 50% of the windows in the 90 min before the seizure were recorded (definition and implementation as in v1, identical to the development-set evaluation)

Seizures are not excluded on pre-ictal data quality.

#### 6. Metrics

| Metric | Definition |
|---|---|
| Sensitivity | Detected evaluable seizures ÷ evaluable seizures; Wilson 95% confidence interval |
| False-alarm rate | False alarms ÷ valid interictal monitoring time (excluding one warning window before every confirmed seizure) |
| Lead time | Time from the earliest alarm in the window to the seizure; median (interquartile range) |
| Segment-level AUC | Pre-ictal segments (one warning-window long) against non-overlapping interictal segments more than 4 h from any seizure; segment score = mean of z; computed per patient |
| Sensitivity–time-in-warning curve | Threshold on z after 5-min causal smoothing; after a crossing the alert is held for one warning-window length |

#### 7. Statistical tests

Every performance figure is reported together with all four of the following:

| Test | Null hypothesis |
|---|---|
| Analytic | A Poisson random predictor at the same false-alarm rate: $P = 1 - \exp(-FPR \times \text{warning-window length})$; one-sided binomial test |
| Alarm-time permutation | Each case keeps its number of alarms, re-placed uniformly over the monitorable windows, 1000 times (minimum spacing preserved) |
| Local jitter ±1 h, ±3 h, ±6 h | Each alarm is moved only within ±J of its original time, 2000 times: slow trends and the diurnal rhythm are preserved and only the fine timing is disrupted |
| Clock-matched AUC | Pre-ictal segments are compared only with sliding interictal blocks (step 10 min) of the same patient, more than 8 h from any seizure, whose centre is within 2 h of the clock time of the pre-ictal segment's centre, with at least 5 blocks (the rule actually used for the development-set estimate; changed from 1.5 h to 2 h on 6 October 2026) |

#### 8. Primary endpoint and decision rule

**Primary endpoint:** on the validation sets, the sensitivity at the primary-endpoint parameters (first row of §1.7) exceeds the analytic chance level. **Decision:** one-sided $p < 0.05$ (analytic) **and** $p < 0.05$ (±1 h local jitter). The p-value of the uniform alarm-time permutation is reported alongside but is not part of the decision: the development set had shown it to be the most lenient of the four chance models (amendment of 6 October 2026; the validation sets had not been obtained at the time).

**Key secondary endpoints** (reported in this order, without multiplicity adjustment, labelled as such):

1. local-jitter ±3 h test, $p < 0.05$
2. per-patient segment-level AUC > 0.5 (one-sided Wilcoxon signed-rank test)
3. sensitivity of secondary endpoints 1 and 2 against their chance levels
4. clock-matched AUC

**Published whatever the outcome.** A failed validation is also a conclusion: it shows that the signal on the development set does not generalise.

*Corresponding estimates on the development set (nested leave-one-patient-out; 45 evaluable seizures; for comparison)*

| | Sensitivity | False-alarm rate | Chance level | p (analytic / permutation) | Local jitter ±1 / ±3 / ±6 h | Median lead |
|---|---|---|---|---|---|---|
| Primary endpoint | 15/45 = 33.3% (21.4–47.9%) | 0.473/h | 20.8% | .034 / ≈ .002 | .030 / .031 / .022 | 14.7 min |
| Secondary 1 | 10/45 = 22.2% | 0.244/h | 11.3% | .026 / .005 | .10 / .042 / .043 | 15.8 min |
| Secondary 2 | 11/45 = 24.4% | 0.414/h | 12.6% | .021 / .008 | .036 / .027 / .029 | 8.7 min |

Segment-level AUC: per-patient mean 0.672 (median 0.714; 10 of 13 cases > 0.5; p = .024), pooled 0.641. Sensitivity at 20% / 30% time in warning: 37% / 46% (interpolated; nearest grid points 20.4% → 37.8% and 30.7% → 46.7%).

**Development-set estimates with E4 applied strictly by time stamp** (44 evaluable seizures; the whole procedure re-run, so the weights and per-fold parameters change slightly):

| | Sensitivity | False-alarm rate | Chance level | p (analytic / permutation) | Local jitter ±1 / ±3 / ±6 h |
|---|---|---|---|---|---|
| Primary endpoint | 16/44 = 36.4% (23.8–51.1%) | 0.483/h | 21.1% | .015 / .001 | .009 / .027 / .013 |
| Secondary 1 | 8/44 = 18.2% | 0.269/h | 12.4% | .17 / .042 | .15 / .09 / .14 |
| Secondary 2 | 8/44 = 18.2% | 0.435/h | 13.2% | .22 / .10 | not run |

Segment-level AUC 0.673 / 0.643, almost identical to the table above. **The primary endpoint is significant under both conventions; the two secondary endpoints are significant only under the original convention and are not robust to the inclusion or exclusion of a single seizure.**

##### How to read these numbers

- They are nested leave-one-patient-out estimates of the *whole procedure*: in every fold the weights and CUSUM parameters come from the other patients, and the folds' parameters differ from one another (of the 20 primary-endpoint folds only 3 happen to equal the frozen parameters). They are not the performance of the frozen parameters themselves; the frozen parameters' result on the development set (19/45) is in-sample and must not be quoted.
- In the permutation test 0 of 1000 permutations reached the observed value, so only p ≤ .001 can be stated; the estimate from 20 000 permutations is .002–.003.
- The analytic p of the primary endpoint is marginal: 15/45 is exactly the smallest number of detections giving p < .05; one detection fewer gives .068.
- The median lead of 14.7 min is not evidence: random alarms falling into a 0.5–30 min window also have a median lead of about 15 min.

**What these numbers mean:** the great majority of the alarms the system raises are false; one seizure in three is warned of in advance. It is statistically better than chance but far from clinically usable.

#### 9. Power

One-sided α = 0.05, primary endpoint (chance level 20.8%):

| Evaluable seizures in the validation set | Detections needed | Power at a true sensitivity of 33.3% | Power at half the effect (27.1%) |
|---|---|---|---|
| 36 (expected for batch 2 alone, 17 cases) | ≥ 13 | 0.42 | 0.15 |
| 45 | ≥ 15 | 0.56 | 0.21 |
| 50 (CHB-MIT, actual evaluable count; counted on 14 October 2026 from the annotations and the frozen quality control) | ≥ 16 | 0.63 | 0.26 |
| 65 | ≥ 20 | 0.71 | 0.29 |
| 86 (batch 2 expected 36 + CHB-MIT actual 50) | ≥ 25 | 0.83 | 0.38 |
| 95 (batch 2 with 21 cases, expected 45 + CHB-MIT actual 50; case number communicated by the hospital on 14 October 2026) | ≥ 27 | 0.87 | 0.42 |
| 100 | ≥ 29 | 0.85 | 0.37 |
| 130 (pooled expectation at drafting; after the CHB-MIT count the pooled expectation is 86) | ≥ 36 | 0.93 | 0.47 |
| 160 | ≥ 43 | 0.97 | 0.55 |

**Risk:** development-set estimates are usually optimistic. If the true effect is only half as large, the power is only about 0.5 even with both validation sets pooled. **There is a substantial chance that the validation will not reach significance.** Validation sets A and B are pooled as the primary analysis (reported by stratum as well) to obtain the largest possible sample. **No cohort may be dropped after any result has been seen.**

#### 10. Comparators

Re-implemented on the same data under the same evaluation rules; each method is reported under its "published convention" and under "this paper's convention":

| Method | Source |
|---|---|
| Original EpiFound (v4B) | This project; results under pre-specification v1 already exist |
| Segal-type criterion | Segal 2025; with seizure-free control epochs |
| MSPC ($T^2$ / Q) | Rukhsar 2019 |
| v5p without state conditioning | Ablation |
| v5p with an unsigned statistic | Ablation |

Corresponding development-set results (30 min, false-alarm cap 0.5/h): full system 15/45 (p = .034 / ≈ .002), AUC 0.672; without state conditioning 11/45 (p = .29 / .086), AUC 0.641; unsigned statistic 9/45 (p = .55 / .50), AUC 0.536; MSPC (this paper's convention) 4/45 at 0.579/h (p = .998).

#### 11. Execution procedure

1. The first author confirms this document → the time it comes into force is recorded in §12; from then on the frozen file may not be changed (the sha256 is the evidence). Filtering, quality-control thresholds, clipping and the like are written in the scripts rather than in the frozen file, so when the document comes into force `config/v5p_code_manifest.json` (the sha256 of the relevant scripts) is regenerated and its hash recorded in §12; any later change to these scripts is recorded as a deviation.
2. A validation set arrives → it is placed in a separate directory → data quality control first (annotation format, channels, sampling rate, gaps), **without looking at any prediction output**.
3. The frozen procedure is run once; the outputs are written to disk and the time recorded.
4. Comparators and ablations are run only after the primary-endpoint results have been written to disk.

#### 12. Entry into force and deviation log

| Date | Item | Content | Validation-set results seen before the change? |
|---|---|---|---|
| 29 Sep 2026 | Drafted | This document | No (validation sets not yet obtained) |
| 29 Sep 2026 | Pre-confirmation revision (after independent re-check) | ① E4 made explicit as computed from time stamps; ② the permutation p restated as ≈ .002; ③ the strict-convention development-set estimates and four reading notes added to §8; ④ time-in-warning figures unified to the interpolated convention; ⑤ first power row of §9 0.55 → 0.56; ⑥ the hash of the H20 frozen file recorded. The frozen file itself was not changed | No (validation sets not yet obtained) |
| 6 Oct 2026 | Pre-confirmation revision (2) | ① §7 clock-matched reference changed to ±2 h sliding blocks (the rule actually used for the development-set estimate); ② §8 decision changed from "analytic ∧ uniform permutation" to "analytic ∧ ±1 h local jitter", the uniform permutation being reported only; ③ §9 power row for 17 cases (36 seizures) added, with the statement that no cohort may be dropped; ④ §8 strict-convention permutation p of secondary endpoint 1 corrected from .05 to .042 (summary.json); ⑤ §1.5 positive-class definition stated as 15–0.5 min with each patient weighted equally within its class; §0 batch 2 changed to 17 cases | No (validation sets not yet obtained) |

| Date | Item | Content | Validation-set results seen before the change? |
|---|---|---|---|
| 13 Oct 2026 | **In force** | The first author confirmed that all five amendments (①–⑤) are adopted. On the same day the project repository was changed to a src layout (logic verified unchanged by regression: the frozen runner re-run on the development set reproduced all 270 summary fields, the alarm table and the z series item by item); the code manifest `config/v5p_code_manifest.json` was regenerated for the new layout (31 files, including the versions of the run-time environment), sha256 = `281c7501ba18d0d9776e255b0460fbc627c8576788de1011ea4ca22aa1c5d167`; the old manifest is kept at `config/archive/v5p_code_manifest_2026-09-29.json` | No (validation sets not yet obtained) |
| 13 Oct 2026 | Runner changed after entry into force and before validation (M2) | `scripts/40_v5p_frozen/v5p_run.py` gained: ① the fourth test of §7, the clock-matched segment-level AUC (new module `src/epifound/v5p/clockmatch.py`; ±2 h sliding blocks, more than 8 h from a seizure, at least 5 blocks; reproduces item by item the seven horizons of a6_out S1 on the development set's out-of-sample z, e.g. 30 min 0.664 → 0.641, $p = 0.082$); ② the §8 decision output (analytic ∧ ±1 h jitter; the uniform permutation reported only) and the key secondary endpoints in fixed order. No other logic changed. The code manifest was regenerated (31 files), sha256 = `789fa3fb544a8eb0ad4ac334298a9eb73e7b21841c7fb320f12ab4e3dfef2408` | No (validation sets not yet obtained; the in-sample development run served only to test the procedure) |
| 14 Oct 2026 | After entry into force and before validation: CHB-MIT adapter frozen (M4) | Based only on the EDF headers, the summary annotation files and the frozen quality control; no state, baseline, z or alarm was computed. Rules: ① one subject directory = one case (24 cases; chb21 and chb01 are the same child recorded 1.5 years apart, handled as two cases and reported as 23 children); ② files ordered by the EDF header clock (the header date is a de-identified substitute date while the time of day is real, consistent file by file with the summary times; the summary of chb24 has no times); an interval of more than 60 s between adjacent files is recorded as a GAP (same rule as batch 1); ③ channels: 683 files are natively the 18 longitudinal bipolar pairs (label mapping FP1 → Fp1, T7 → T3, T8 → T4, P7 → T5, P8 → T6, FZ/CZ/PZ; duplicate labels take the first), while chb12_27–29 are referential recordings (CS2 or unspecified reference; in chb12_28/29 O1 is labelled "01") derived by subtraction with `bipolar_from_ref`, which cancels the reference; ④ seizure = every entry | No (no prediction output of any kind) |

| Date | Item | Content | Validation-set results seen before the change? |
|---|---|---|---|
| | | in the summary, E1 = E2 = 1 (no provocation tests recorded); E5 as defined in v1 (at least 50% of the windows in the 90 min before the seizure recorded); E4 ≥ 5400 s by time stamp; calibration by the v5p_meta rule; no case excluded; ⑤ missing ECG channel (ecg = NaN): verified on one development case that state, baseline coverage, z and the evaluable seizures are unchanged item by item. Inventory: 686 files, 982.8 h, 75.9% of windows valid under the frozen quality control (development set 81.6%), 22 of 24 cases completed calibration (chb12 and chb24 had too few clean windows during calibration), 198 annotated seizures, 50 evaluable (17 cases). Code: `src/epifound/v5p/chbmit.py`, `scripts/40_v5p_frozen/chbmit_inventory.py`, `chbmit_extract_features.py`, `chbmit_build_cohort.py`, `scripts/60_paper/verify_chbmit.py`; processed directory `data/processed_chbmit/`; the code manifest was regenerated (36 files), sha256 `52d0790cd7ad4f5851e3d431d0edc4b1a296e9cacf1f0f42278496e11cb0fcb2`. Rows for 50 and 86 added to §9 | |
| 14–15 Oct 2026 | After entry into force and before validation: pre-unblinding supplementary analyses A1 and A2 (approved by the first author on 14 October; development set only, no validation data touched) | **A1, simulation calibration of the chance tests** (`scripts/30_v5_development/a1_null_calibration.py`; three null models without a pre-ictal effect, 300 replicates each, alarm parameters per fold): false-positive rate of the decision rule (analytic $\wedge$ ±1 h jitter) 2.0% / 3.0% / 0.3% (null models A / C / B), analytic 3.3 / 3.0 / 0.3%, ±1 h jitter 5.0 / 8.3 / 4.7%, ±3 h 1.7 / 2.3 / 1.3%; uniform permutation 15.3 / 16.7 / 3.7%; per-patient segment AUC 31.7 / 35.0 / 3.3%; clock-matched AUC 16.7 / 19.7 / 6.7%. **Interpretative note (no change of rule):** the nominal p of key secondary endpoints 2 (segment AUC) and 4 (clock-matched AUC) is clearly too lenient under the clock-time, state and admission-day structure; at validation they are computed and reported as usual but are descriptive only and take part in no decision; the decision rule and secondary endpoints 1 and 3 are unchanged. **A2, confound-only comparator predictor** (`a2_confound_predictor.py`; two circadian harmonics of clock time + this scheme's state estimate + days since admission; trained leave-one-patient-out, same scoring, same tests): with nested parameter selection 13/45 = 28.9% at 0.410/h (analytic p .055, permutation .024, ±1 h jitter .130; | No (no validation-set output of any kind) |

| Date | Item | Content | Validation-set results seen before the change? |
|---|---|---|---|
| | | decision rule not met); segment AUC 0.594 versus 0.672, paired difference over 13 cases +0.078, p .19; with in-sample optimal parameters 16/45 at 0.410/h, passing all tests. The conclusions were written into the manuscript (statistical methods, results, discussion, Supplementary Notes S5/S6, one more row in Fig. 3A) | |
| 15 Oct 2026 | After entry into force and before validation: **key secondary endpoint 5 added (approved by the first author, P17)**: the confound-only comparator predictor | Implemented and frozen: `src/epifound/v5p/confound.py` + `scripts/40_v5p_frozen/confound_freeze.py` → `config/confound_frozen.json` (sha256 `98dcef7b011e97bbf4e40bf0386e3d6593096e0243bc9fe13aa62704fee1bb60`, verified at run time). Content: logistic regression on 8 non-EEG inputs (two circadian harmonics of clock time, days since admission, this scheme's EEG state estimate, state × first harmonic), fitted on all development cases with the same labels as the frozen weights (15–0.5 min before the first seizure positive, more than 4 h from a seizure negative, each case weighted equally within its class, C = 1); probability integral transform against a fixed per-state quantile grid of the development-set scores (2001 points; not adapted per patient; causal); CUSUM parameters chosen on the pooled development z_c over 420 combinations without pruning, the highest sensitivity at a false-alarm rate ≤ 0.5/h: k = 0.5, h_eff = 40.0, N = 8; lock-outs and scoring as at the primary operating point. The runner `v5p_run.py`, sections 2b/3b: computed on exactly the same monitored windows as the frozen scheme (make_series), with the same scoring and chance references (analytic, permutation, ±1 h jitter), and the segment-level AUC output (nominal); item 5 of the §8 decision block reports it side by side, **with no pass/fail**, and it takes no part in the decision. In-sample development-set procedure check (DEV_insample_P17, not to be quoted): comparator 17/44 at 0.466/h, frozen scheme 18/44. The out-of-sample development reference remains the leave-one-patient-out result of A2 (13/45 at 0.410/h). The code manifest was regenerated (40 files), sha256 `90330c496d0f9ef7d7f27807734a45567f258764b77bead630b2b2eb94e2c409` | No (no validation-set output of any kind) |
| — | To do (before validation) | ① external time-stamped deposit of v1/v2 (the deposit pack in `docs/deposit/` was prepared on 14 October 2026 and awaits upload by the first author and the recording of its | No |

| Date | Item | Content | Validation-set results seen before the change? |
| --- | --- | --- | --- |
| | | identifier here); ② secondary endpoint 2 to be run once, separately, with `config/v5p_frozen_H20.json`; ③ on arrival of batch 2, inventory and quality control only | |
